# CSF Hypo-Inflammation Drives Mortality in HIV-Associated Tuberculous Meningitis

**DOI:** 10.1101/2025.09.24.25336544

**Authors:** Anna L. Wilt, David B. Meya, Fiona V. Cresswell, Abduljewad Wele, Mable Kabahubya, Enock Kagimu, Jane Gakuru, Timothy Mugabi, Sarah Kimuda, Suzan Namombwe, Asmus Tukundane, Elizabeth C. Okafor, Biyue Dai, Nicole Engen, Nathan C. Bahr, David R. Boulware, Tyler D. Bold

**Author notes:** **Corresponding author:** Tyler D. Bold, 2101 6th St SE – WMBB 2-118, Minneapolis, MN 55455, USA.

## Abstract

**Background:** Excessive CNS inflammation is associated with poor outcomes in tuberculous meningitis (TBM). Anti-inflammatory corticosteroid therapy improves survival in HIV-negative TBM, but not in people with HIV. In people with HIV, TBM is fatal in 40-50% of cases. Therefore, we investigated how mortality is associated with the local immune dynamics in people with HIV-associated TBM.

**Methods:** We measured baseline concentrations of immune signaling mediators in cerebrospinal fluid (CSF) of 149 adults with HIV in Uganda, who presented with definite or probable TBM. Participants received both antimycobacterial and corticosteroid therapy.

**Results:** Non-survivors had more severe TBM disease stage and lower blood CD4 T cells than survivors at baseline. Mortality at 90 days was strongly associated with CSF hypo-inflammation. CSF interferon gamma (IFN-γ) was the cytokine most differentially expressed by survivors (2.2 log_2_-fold change higher, p=.003), and 90-day mortality was lower with increasing concentrations (Tertile-1=50%, Tertile-2=41%, Tertile-3=18%; p=.006). Even among people who successfully mounted a CSF cellular immune response (>5 white cells/μL CSF), those with low CSF IFN-γ had higher risk of death (Hazard Ratio =3.10 (1.44– 6.68). In contrast, people with intermediate CSF interleukin-13 had lower mortality than extreme (Tertile-1=45%, Tertile-2=22%, Tertile-3=40%; p=.017). Of all sub-groups, those with both peripheral CD4 depletion and low CSF IFN-γ had the highest mortality (63%).

**Conclusions:** In adults with HIV-associated TBM receiving dexamethasone, mortality was strongly associated with CSF hypo-inflammation. Although steroids may be appropriate in those with high inflammation, personalized approaches to immunotherapy are likely necessary to improve outcomes.

## Background

Protection against *Mycobacterium tuberculosis (Mtb)* infection relies on a coordinated immune response that generates sufficient inflammation to control bacterial growth but avoids excessive tissue damage—a balance modeled by the damage-response framework of microbial pathogenesis [1]. If the immune system fails to contain pulmonary infection, *Mtb* can disseminate to other organs including the central nervous system (CNS) [2]. Tuberculous meningitis (TBM) is the deadliest form of TB: nearly 50% of adults who develop TBM succumbing to the disease and over 30% of survivors being left with neurological deficits [3,4]. People with HIV co-infection fare considerably worse than those without HIV, with mortality rates approaching 70% in some settings [3]. This increased risk likely results from HIV-associated depletion of CD4 T cells, which are essential for controlling *Mtb* [5,6]. However, even before significant CD4 loss, people with HIV are more susceptible to TB [7]. The complex interplay between these two infections likely impacts the immunopathogenesis of HIV-associated TBM [8].

Several studies have characterized immune signaling mediators in the cerebrospinal fluid (CSF) of patients with TBM [9–17]. However, relatively few of these have focused specifically on people with HIV [18–20]. We previously found that people with a higher CSF bacterial burden, associated with higher levels of inflammatory CSF cytokines, had worse outcomes than those with an intermediate or lower burden of CNS *Mtb* [21]. Hyper-inflammation contributes to immunopathology, and adjunctive corticosteroid treatment benefited Vietnamese adults with TBM who did not have HIV [22]. But, it has not been found to benefit people with HIV [23]. In Vietnam, in people without HIV, the *LTA4H* TT genotype that causes excessive CSF inflammation is associated with the greatest benefit of adjunctive dexamethasone treatment [11,14]. However, in this same Vietnamese population with TBM, among people without HIV, non-survivors were found to have lower baseline concentrations of 10 CSF cytokines. In contrast, among those with HIV co-infection, survivors and non-survivors did not differ in CSF cytokine expression [11]. The generalizability of these findings to other populations where HIV and TBM are prevalent, including in African countries, remains unknown.

We hypothesized that the local immune response in HIV-associated TBM contributes to disease outcome and could identify subgroups of individuals at higher risk of death. To test this, we studied baseline CSF immune responses in 149 Ugandan adults with HIV-associated definite or probable TBM, profiling 50 cytokines, chemokines, and growth factors.

## Materials and Methods

### Study design

Baseline CSF specimens from individuals treated for HIV-associated TBM were obtained via lumbar puncture from participants who provided informed consent for CSF specimen collection; appropriate IRB approvals occurred. Enrolled individuals were included in this study if the met the consensus case definition for either definite TBM (microbiologically-confirmed by CSF AFB smear, culture (Mycobacterial Growth Indicator Tube, Becton Dickinson) or Xpert MTB/RIF Ultra PCR (Cepheid)) or probable TBM, having negative CSF microbiology but meeting the uniform case definition for probable TBM [24]. Persons not meeting these definitions (e.g., possible or not TBM) were not included. All individuals enrolled received treatment for TBM with standard quadruple anti-mycobacterial therapy and adjunctive corticosteroids which was the standard of care during this period.

### CSF collection and processing

After collection, CSF was centrifuged, and supernatant stored at −80°C. Cryopreserved CSF was transported at −20°C to Minnesota for batched analysis using custom 10- and 40-plex Human Luminex Discovery Assays (R&D, Minneapolis, MN). Per the manufacturer’s protocol, CSF was diluted 1:2 and analyzed with the Luminex MAGPIX instrument and Quantist Analysis Software (R&D, Minneapolis, MN).

### Data processing

To correct for technical limitations across Luminex assay runs, concentrations were adjusted according to a uniform set of predefined rules. Values that differed by more than two-fold from the next ranked value were considered outliers and were set to one unit above the next highest detectable value in rank order. Values below the limit of detection were set at the next non-outlier minimum. Additionally, three analytes—IL-25, IL-31, and EGF—were excluded due to undetectable levels in >40% of participants.

### Statistical analysis

All analyses were performed using R version 4.4.3 and were limited to only participants with HIV-associated definite or probable TBM. Among those who died within 90-days and survivors who completed 90-day follow-up, participants were grouped by 90-day survival status. Demographic and clinical variables were compared between survivors and non-survivors using the Wilcoxon rank-sum test for continuous variables and Pearson’s chi-squared or Fisher’s exact test for categorical variables. Raw concentrations of the CSF immune mediators were compared between survivors and non-survivors, using the Wilcoxon rank-sum test. Log2-fold changes were calculated quantify the direction and magnitude of expression differences. For analytes found to be significantly different among survivor and non-survivors, Pearson’s correlation coefficient was calculated to assess the correlation among themselves and with key clinical parameters. These variables were further dichotomized using either clinically established thresholds or cutoffs derived from receiver operating characteristic (ROC) curves based on the binary 90-day survival status. ROC-based cutoffs were identified via optimizing the Youden’s index, which combines sensitivity and specificity. Survival was then compared between the resulting CSF inflammation groups among all individuals, where survivors who did not complete 90 days were also included and censored at the last day of follow up. Kaplan-Meier curves were generated and differences in survival were assessed using the log-rank test. Cox proportional hazards models were used to estimate hazard ratios (HRs). Statistical significance was defined as p<0.05. Benjamini-Hochberg false discovery rate (FDR), or q-values, were also reported to account for multiple testing.

## Results

### Non-survivors have more severe CD4 T cell depletion

Among 149 adults with HIV-associated definite or probable TBM in Uganda, 54 (36%) were non-survivors who died within 90 days of presentation. Of the others, 67 (45%) were 90-day survivors followed for the entire study period, while 28 (19%) were not followed for the full 90-day study period, being censored after hospital discharge. To identify features associated with mortality, we compared baseline clinical and laboratory characteristics between non-survivors and survivors who completed the 90-day follow up period (**Table 1**). The 90-day follow-up cohort included 61 participants with definite TBM and 60 with probable TBM. Non-survivors presented with more severe disease: 57% (30/53) had a Glasgow Coma Scale score ≤12 at enrollment compared to 29% (19/66) of survivors (p=.009), and 37% (19/51) were classified as MRC grade III compared to 14% (9/66) of survivors (*p=.*007). Non-survivors had lower peripheral blood CD4 T cell counts (median 49 vs. 117 cells/μL; *p=.*004). Fewer non-survivors than survivors had a blood CD4 count >100 cells/μL (24% (9/37) vs. 60% (37/62); *p*<.001). Despite this, antiretroviral treatment status at diagnosis did not differ significantly between these groups (p=.2), and the proportion who initiated HIV therapy within 30 days of TBM diagnosis was also similar (32% (10/31) vs. 24% (4/27)). These findings suggested that impaired CD4 T cell immunity contributes to disease severity and mortality in HIV-associated TBM.

**Table 1.**
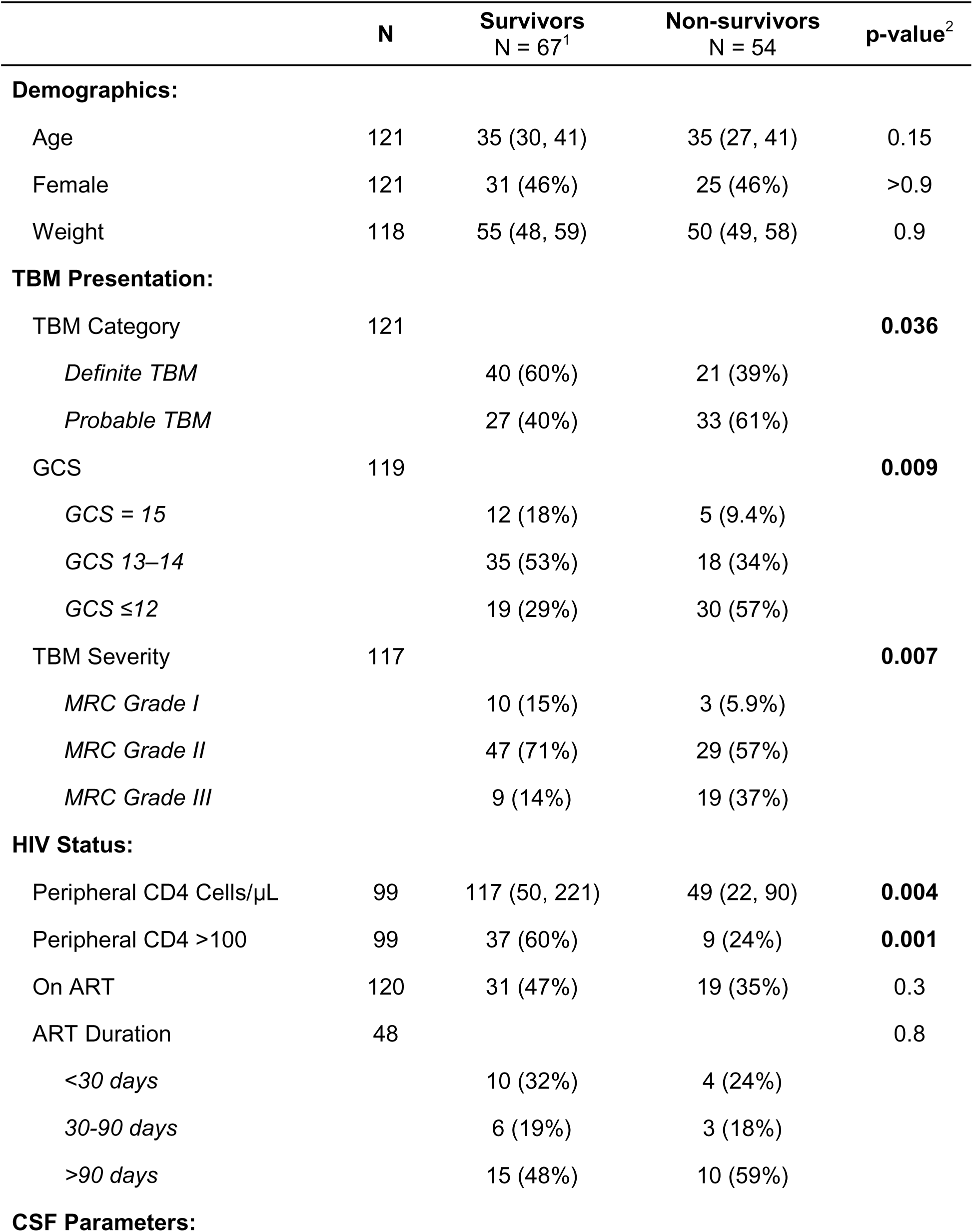
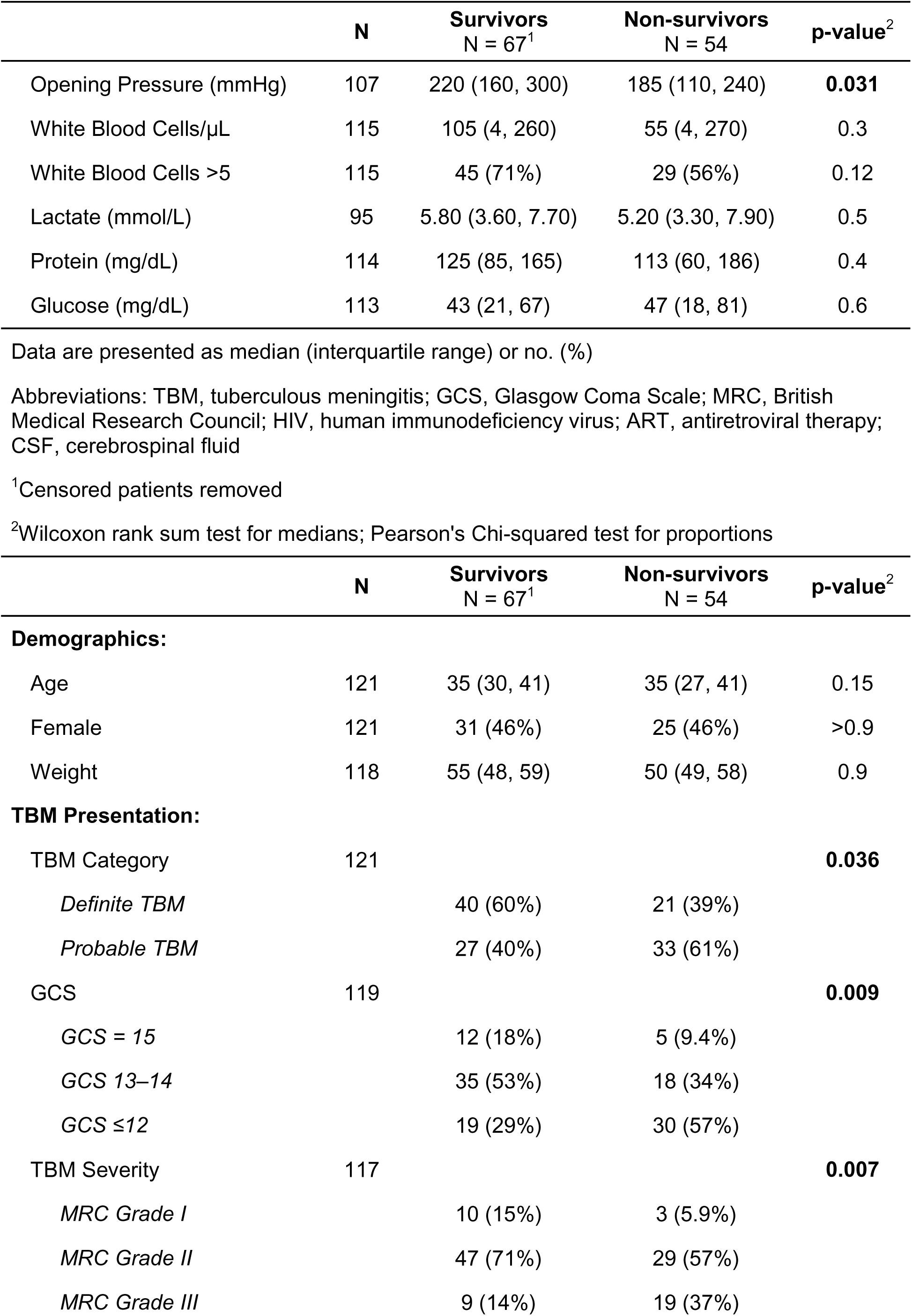
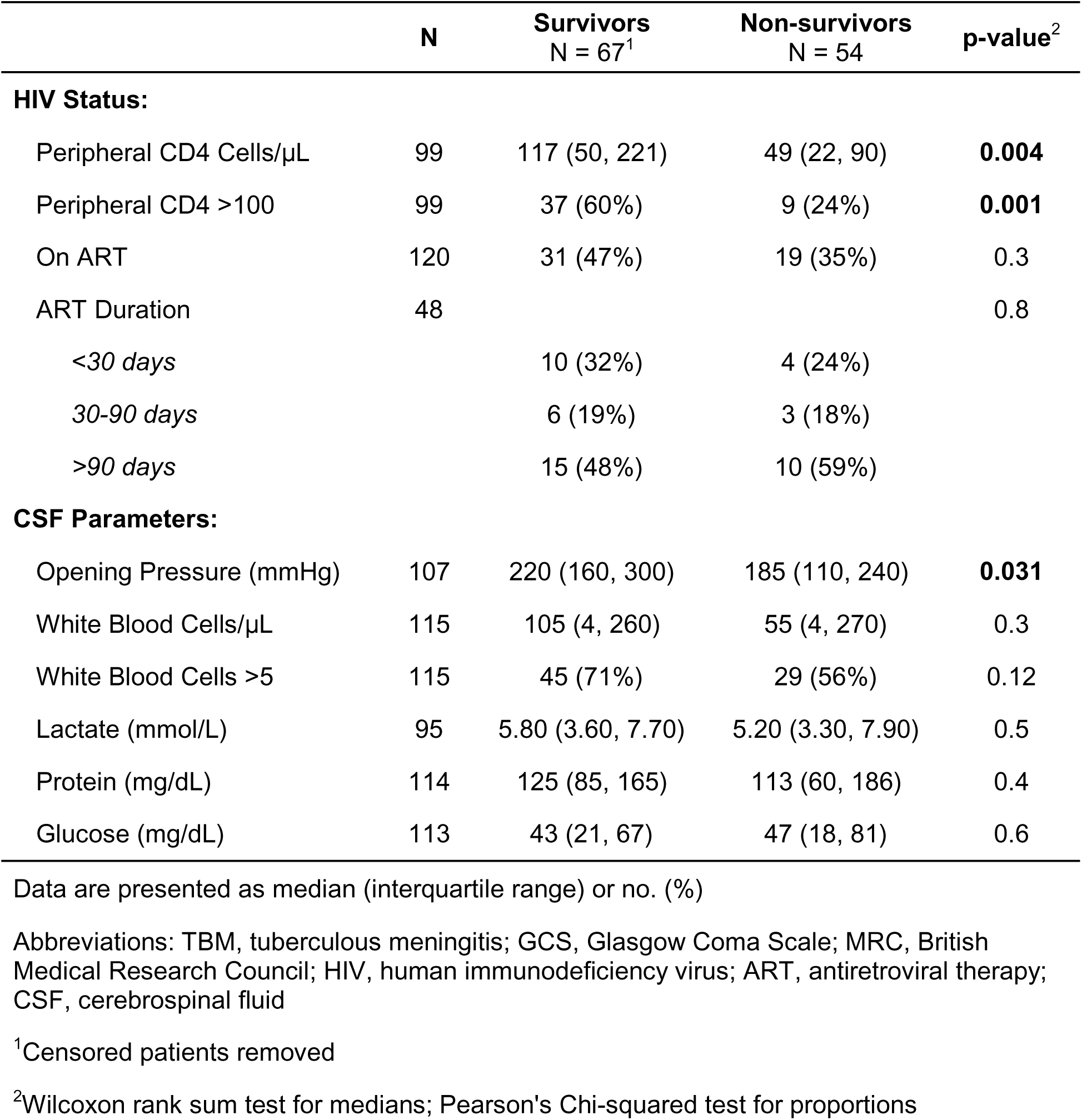
Comparison of Baseline Demographics, Clinical Characteristics, and Cerebrospinal Fluid Parameters Between 90-day Survivors and Non-survivors with HIV-associated TBM.

### Non-survivors have deficient CSF Th1 immunity

Given the marked peripheral CD4 T cell depletion in non-survivors, we next examined whether the difference between these groups in immune status also manifested locally in the CNS, by comparing baseline CSF immune analyte levels. This analysis revealed that mortality is associated with a deficient CNS immune response (**Table 2**, **Fig. 1A**). Of the 47 analytes measured, 20 were significantly different between the two groups, and all of these were present at lower concentrations in non-survivors than survivors (**Fig. S1**). The most pronounced difference was in IFN-γ (**Fig. 1B)**, with a median concentration over four-fold lower in non-survivors compared to survivors (57 vs. 251 pg/μL; *p<*.001). CSF concentrations of several other Th1-associated cytokines including TNF-α, IL-2, IL-12, and GM-CSF were also significantly lower in non-survivors (**Fig. S1)**. In contrast, Th2 cytokines including IL-13, IL-4, and IL-5 did not differ between the groups (**Figs. 1B and S1**). We therefore calculated the ratio of CSF IFN-γ/IL-13 production and found that it was significantly lower in non-survivors (**Fig. 1B**). These results suggest that, in contrast to people without HIV co-infection, mortality in HIV-associated TBM stems from inadequately protective local immunity, not excessive inflammation.

**Figure 1.**
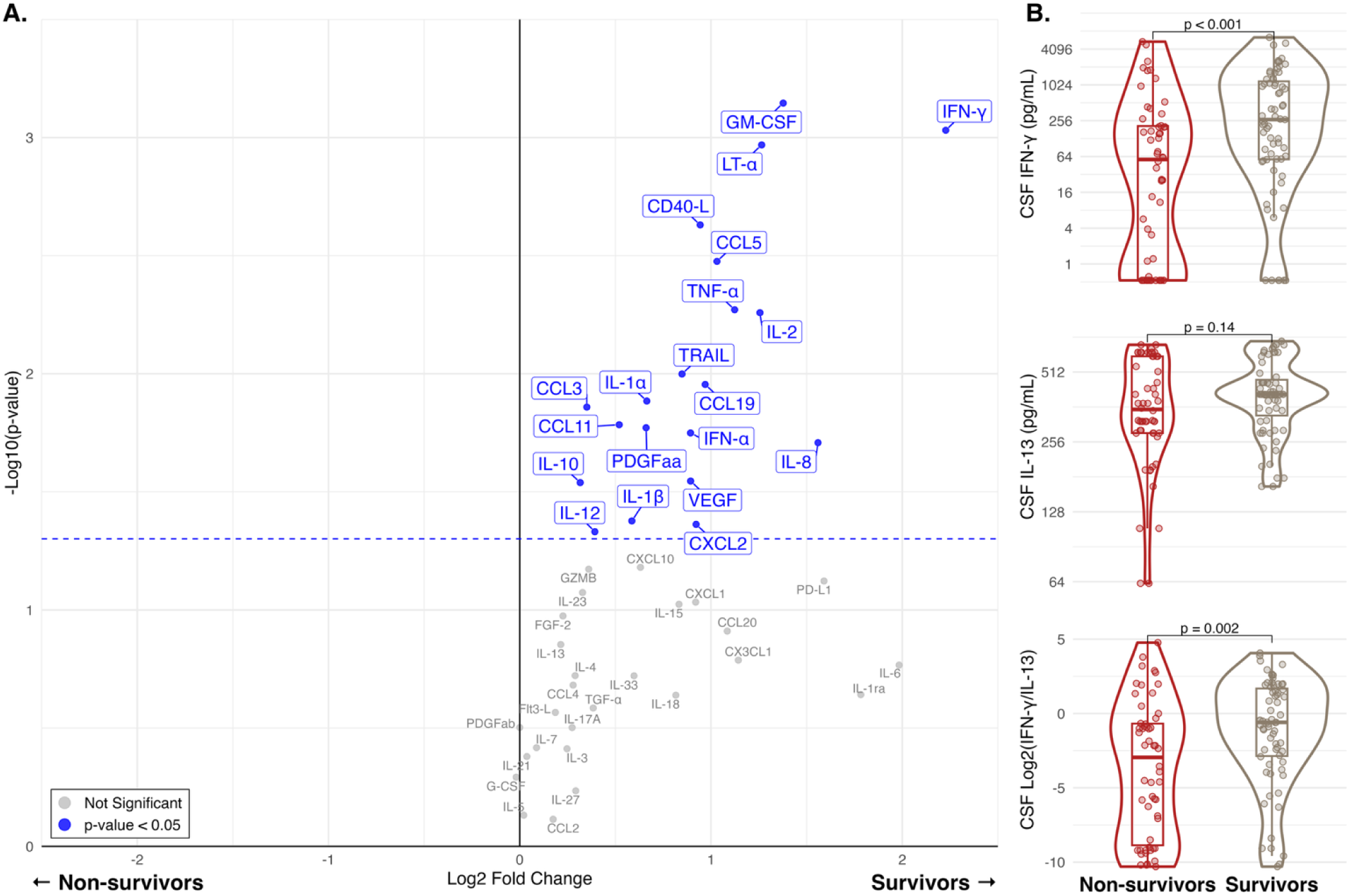
Comparison of baseline CSF immune mediator levels between TBM survivors and non-survivors living with HIV. (A) Volcano plot of differentially expressed analytes across survival groups. The x-axis shows log_2_ fold-change and the y-axis shows –log_10_(p-value), each point represents one analyte (47 total). Analytes upregulated in survivors appear to the right of the vertical black line (x = 0), and those upregulated in non-survivors to the left. The horizontal blue dotted line denotes a p-value threshold of 0.05; analytes under this threshold are labeled in blue. (B) Violin plots showing the distribution of CSF concentrations for selected analytes and cytokine ratios in survivors (right; beige) and non-survivors (left; red). Boxes indicate the interquartile range, horizontal lines mark the median, and whiskers extend 1.5 times the interquartile range from the 25th and 75th percentiles. Individual data points are overlaid as dots. Group comparisons were performed using the Wilcoxon rank-sum test, with p-values shown above each plot. Censored participants excluded (n= 121). Abbreviations: CSF, cerebrospinal fluid; IFN, interferon; IL, interleukin.

**Table 2.**
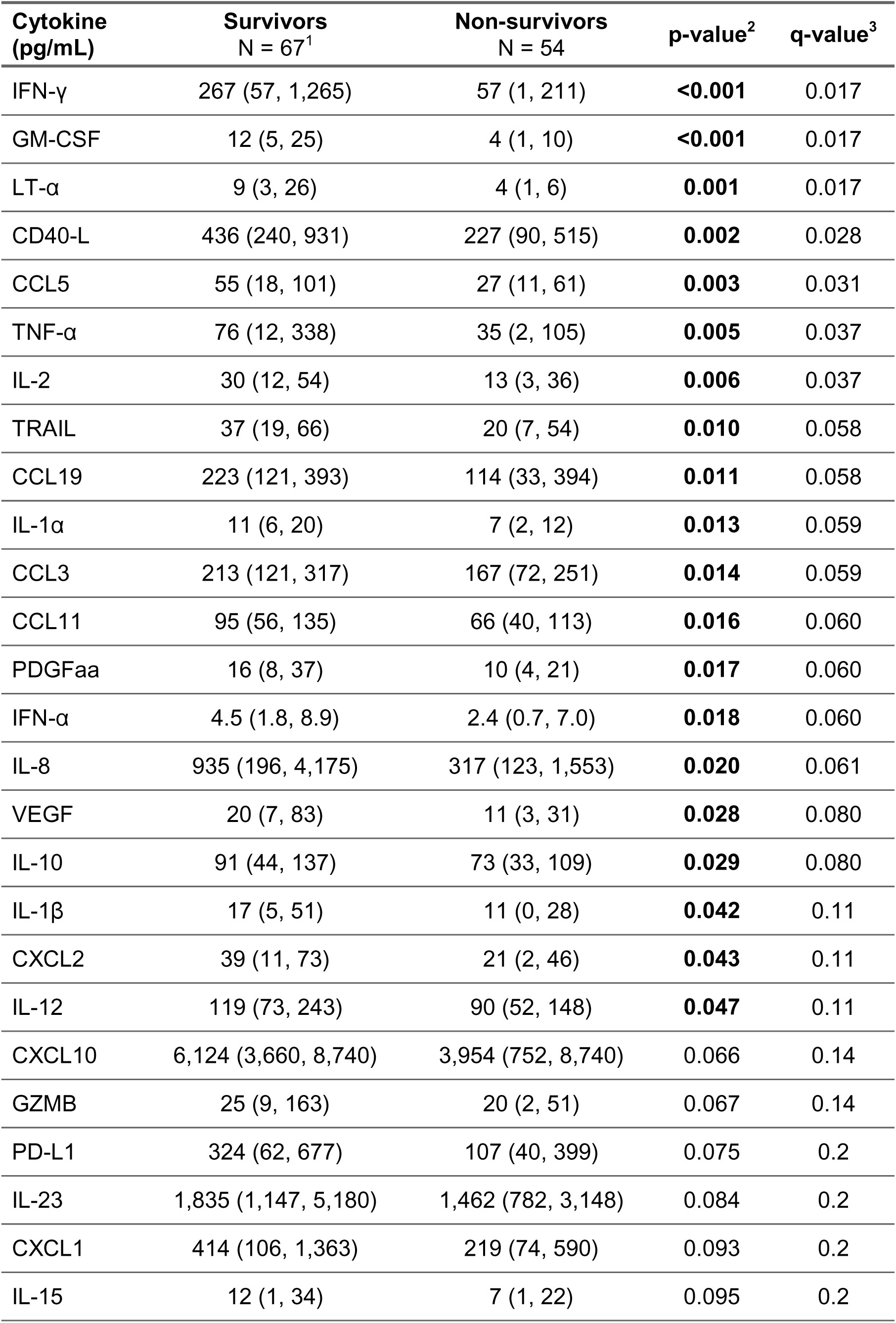
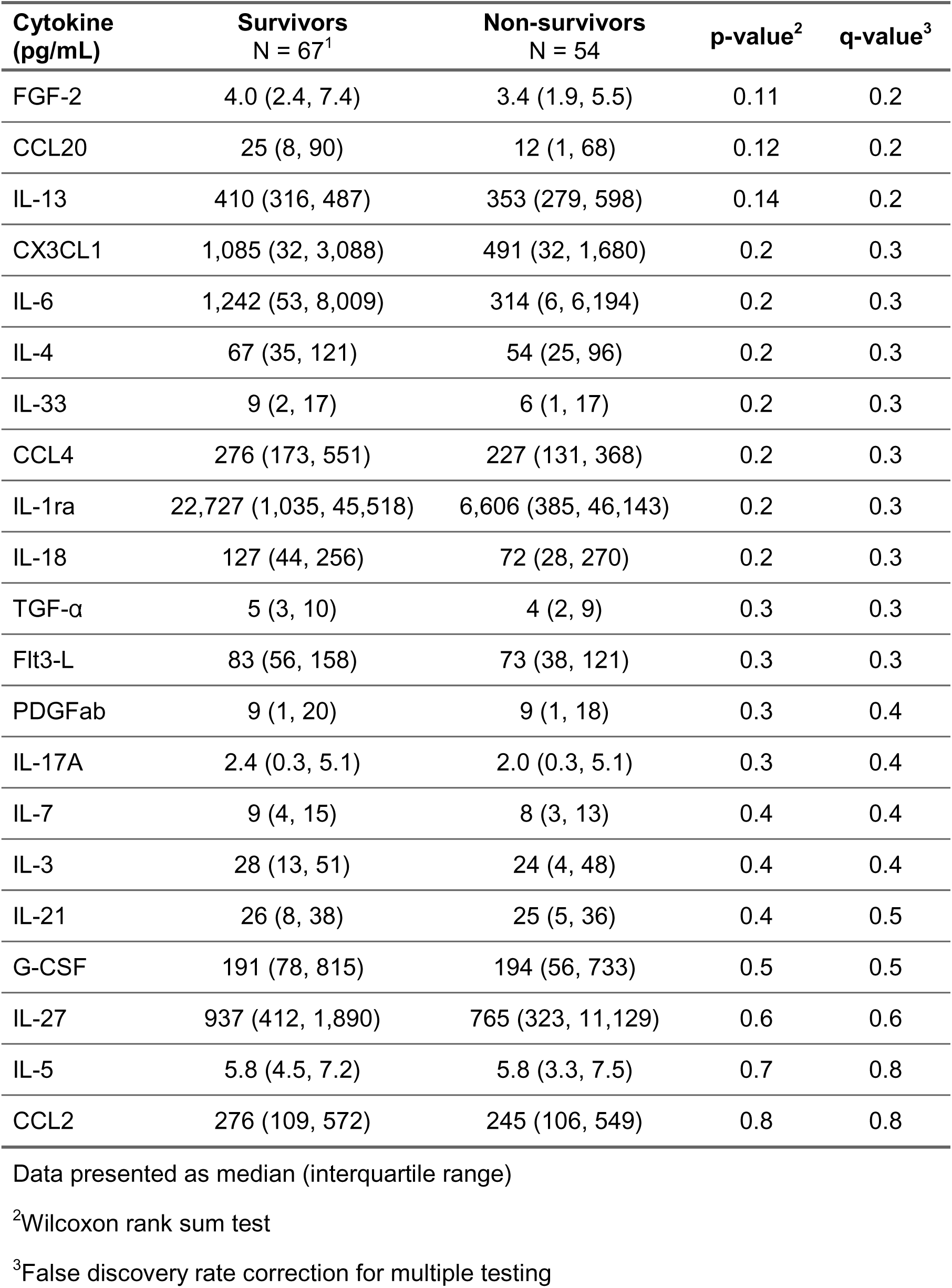
Baseline CSF Cytokine Concentration Differences Between 90-day Survivors and Non-survivors with HIV-associated TBM.

### Association between mortality and CSF inflammation is dose-dependent and cytokine-specific

To further test the hypothesis that deficient local Th1 immunity drives mortality in HIV-associated TBM, we investigated dose-dependent effects of individual cytokines on 90-day survival. We compared survival across tertiles (bottom, middle, top) based on CSF concentration of each cytokine in the full cohort of 149 individuals (**Supplemental Data**). For IFN-γ, 90-day mortality was 50% (25/50) for the bottom tertile, 41% (20/49) for the middle, and 18% (9/50) for the top tertile (**Fig. 2A**). A similar pattern was observed for other Th1 cytokines including TNF-α, GM-CSF, and IL-2 (**Fig. S2**). In contrast, for IL-13, individuals with intermediate CSF expression (middle tertile), had the lowest 90-day mortality (22% (10/46)), compared to those in the bottom or top tertiles (**Fig. 2B**). These data further support the key role for local Th1 immune responses in protection against HIV-associated TBM. They also suggest that mortality may be driven by either insufficient immunity, or inappropriate non-protective (*i.e*. Th2) responses.

**Figure 2.**
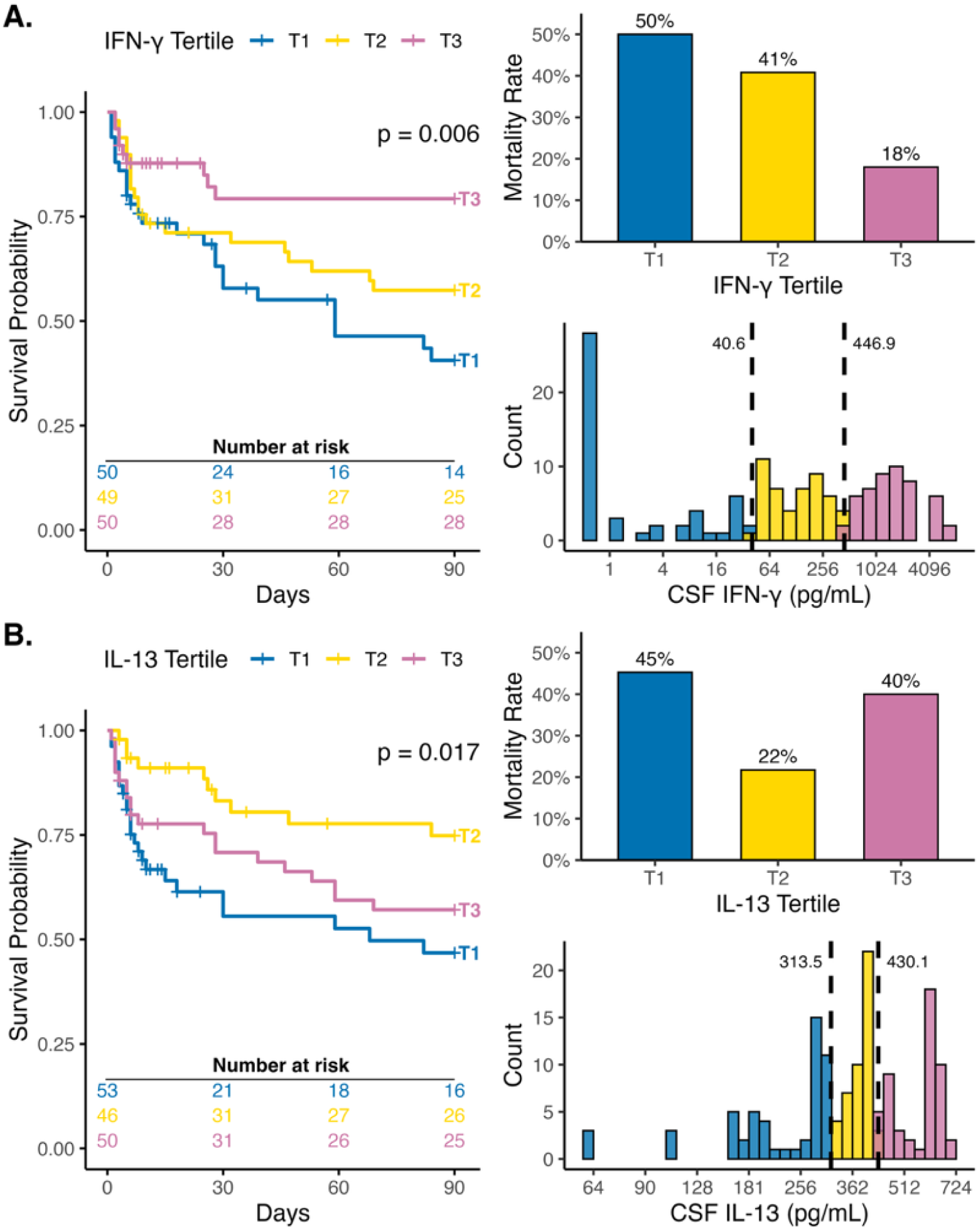
90-day survival by CSF IFN-γ and IL-13 tertiles in HIV-related TB meningitis. Kaplan–Meier survival analyses of participants (n = 149) stratified by tertiles of CSF IFN-γ (A) and CSF IL-13 (B). Log-rank p-values are shown, with number at risk listed below each plot. Vertical tick marks indicate censored participants. Each panel also includes a bar plot showing 90-day mortality rate by tertile, and a histogram of cytokine concentrations in pg/mL) with tertile ranges indicated by vertical black dashed lines. Tertiles are colored as T1 (blue), T2 (yellow), and T3 (pink). Abbreviations: CSF, cerebrospinal fluid; IFN-γ, interferon-γ; IL-13, interleukin-13.

### CSF Th1 cytokine expression is tightly coordinated

Non-survivors exhibited both severe peripheral CD4 T cell depletion and marked suppression of multiple CSF cytokines, prompting investigation into whether these processes are mechanistically linked. To address this, we calculated pairwise correlations among the 20 mortality-associated CSF cytokines and their relationships to peripheral CD4 and CSF WBC counts (**Fig. 3A**). This analysis showed that IFN-γ, TNFα, and IL-2 were strongly correlated (e.g., IFN-γ:TNF-α *r* = 0.88; IFN-γ:IL-2 *r* = 0.87), consistent with their function as co-effector cytokines within the Th1 immune response (**Fig. 3B**). IFN-γ was also positively correlated with other canonical inflammatory mediators, including IL-1α (*r*= 0.79), but not with IL-13 (**Fig. 3B**). This absence of immune signaling was accompanied by diminished CNS cellular immunity, reflected in the significant correlation between multiple cytokine levels and CSF WBC counts (**Fig. 3A**). Notably, IFN-γ was the only mortality-associated cytokine associated with peripheral CD4 counts. These analyses reveal the linkage between protective systemic CD4 T cell immunity and the local production of Th1 cytokines in the CSF in HIV-associated TBM.

**Figure 3.**
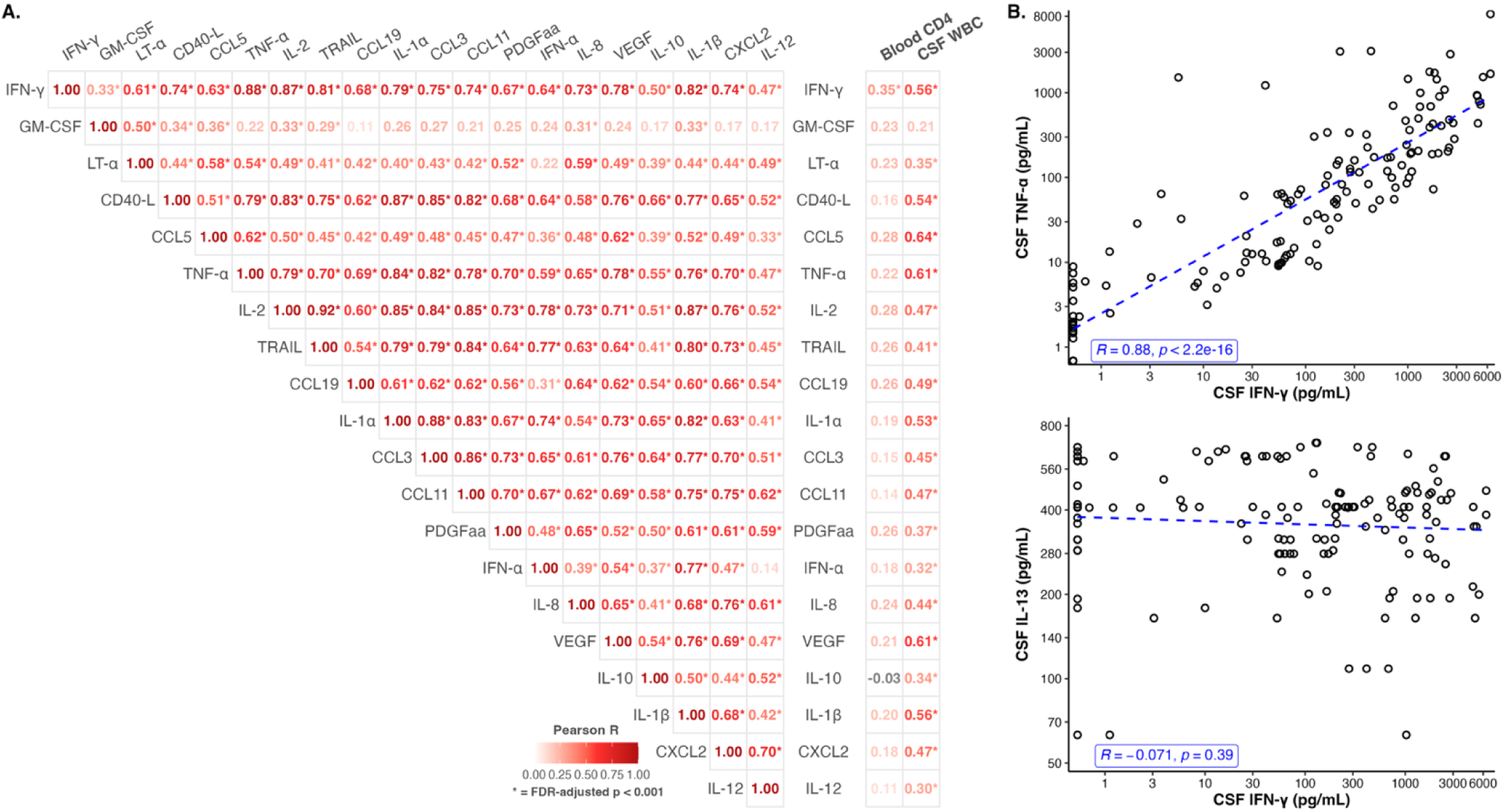
Correlations between mortality-associated CSF immune mediators, CSF WBC counts, and blood CD4 T cell counts. (A) Pairwise Pearson correlation matrix of the 14 immune mediators significantly different between survivors and non-survivors, together with CSF WBC counts and blood CD4 T cell counts. Pearson R correlation coefficients are shown and colored by their value, with darker red indicating stronger correlation. Asterisks (*) mark significant correlations with FDR-adjusted p value < 0.001. (B) Scatterplots showing relationships between the CSF concentrations (pg/mL) of IFN-γ and TNF-α or IL-13. Each point represents an individual patient (n = 149). Blue dashed lines indicate linear regression fit. Pearson R coefficients and p-values are shown within each. Abbreviations: CSF, cerebrospinal fluid; WBC, white blood cell; IFN-γ, interferon-γ; IL-13, interleukin-13; TNF-α, tumor necrosis factor-α.

### Cellular immunity and CSF cytokine production interact to provide protection

To further characterize how CNS inflammation relates to systemic and local immune cell function, we compared IFN-γ concentrations with peripheral CD4 counts and CSF WBC counts. Break points for CD4 (≤100 cells/μL) and IFN-γ (≤214 pg/mL) were derived from ROC curve analysis (**Fig. S2**) using Youden’s index, and CSF WBC was dichotomized at the clinical pleocytosis threshold (>5 cells/μL). We found that the lowest mortality (12% (3/25)) occurred in individuals with both high CD4 count and CSF IFN-γ level (**Fig. 4A**). Conversely, the highest mortality (63% (24/38)) was observed in those with low levels of both parameters. Of note, even among those with peripheral CD4 depletion ≤100 cells/μL, participants who mounted a high CSF IFN-γ had lower mortality (27% (4/15) vs. 63% (24/38)). Kaplan-Meier survival analysis including the full cohort of 149 individuals confirmed these survival differences, with the higher risk for death in people with CD4 T cell counts ≤100 cells/μL (HR 3.30 (1.55, 6.99), **Fig 4B**). Among those with CD4 T cell counts ≤100 cells/μL, the presence of a CSF IFN-γ response was protective (HR 0.32 (0.11, 0.91), **Fig 4C**) associated with survival comparable to those with relatively preserved blood CD4 T cells (**Fig 4D**).

**Figure 4.**
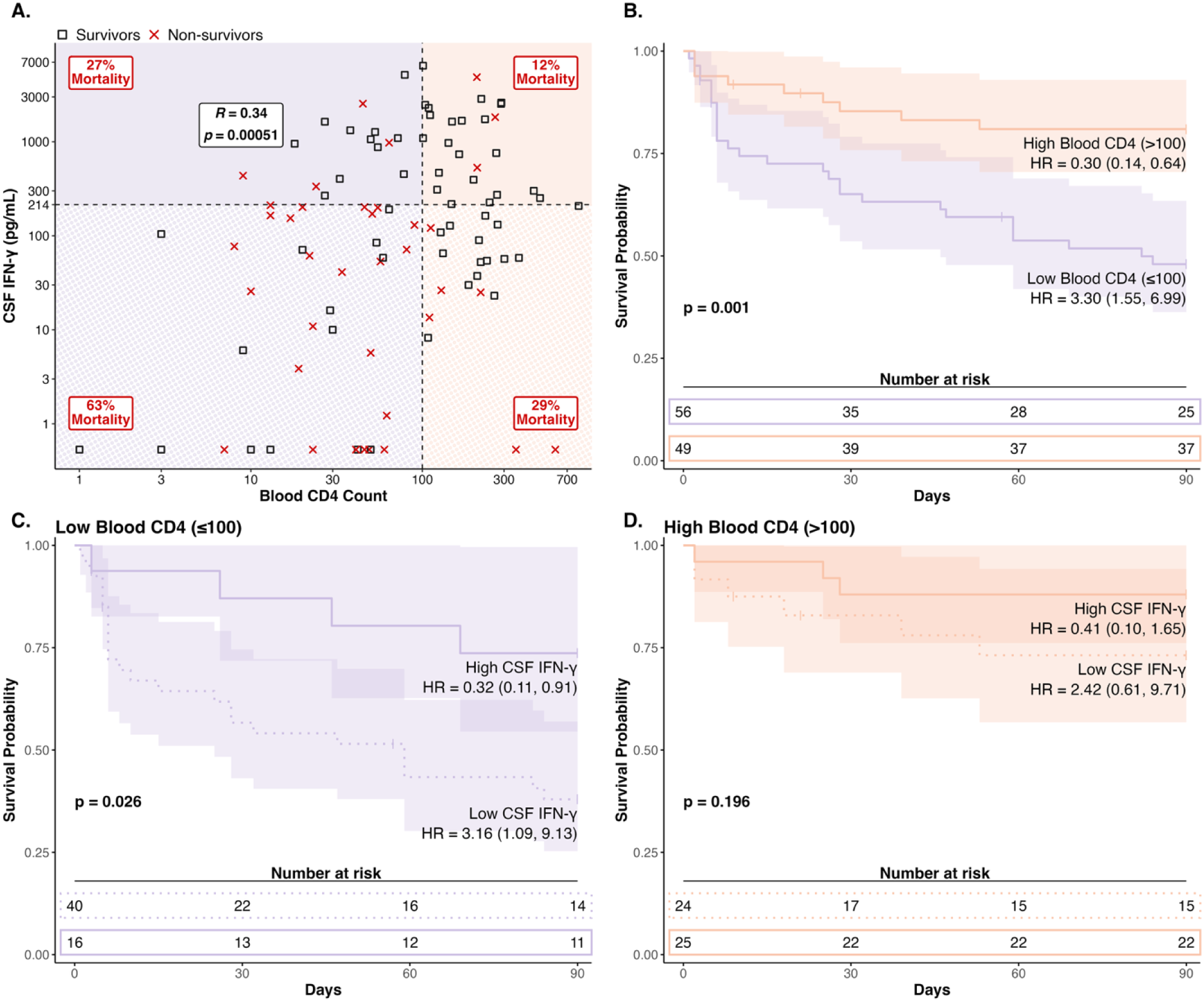
Interaction of blood CD4 T cell counts and CSF IFN-γ concentration on 90-day survival. (A) Scatterplot showing the linear relationship and Pearson *r* between blood CD4 T cell counts and CSF IFN-γ concentrations (pg/mL). ROC-derived cutoff values (100 cells/μL for CD4; 214 pg/mL for IFN-γ) are shown as dotted black lines. Mortality rates are ndicated in each quadrant. The color and overlay patterns in A correspond to the survival curve groupings in B–D (e.g., high CD4/high IFN-γ = solid orange; high CD4/low IFN-γ = dotted orange). (B) Kaplan–Meier survival curves comparing participants with low (purple) vs. high (orange) blood CD4 T cell counts. These groups were further stratified by CSF IFN-γ evel (low, dotted; high, solid) within the (C) low CD4 group and (D) high CD4 group. HRs from Cox proportional hazards models, number at risk, and censored participants (ticks) are shown for B–D. All are limited to participants with available CD4 data (n = 105), while A further excludes censored participants (n = 99). Abbreviations: CSF, cerebrospinal fluid; IFN, nterferon; ROC, receiver operating characteristic; HR, hazard ratio.

In contrast, people with CSF pleocytosis (>5 cells/μL) had lower mortality only when accompanied by elevated IFN−γ (24% (10/42) vs. 59% (19/32), **Fig. 5A-D**). When CSF WBC was ≤5 cells/μL, mortality remained high (∼60%) regardless of CSF IFN−γ concentration. We next examined whether this IFN-γ-dependent survival advantage was associated with specific CSF leukocyte subsets. CSF IFN-γ concentrations correlated strongly with absolute lymphocyte counts (r = 0.45, p=.002, **Fig. S3A**) but showed no relationship with neutrophil counts (r = –0.05, p=.78, **Fig. S3B**), Collectively, these findings argue that mortality in HIV-associated TBM is driven by the failure to mount a protective, Th1-driven inflammatory response within the CNS.

**Figure 5.**
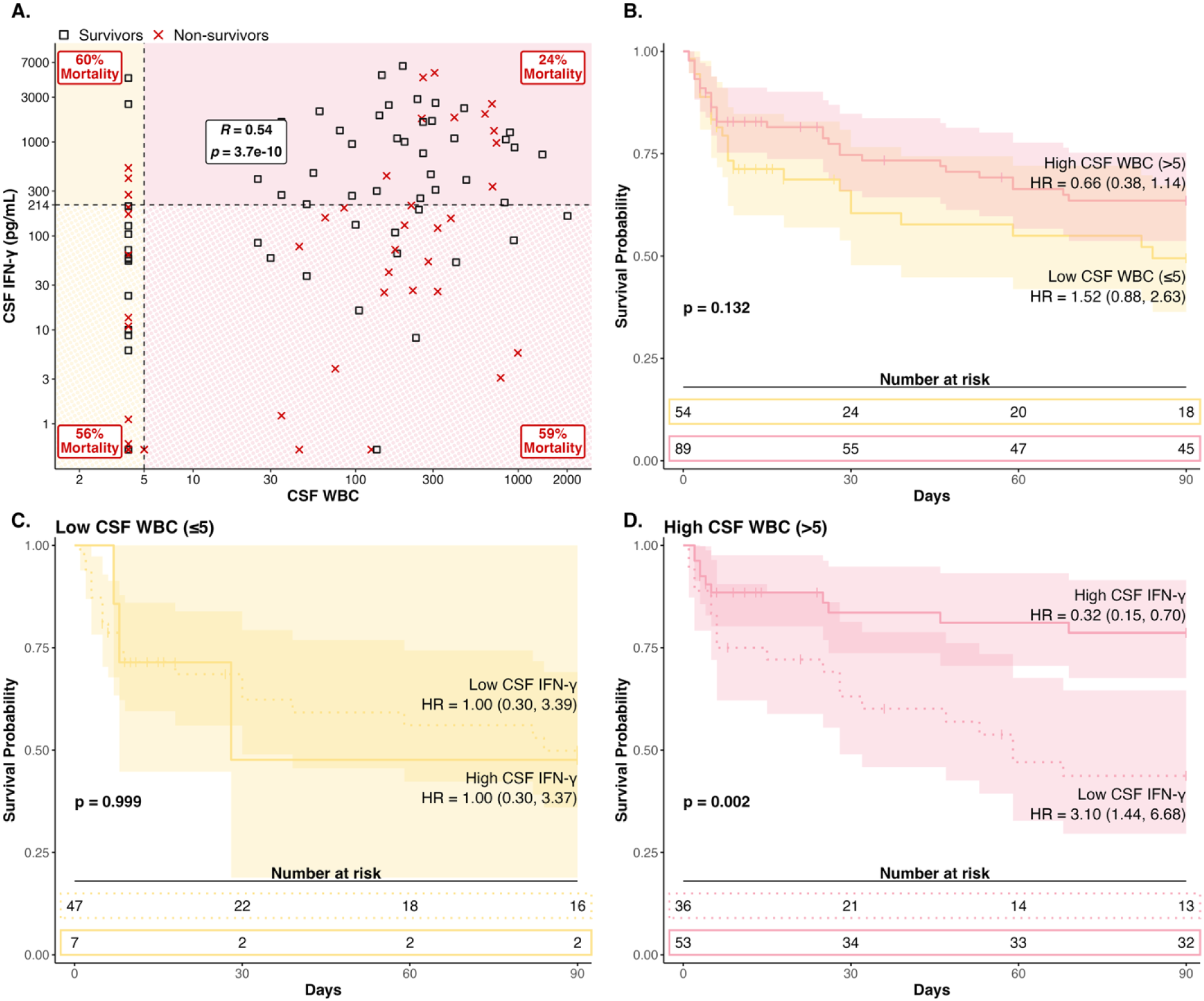
Interaction of CSF WBC counts and CSF IFN-γ concentration on 90-day survival. (A) Scatterplot showing the linear relationship and Pearson correlation between CSF white cell counts and CSF IFN-γ concentrations in pg/mL. The clinical pleocytosis cutoff (5 cells/μL for WBC) and ROC-derived cutoff for IFN-γ (214 pg/mL) are shown as dotted black lines. Mortality rates are indicated within each quadrant. The color and overlay patterns in panel A correspond to the survival curve groupings in panels B–D (e.g., high WBC/high IFN-γ = solid pink; high WBC/low IFN-γ = dotted pink). (B) Kaplan–Meier survival curves comparing participants with low (yellow) versus high (pink) CSF WBC counts. These groups were then further stratified by CSF IFN-γ level (low, dotted; high, solid) within the (C) low WBC group and (D) high WBC group. Hazard ratios from Cox proportional hazards models, number at risk, and censored participants (tick marks) are shown for all survival panels (B–D). All panels are limited to participants with available CSF WBC data (n = 143), which were used for survival analyses (B–D), while panel A further excludes censored participants (n = 115). Abbreviations: CSF, cerebrospinal fluid; IFN, interferon; WBC, white blood cells; ROC, receiver operating characteristic; HR, hazard ratio.

## Discussion

TBM is a persistently high and disproportionate cause of TB mortality [3,4,25]. Outcomes remain particularly poor among those with HIV, even in the modern era of antiretroviral therapy [3]. Therefore, there is an urgent clinical need to improve diagnosis, risk stratification, and treatment of TBM. Identifying those at greatest risk and elucidating the mechanisms underlying their poor outcomes could guide the development of tailored therapeutics that save lives. Our study of CSF inflammatory mediators among Ugandan adults with HIV-associated TBM is the broadest characterization of the CNS immune signaling landscape in this vulnerable population to date, providing new insights into mechanisms of protection. Although non-survivors in our study presented with more severe disease, few other standard clinical characteristics distinguished this high-risk group from those who survived to 90 days. Notably, while individuals with TBM, without HIV, benefit from adjunctive corticosteroid treatment, those with HIV co-infection do not universally benefit [22,23], raising questions about the role of inflammation in this vulnerable population. Our study suggests that the greatest risk of mortality in people with HIV-associated TBM is among those with the worst immune function. These findings illustrate the need for new biomarkers of risk, better definition of distinct disease phenotypes, and a clearer understanding of how immune protection against this disease is mediated.

In people with HIV-associated TBM, we found that mortality was strongly associated with worse systemic CD4 T cell depletion. In this cohort, utilization of HIV therapy was similar between survival groups, but blood CD4 counts differed substantially, with non-survivors being more likely to have severe depletion of <100 cells/μl. One limitation of our study is that, while antiretroviral therapy duration was recorded, viral suppression and adherence data were not [26]. Understanding the HIV disease mechanisms that underlie worse immunosuppression in this cohort is an important goal for future work. These data also underscore the utility of baseline CD4 T cell count testing for risk stratification in HIV-associated TBM and, likely, for TB in general and other opportunistic infections. In the era of universal HIV test-and-treat, CD4 T cell count testing is performed less often in many settings but these data suggest that there is a role for point of care CD4 tests in persons with suspected TBM.

We found that local Th1 immunity in the CSF is highly coordinated and critically important for control of CNS *Mtb* in the context of HIV. Th1 cytokines like IFN-γ, TNF-α, and IL-2 were more abundant in the CSF of survivors. As expected, these Th1 cytokines were highly correlated with each other, but not with Th2 cytokines such as IL-13. IFN-γ and TNF-α are pro-inflammatory cytokines that are essential for protection against *Mtb* infection. However, they can play a dual role: while insufficient signaling impairs mycobacterial control, excessive production of either can contribute to immunopathology [16,17]. In this cohort of patients with HIV, we found little evidence to suggest that hyper-production of either cytokine contributed to worse outcome. However, one potential explanation for the differentially greater expression of Th1 cytokines among survivors could be that the adjunctive corticosteroid treatment received by all individuals in this cohort only benefited people with abundant CSF inflammation. When survival was examined by stratifying each baseline CSF cytokine into tertiles, we observed that only one—IL-13—was significantly associated with better outcome at intermediate levels. Higher mortality of people with the highest CSF IL-13 levels suggests that Th2-skewed local immunity may be detrimental to protection against TB. Concordantly, survivors had a significantly higher ratio of IFN-γ:IL-13 than non-survivors, consistent with the protective role of Th1 immunity. IL-13, which contributes to defense against parasites, is one of several cytokines that are relatively abundant in the CSF of people with TBM compared to other CNS infections [27]. Although in a prior Vietnamese study people with concurrent *Strongyloides stercoralis* infection had fewer neurological complications from TBM, this difference was not associated with higher CSF concentrations of Th2 cytokines, including IL-13 [28]. Future trials investigating the potential benefit of anti-inflammatory host-directed therapies in TBM should consider strategies to target these therapies towards cytokines with a clear association to risk, and in individuals with evidence of excessive or inappropriate CSF inflammation. A one-size fits all approach will not be optimal for all persons.

Our study identifies an important link between systemic immunosuppression and local CNS inflammation. Due to logistical complexities, few studies of humans with TB have clearly linked measures of immune function in the peripheral blood to those at the site of infection. In contrast to pulmonary TB, samples from infected tissues are routinely obtained in TBM, providing a unique opportunity to gain mechanistic insight into human TB immunology. Consistent with the classical model of Th1 immune protection in TB, we found that blood CD4 count was significantly correlated with the CSF concentration of IFN-γ. However, no other CSF cytokine was strongly concordant with blood CD4 T cell count, arguing that distinct immunologic mechanisms may occur at different tissue sites. While non-survivors had relatively lower levels of both blood CD4 count and CSF IFN-γ compared to survivors, there was also an interaction between the two parameters. Individuals with concordantly low levels of both had the highest risk of 90-day mortality. Importantly, even among people with severe CD4 T cell depletion ≤100 cells/μL, survival was significantly better among those who mounted a CSF IFN-γ response. These data indicate the potential utility of CSF IFN-γ as a biomarker of protection, particularly when combined with blood CD4 T cell count. Prospective evaluation of CSF IFN-γ could establish its prognostic value and enable immunophenotyping to guide tailored therapies, distinguishing patients who may benefit from corticosteroids from those who may require immune-boosting strategies.

In summary, this study reveals the clinical value of defining CSF inflammation in CNS TB, illustrates the protective importance of mounting an appropriate local immune response, identifies immunologic endotypes that may underlie the heterogenous disease outcomes in HIV-associated TBM, and suggests groups of high-risk individuals who may not benefit from adjunctive anti-inflammatory treatment with corticosteroids. These findings provide strong justification for the continued, and more comprehensive immunologic characterization of CSF and other CNS tissue sites in people with TBM, particularly for those with HIV co-infection.

## Data Availability

All data produced in the present study are available upon reasonable request to the authors

## Collaborators

Derrick Kasozi, Morris K Rutakingirwa, Laura Nsangi, Jayne Ellis, Isaac Turyasingura, Sylvia Namanda, Florence Kigundu, Jane Francis Ndyetukira, Carol Namujju, Alice Namudde, Grace Menya, Shifah Nabbaale, Cynthia Ahimbisibwe, Alisat Sadiq, Kiiza Tadeo Kandole, Wilber Bakar, Dorothy Babirye, Tonny Luggya, Andrew Akampulira, Richard Kwizera, Michael Okiror, John Kisembo, Rhona Muyise, Mayaja Hamuza, David Ssebunya, Suzan Mulwana, Eunice Okiror, David Tenywa

## Author contributions

**Anna L. Wilt**: Methodology, Software, Formal analysis, Investigation, Data Curation, Writing − Original Draft, Visualization; **David B. Meya:** Conceptualization, Resources, Writing, Project administration, Funding acquisition; **Fiona V. Cresswell**: Investigation, Reviewing draft; **Abduljewad Wele:** Data Curation, Formal analysis, Methodology; **Mable Kabahubya**: Investigation; **Enock Kagimu:** Investigation**; Jane Gakuru**: Investigation; **Timothy Mugabi:** Investigation; **Sarah Kimuda:** Investigation; **Suzan Namombwe:** Investigation; **Asmus Tukundane:** Investigation**; Elizabeth C. Okafor**: Methodology, Investigation; **Biyue Dai:** Formal analysis, Methodology, Reviewing draft; **Nicole Engen:** Formal analysis, Methodology**; Nathan C. Bahr:** Conceptualization, Methodology, Resources, Writing, Supervision, Project administration, Funding acquisition; **David R. Boulware**: Conceptualization, Methodology, Resources, Writing, Supervision, Project administration, Funding acquisition; **Tyler D. Bold**: Conceptualization, Methodology, Resources, Writing, Supervision, Project administration, Writing - Original Draft, Funding acquisition.

