## Supplemental Figures for "CSF Hypo-Inflammation Drives Mortality in HIV-Associated Tuberculous Meningitis"

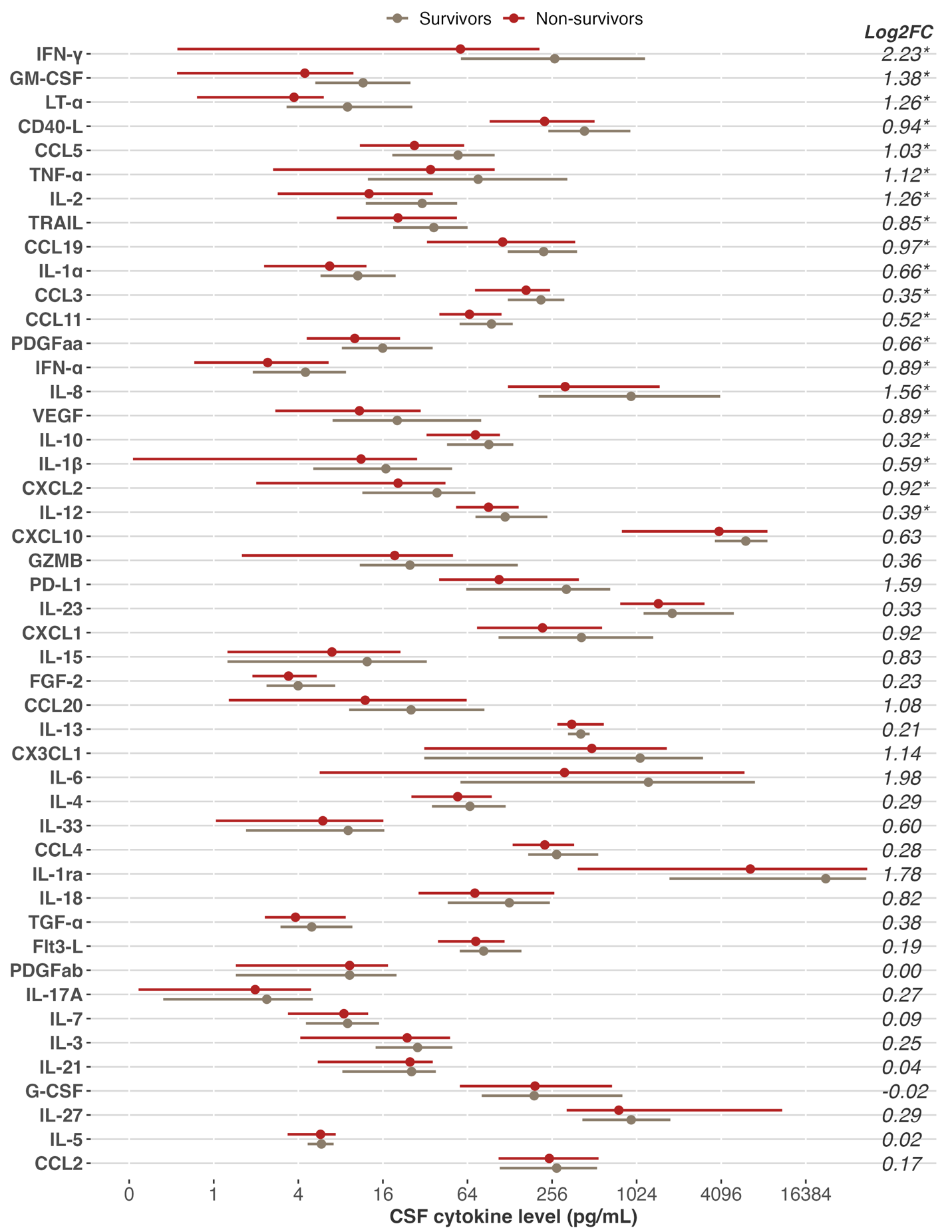

**Supplementary Figure 1. Median CSF immune mediator concentrations by survival group.** Median (dot) and interquartile range (line) of each of the 47 CSF analytes are shown for survivors (bottom; beige) and non-survivors (top; red). X-axis values are CSF concentrations (pg/mL). Log_2_ fold change (survivors – non-survivors) is displayed on the right side. Analytes are ordered by p-value from Wilcoxon rank-sum tests, with asterisks (*) indicating p < 0.05. Eight right-hand censored participants were excluded (n = 121). Abbreviations: CSF, cerebrospinal fluid; FC, fold change.

**Supplementary Figure 2. ROC analysis to determine optimal cutoffs for mortality prediction.** ROC curves are shown for (A) CSF IFN-γ concentrations (pg/mL) and (B) blood CD4 T cell counts (cells/μL), with specificity (x-axis) plotted against sensitivity (y-axis). Optimal cutoffs were defined using Youden’s index; red dashed lines indicate the sensitivity and specificity at the selected threshold, with values listed in the inset box. The AUC is shown within each panel. Censored participants were excluded (n = 121). Abbreviations: CSF, cerebrospinal fluid; IFN-γ, interferon-γ; ROC, receiver operating characteristic; AUC, area under the curve.

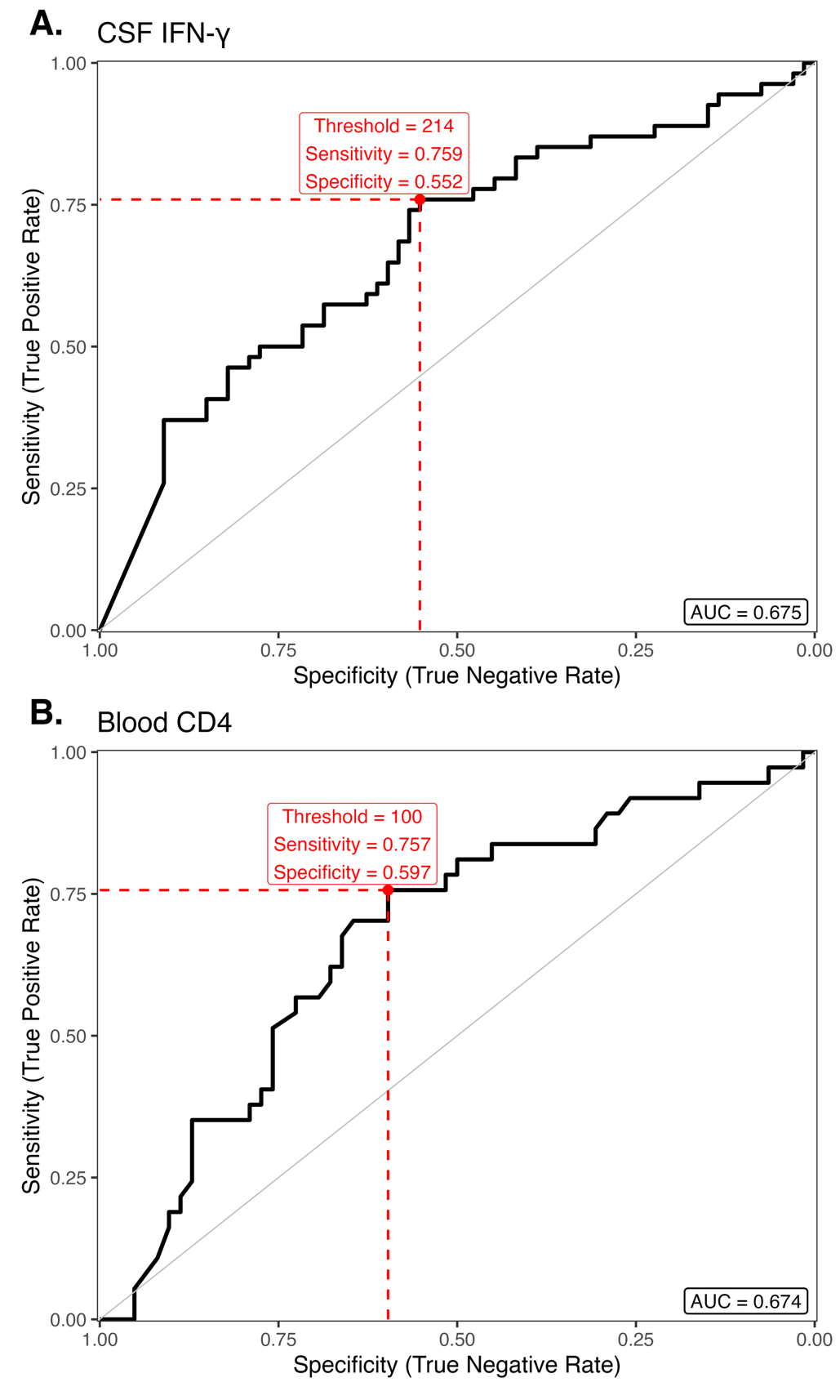

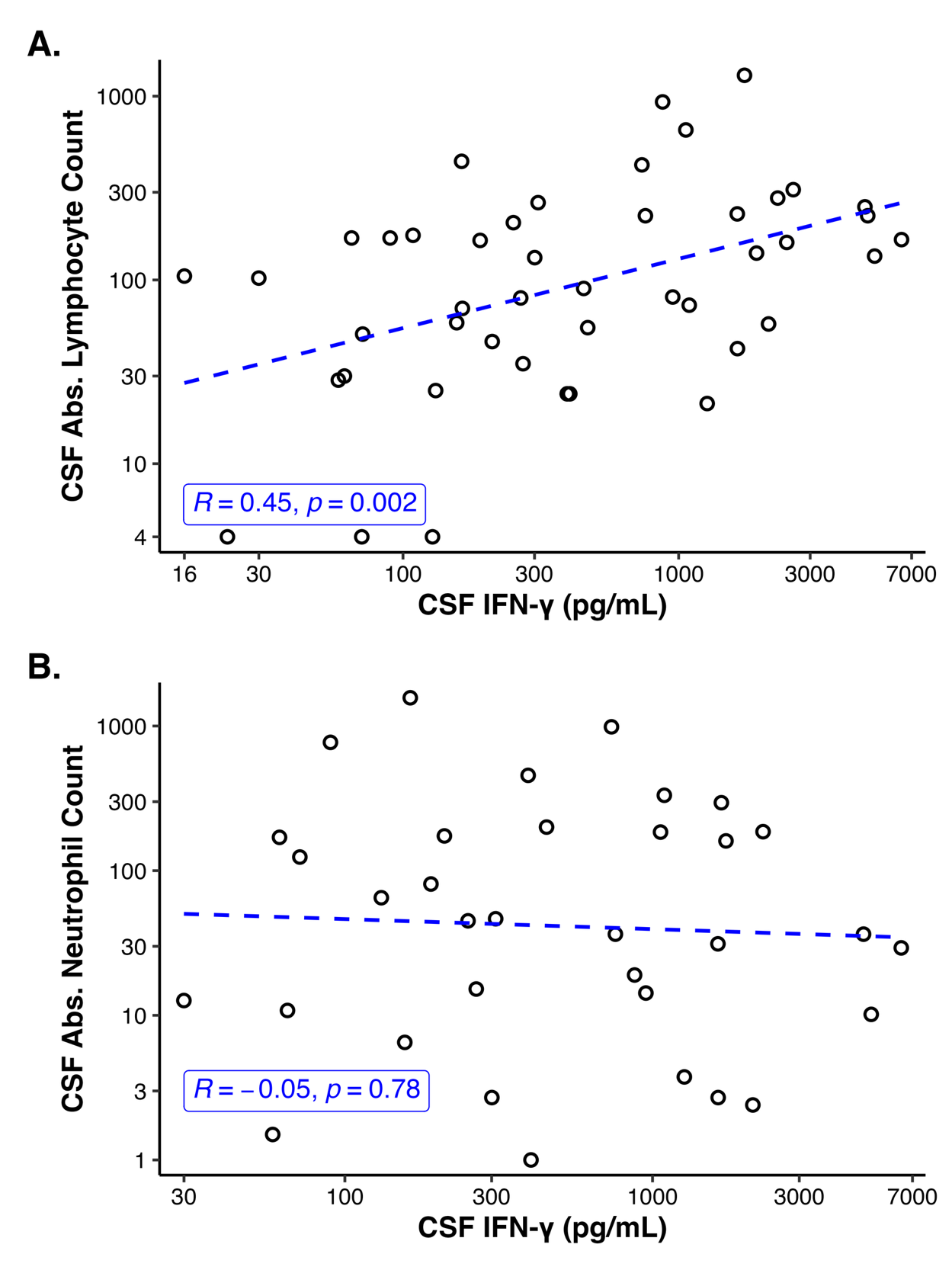

**Supplementary Figure 3. Correlations between CSF IFN-γ concentrations and leukocyte subsets.** Scatterplots display the relationships between CSF IFN-γ (pg/mL) and absolute CSF (A) lymphocyte or (B) neutrophil counts (cells/μL). Each point represents an individual patient with counts > 0 (A: n = 45; B: n = 34). Blue dashed lines indicate linear regression fit. Pearson R correlation coefficients and p-values are shown within each panel. Abbreviations: CSF, cerebrospinal fluid; IFN-γ, interferon-γ; Abs., absolute.
