## Supplemental Data for "CSF Hypo-Inflammation Drives Mortality in HIV-Associated Tuberculous Meningitis"

### Log-Rank Summary (Tertiles)

| Cytokine | Log-rank p | Significant |
|---|---|---|
| LT-α | 0.004 | Yes |
| GM-CSF | 0.004 | Yes |
| TNF-α | 0.017 | Yes |
| CCL5 | 0.022 | Yes |
| CD40-L | 0.032 | Yes |
| CXCL2 | 0.042 | Yes |
| IL-2 | 0.043 | Yes |
| PD-L1 | 0.052 | |
| TGF-α | 0.054 | |
| CCL19 | 0.056 | |
| CXCL1 | 0.064 | |
| CCL3 | 0.069 | |
| TRAIL | 0.074 | |
| IL-1β | 0.090 | |
| GZMB | 0.091 | |
| CXCL10 | 0.091 | |
| PDGFaa | 0.092 | |
| CCL20 | 0.093 | |
| IL-1α | 0.113 | |
| FGF-2 | 0.113 | |
| IL-10 | 0.122 | |
| PDGFab | 0.129 | |
| IFN-α | 0.129 | |
| Flt3-L | 0.166 | |
| IL-12 | 0.172 | |
| IL-33 | 0.199 | |
| CCL11 | 0.226 | |
| IL-23 | 0.240 | |
| IL-3 | 0.265 | |
| IL-15 | 0.270 | |
| VEGF | 0.283 | |
| IL-18 | 0.288 | |
| IL-27 | 0.321 | |
| IL-8 | 0.321 | |
| IL-17A | 0.375 | |
| CCL4 | 0.391 | |
| IL-4 | 0.410 | |
| IL-6 | 0.474 | |
| IL-7 | 0.590 | |
| IL-21 | 0.716 | |
| IL-1ra | 0.730 | |
| G-CSF | 0.809 | |
| IL-5 | 0.887 | |
| CCL2 | 0.967 | |

# LT-α

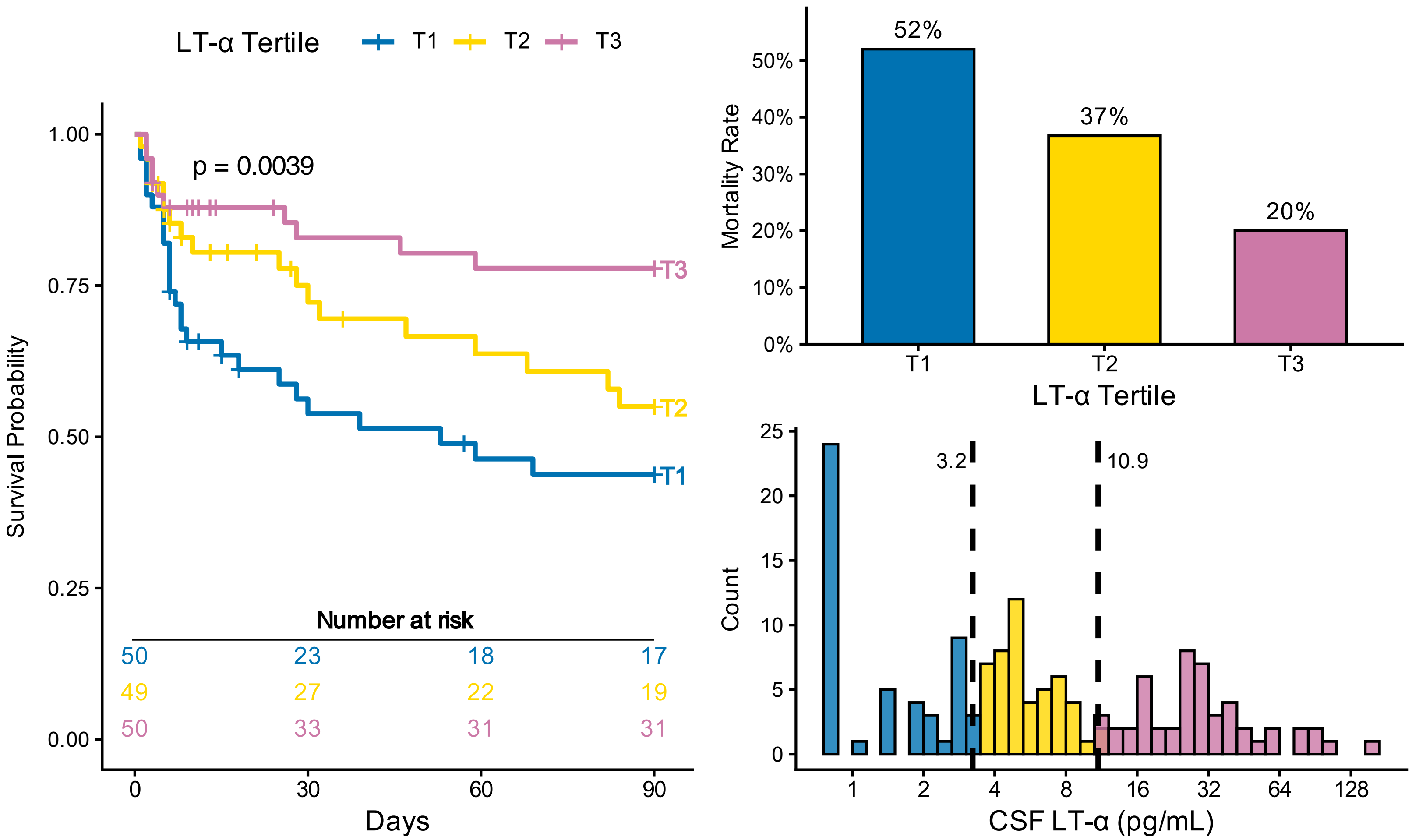

LT-α Tertile T1 T2 T3
p = 0.0039
Survival Probability
1.00
0.75
0.50
0.25
0.00
T1
T2
T3
Number at risk
50 23 18 17
49 27 22 19
50 33 31 31
0 30 60 90
Days
52%
37%
20%
Mortality Rate
50%
40%
30%
20%
10%
0%
T1 T2 T3
LT-α Tertile
Count
25
20
15
10
5
0
3.2
10.9
1 2 4 8 16 32 64 128
CSF LT-α (pg/mL)


### GM-CSF

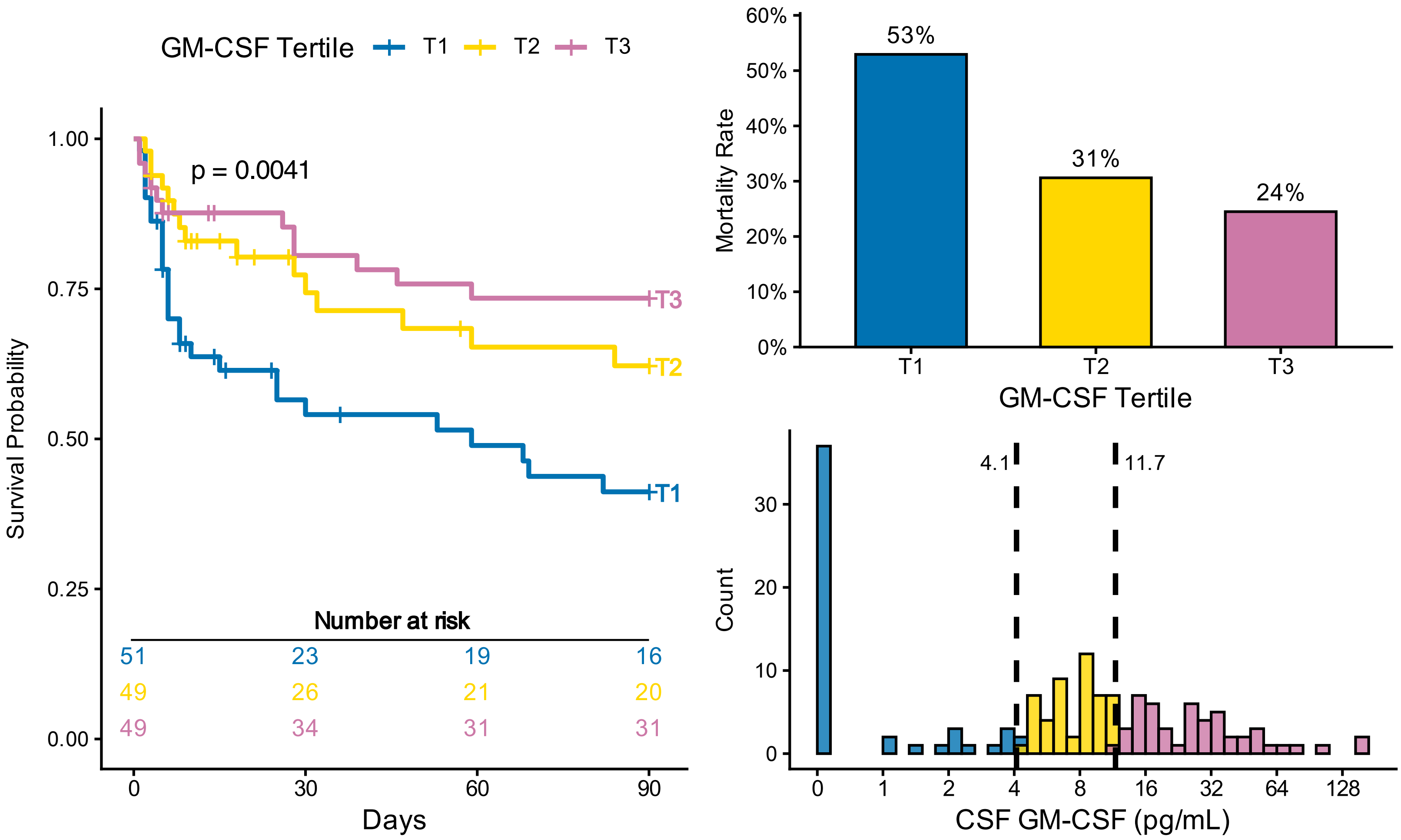

GM-CSF Tertile T1 T2 T3
p = 0.0041
Survival Probability
1.00
0.75
0.50
0.25
0.00
T1
T2
T3
Number at risk
51 23 19 16
49 26 21 20
49 34 31 31
0 30 60 90
Days
53%
31%
24%
Mortality Rate
60%
50%
40%
30%
20%
10%
0%
T1 T2 T3
GM-CSF Tertile
Count
30
20
10
0
4.1
11.7
0 1 2 4 8 16 32 64 128
CSF GM-CSF (pg/mL)

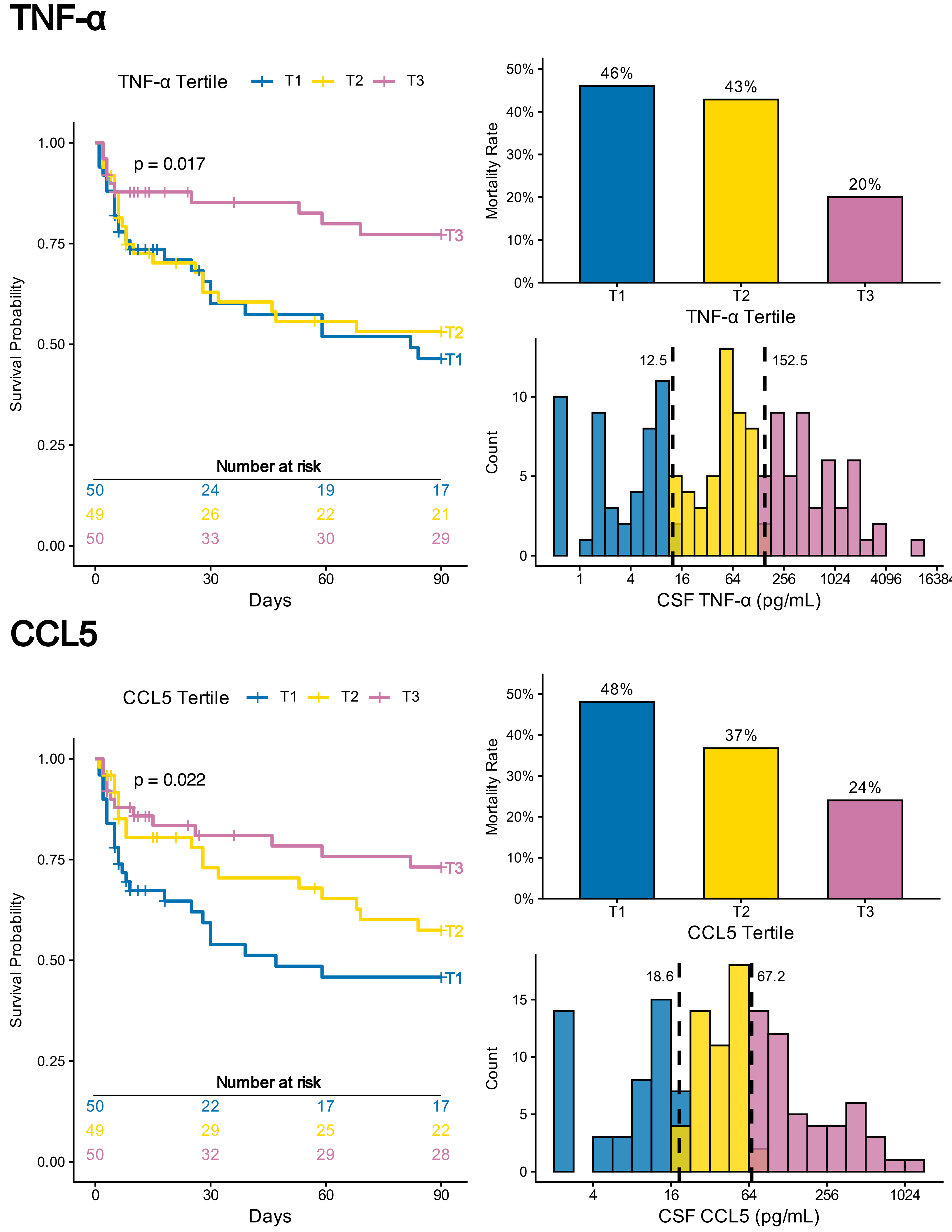

TNF-α
TNF-α Tertile
T1
T2
T3
p = 0.017
Survival Probability
1.00
0.75
0.50
0.25
0.00
Number at risk
50 24 19 17
49 26 22 21
50 33 30 29
0 30 60 90
Days
Mortality Rate
50%
40%
30%
20%
10%
0%
46%
43%
20%
T1 T2 T3
TNF-α Tertile
12.5
152.5
Count
10
5
0
1 4 16 64 256 1024 4096 16384
CSF TNF-α (pg/mL)
CCL5
CCL5 Tertile
T1
T2
T3
p = 0.022
Survival Probability
1.00
0.75
0.50
0.25
0.00
Number at risk
50 22 17 17
49 29 25 22
50 32 29 28
0 30 60 90
Days
Mortality Rate
50%
40%
30%
20%
10%
0%
48%
37%
24%
T1 T2 T3
CCL5 Tertile
18.6
67.2
Count
15
10
5
0
4 16 64 256 1024
CSF CCL5 (pg/mL)

# CD40-L

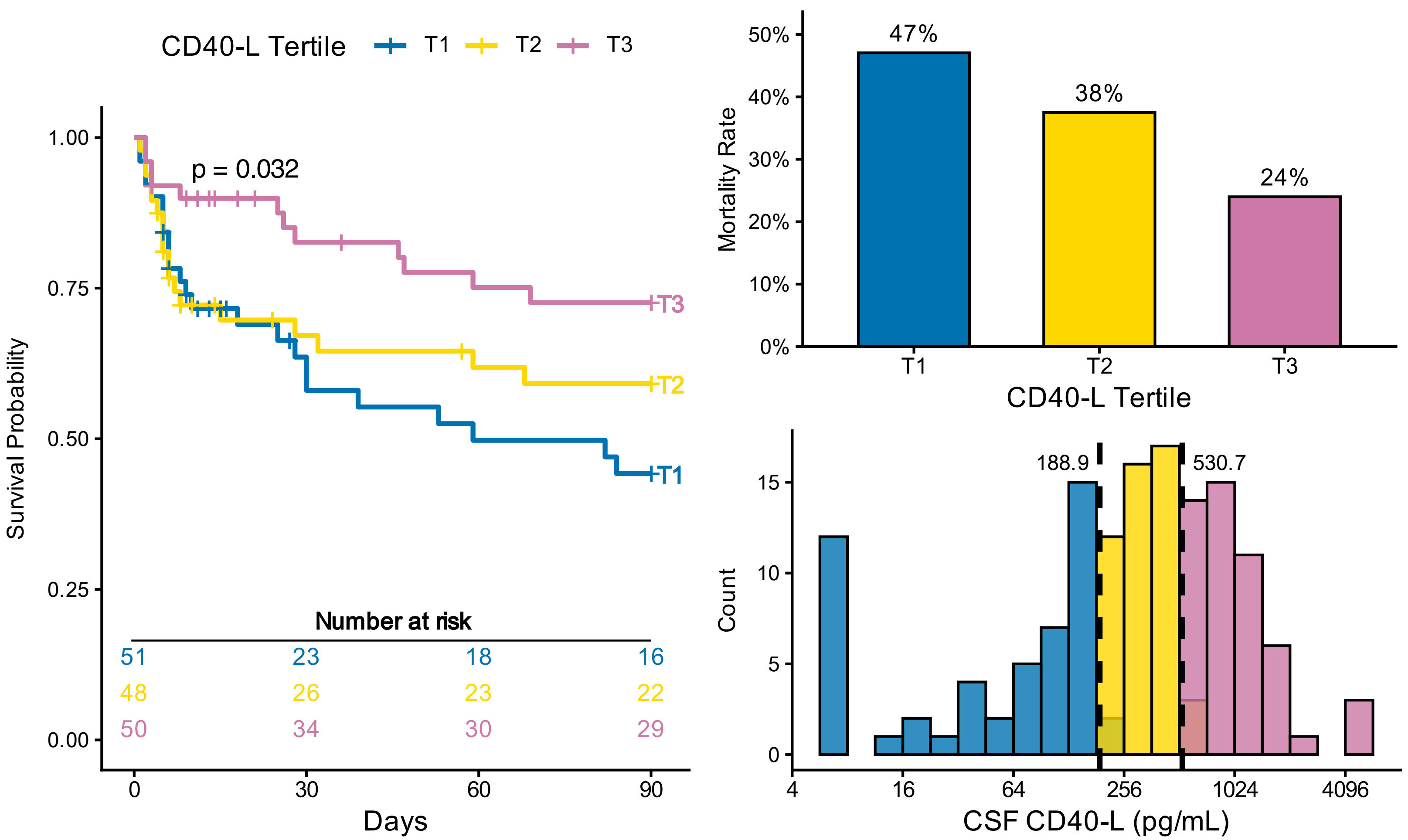
CD40-L Tertile
T1
T2
T3
p = 0.032
Survival Probability
1.00
0.75
0.50
0.25
0.00
Number at risk
51 23 18 16
48 26 23 22
50 34 30 29
0 30 60 90
Days
Mortality Rate
50%
40%
30%
20%
10%
0%
47%
38%
24%
CD40-L Tertile
Count
15
10
5
0
188.9
530.7
4 16 64 256 1024 4096
CSF CD40-L (pg/mL)

### CXCL2

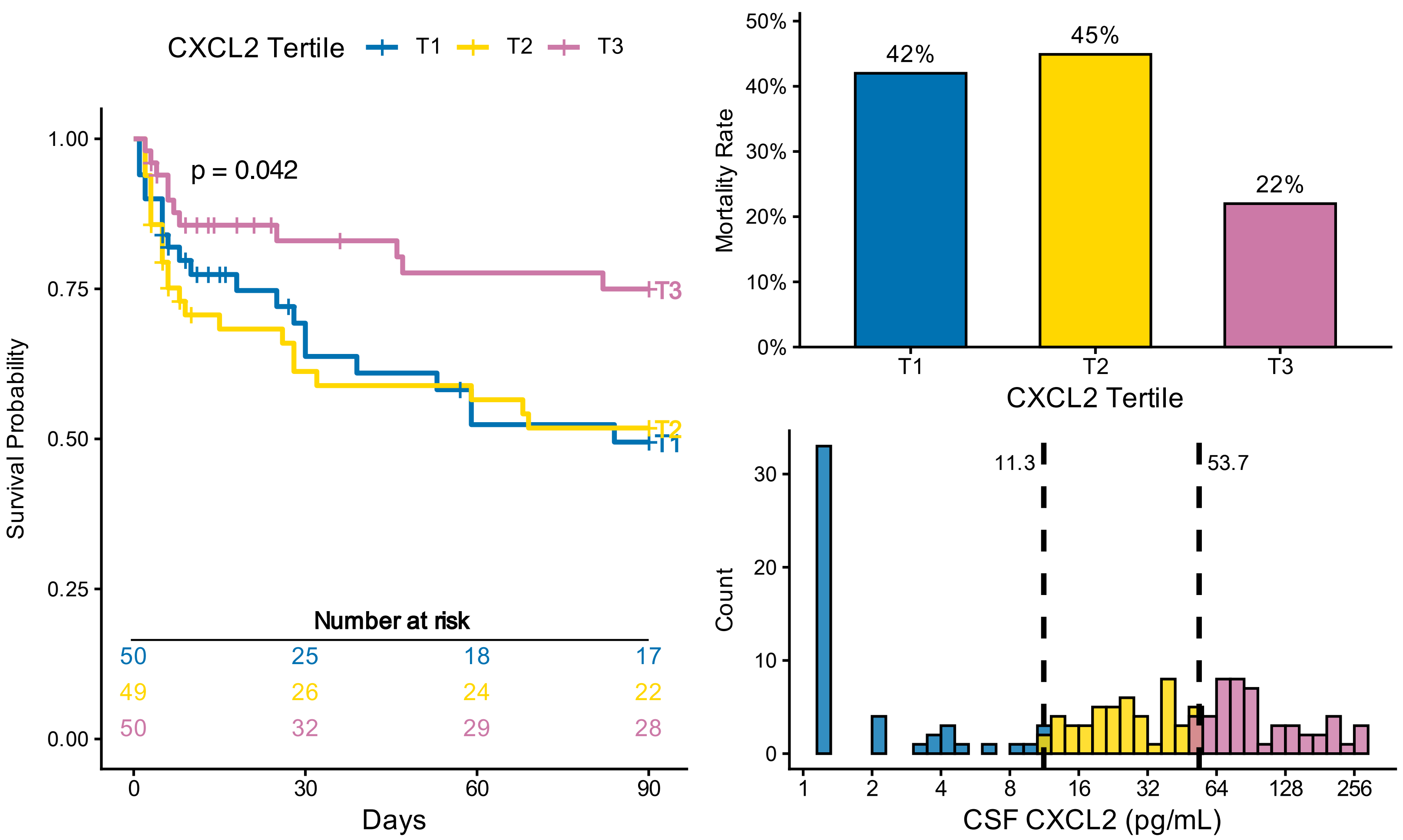
CXCL2 Tertile
T1
T2
T3
p = 0.042
Survival Probability
1.00
0.75
0.50
0.25
0.00
Number at risk
50 25 18 17
49 26 24 22
50 32 29 28
0 30 60 90
Days
Mortality Rate
50%
40%
30%
20%
10%
0%
42%
45%
22%
CXCL2 Tertile
Count
30
20
10
0
11.3
53.7
1 2 4 8 16 32 64 128 256
CSF CXCL2 (pg/mL)

# IL-2

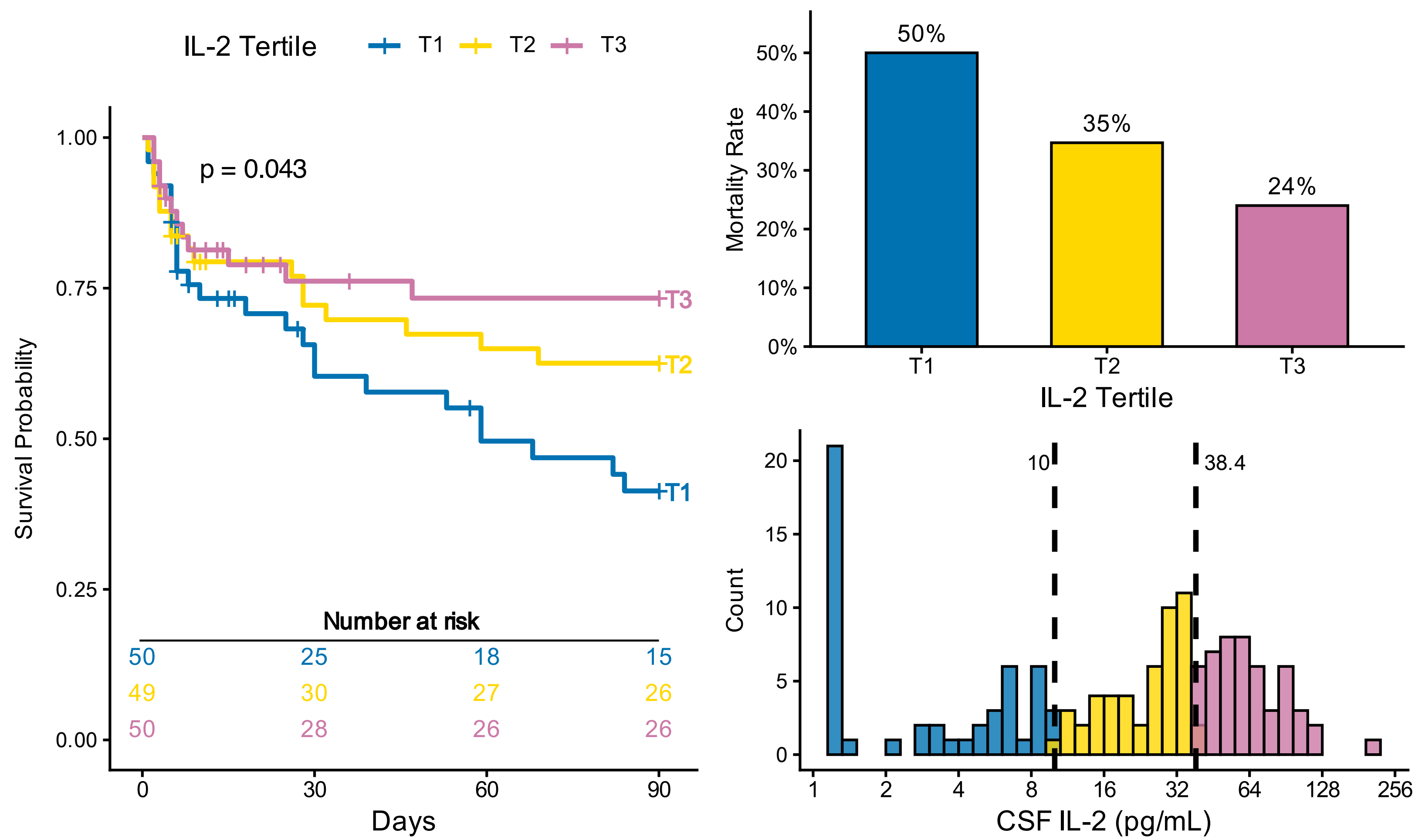
IL-2 Tertile
T1
T2
T3
p = 0.043
Survival Probability
1.00
0.75
0.50
0.25
0.00
Number at risk
50 25 18 15
49 30 27 26
50 28 26 26
0
30
60
90
Days
50%
35%
24%
Mortality Rate
IL-2 Tertile
10
38.4
Count
CSF IL-2 (pg/mL)

# PD-L1

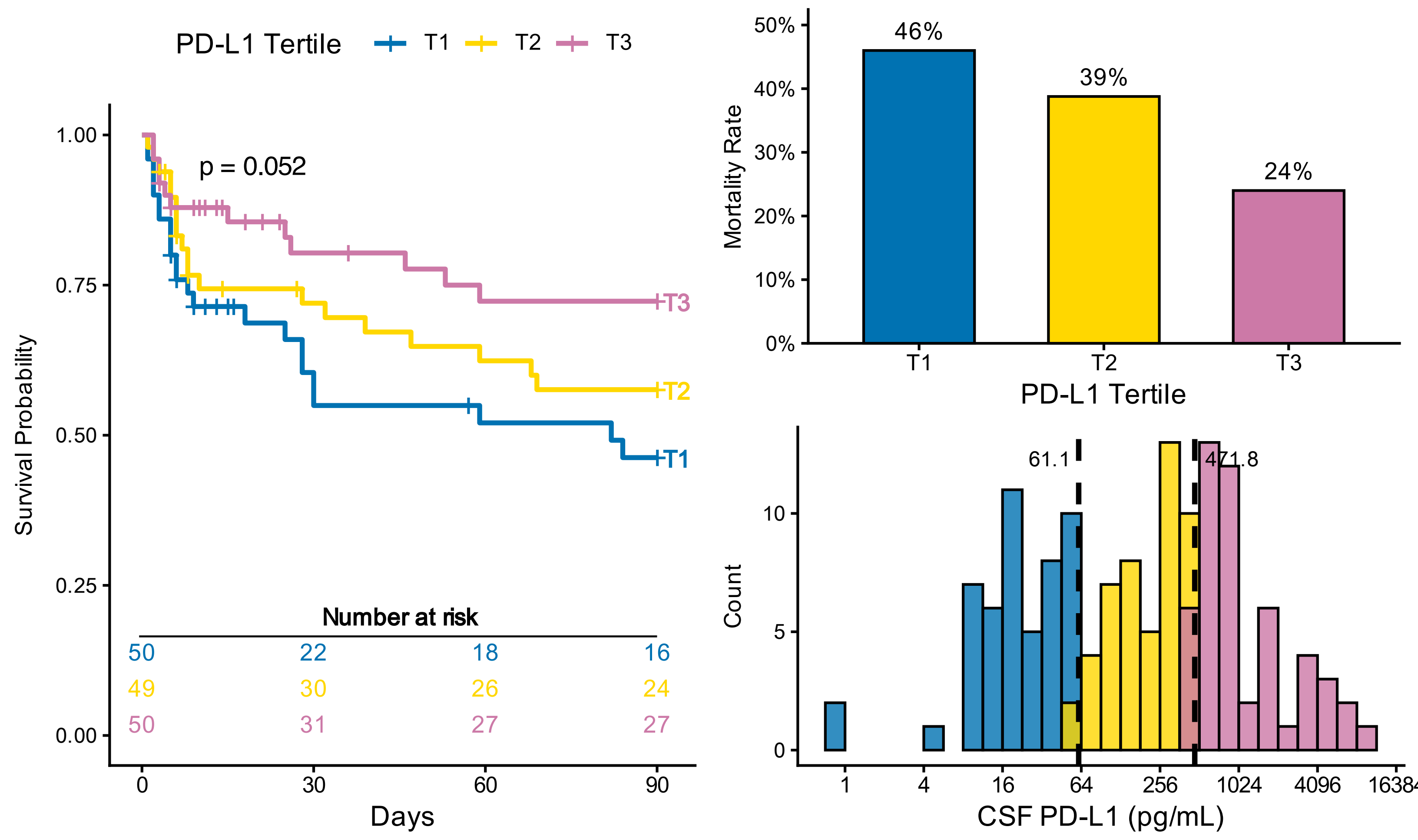
PD-L1 Tertile
T1
T2
T3
p = 0.052
Survival Probability
1.00
0.75
0.50
0.25
0.00
Number at risk
50 22 18 16
49 30 26 24
50 31 27 27
0
30
60
90
Days
46%
39%
24%
Mortality Rate
PD-L1 Tertile
61.1
471.8
Count
CSF PD-L1 (pg/mL)

### TGF-α

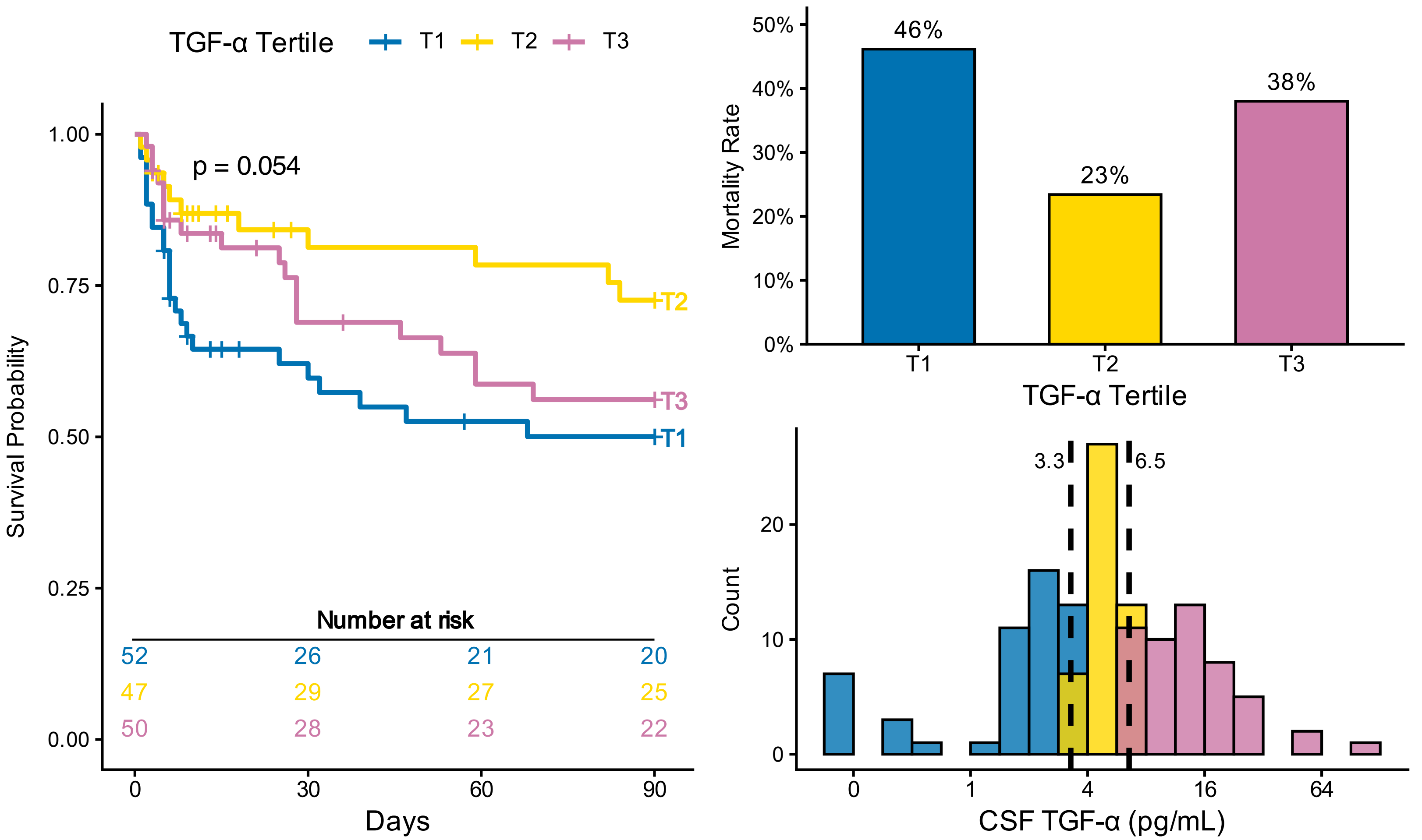

TGF-α Tertile
T1
T2
T3
p = 0.054
Survival Probability
1.00
0.75
0.50
0.25
0.00
Number at risk
52 26 21 20
47 29 27 25
50 28 23 22
0
30
60
90
Days
Mortality Rate
50%
40%
30%
20%
10%
0%
46%
23%
38%
TGF-α Tertile
3.3
6.5
Count
20
10
0
0
1
4
16
64
CSF TGF-α (pg/mL)


### CCL19

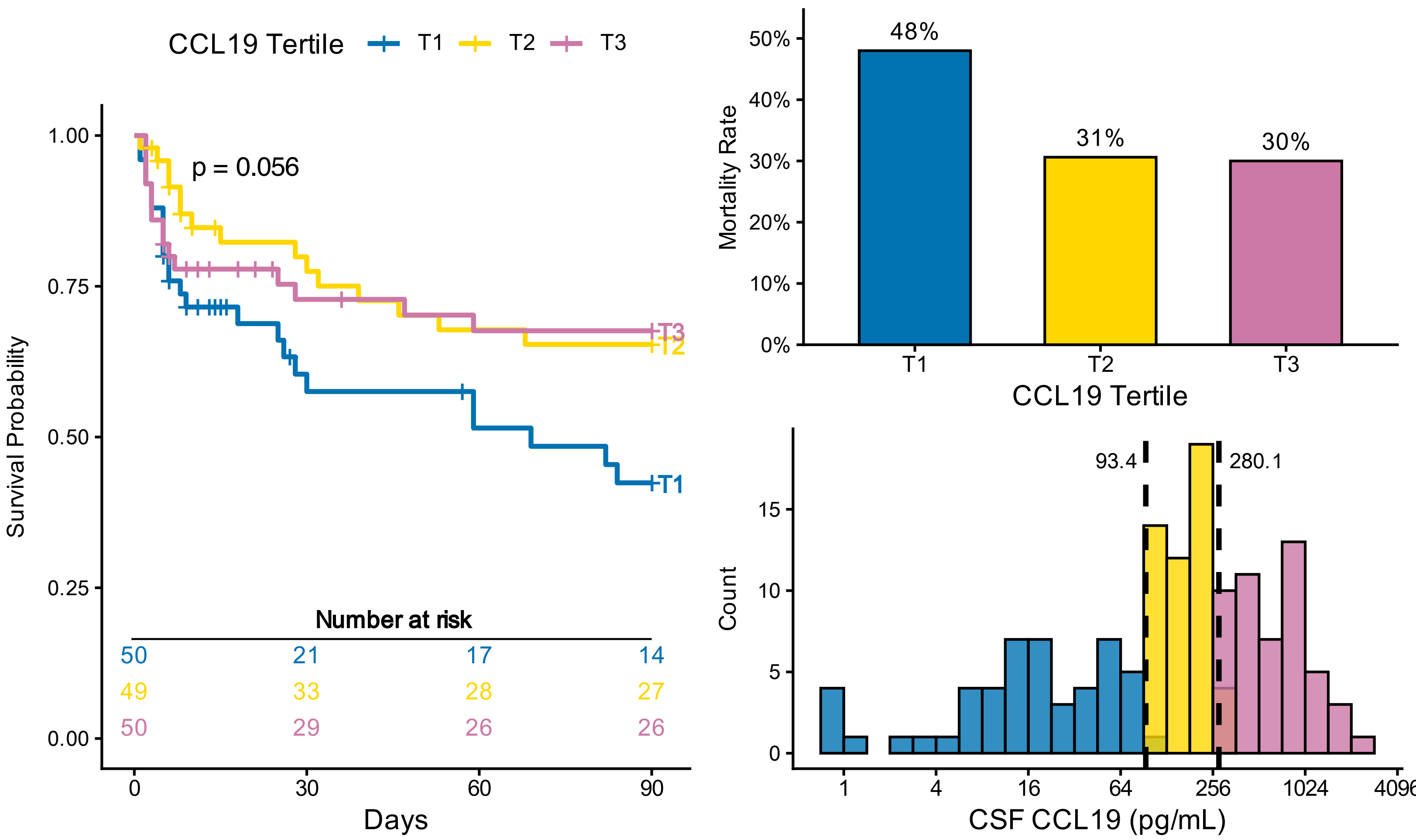

CCL19 Tertile
T1
T2
T3
p = 0.056
Survival Probability
1.00
0.75
0.50
0.25
0.00
Number at risk
50 21 17 14
49 33 28 27
50 29 26 26
0
30
60
90
Days
Mortality Rate
50%
40%
30%
20%
10%
0%
48%
31%
30%
CCL19 Tertile
93.4
280.1
Count
15
10
5
0
1
4
16
64
256
1024
4096
CSF CCL19 (pg/mL)

### CXCL1

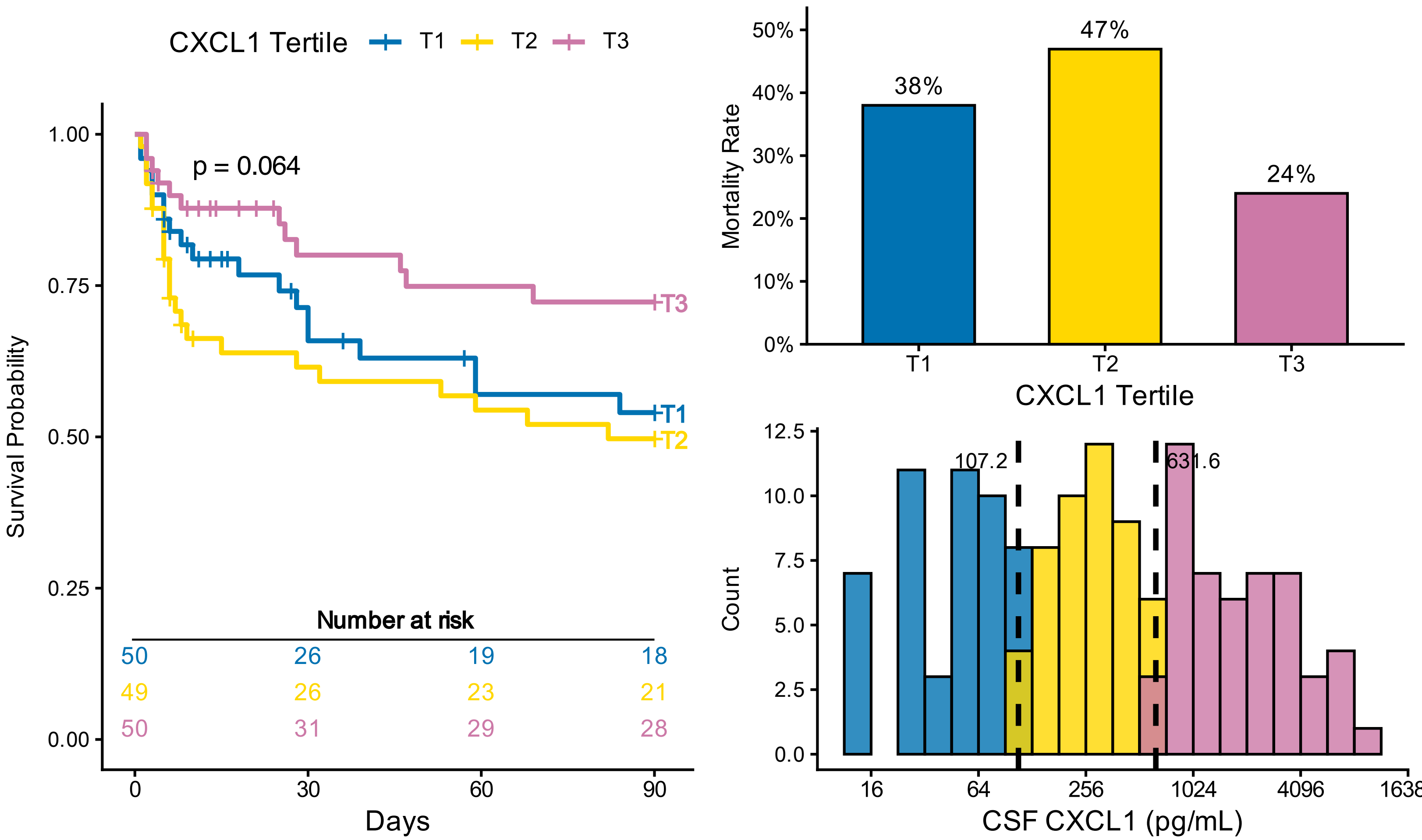

CXCL1 Tertile
T1
T2
T3
p = 0.064
Survival Probability
1.00
0.75
0.50
0.25
0.00
Number at risk
50 26 19 18
49 26 23 21
50 31 29 28
0
30
60
90
Days
Mortality Rate
50%
40%
30%
20%
10%
0%
38%
47%
24%
CXCL1 Tertile
Count
12.5
10.0
7.5
5.0
2.5
0.0
107.2
631.6
16
64
256
1024
4096
1638
CSF CXCL1 (pg/mL)


### CCL3

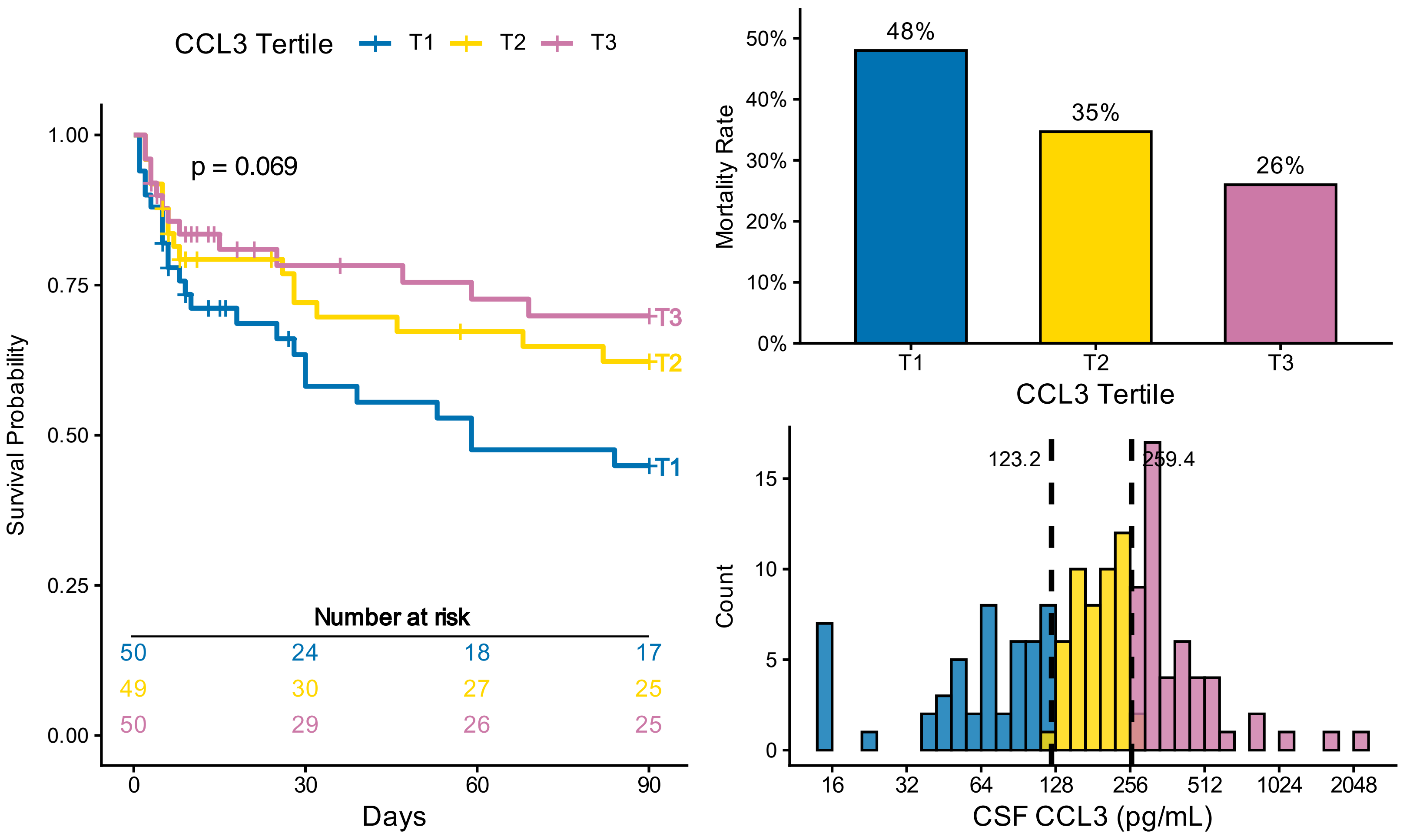

CCL3 Tertile
T1
T2
T3
p = 0.069
Survival Probability
1.00
0.75
0.50
0.25
0.00
Number at risk
50 24 18 17
49 30 27 25
50 29 26 25
0
30
60
90
Days
Mortality Rate
50%
40%
30%
20%
10%
0%
48%
35%
26%
CCL3 Tertile
Count
15
10
5
0
123.2
259.4
16
32
64
128
256
512
1024
2048
CSF CCL3 (pg/mL)

### TRAIL

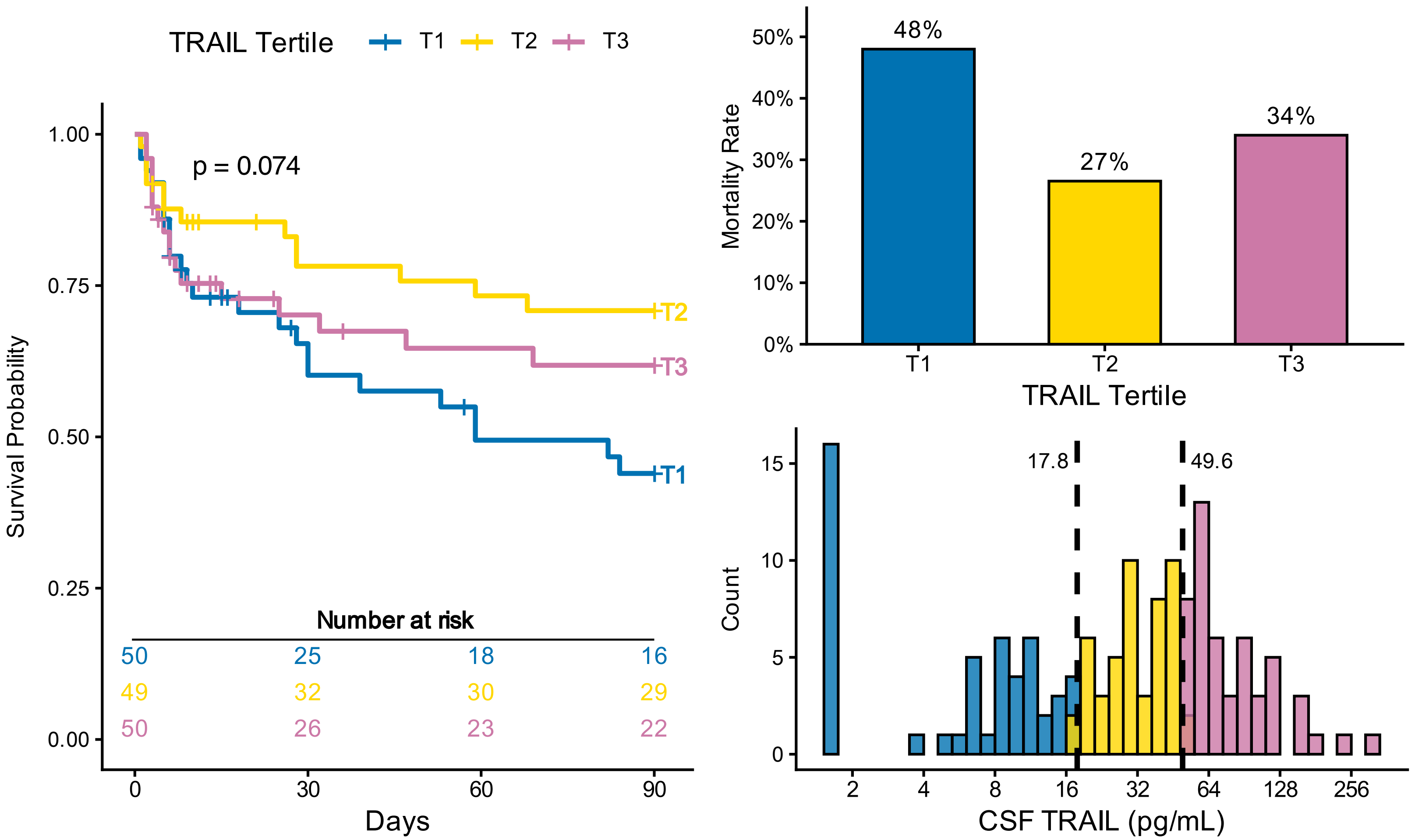
TRAIL Tertile T1 T2 T3
p = 0.074
Survival Probability
1.00
0.75
0.50
0.25
0.00
T2
T3
T1
Number at risk
50 25 18 16
49 32 30 29
50 26 23 22
0 30 60 90
Days
Mortality Rate
50%
40%
30%
20%
10%
0%
48%
27%
34%
T1 T2 T3
TRAIL Tertile
Count
15
10
5
0
17.8
49.6
2 4 8 16 32 64 128 256
CSF TRAIL (pg/mL)

# IL-1β

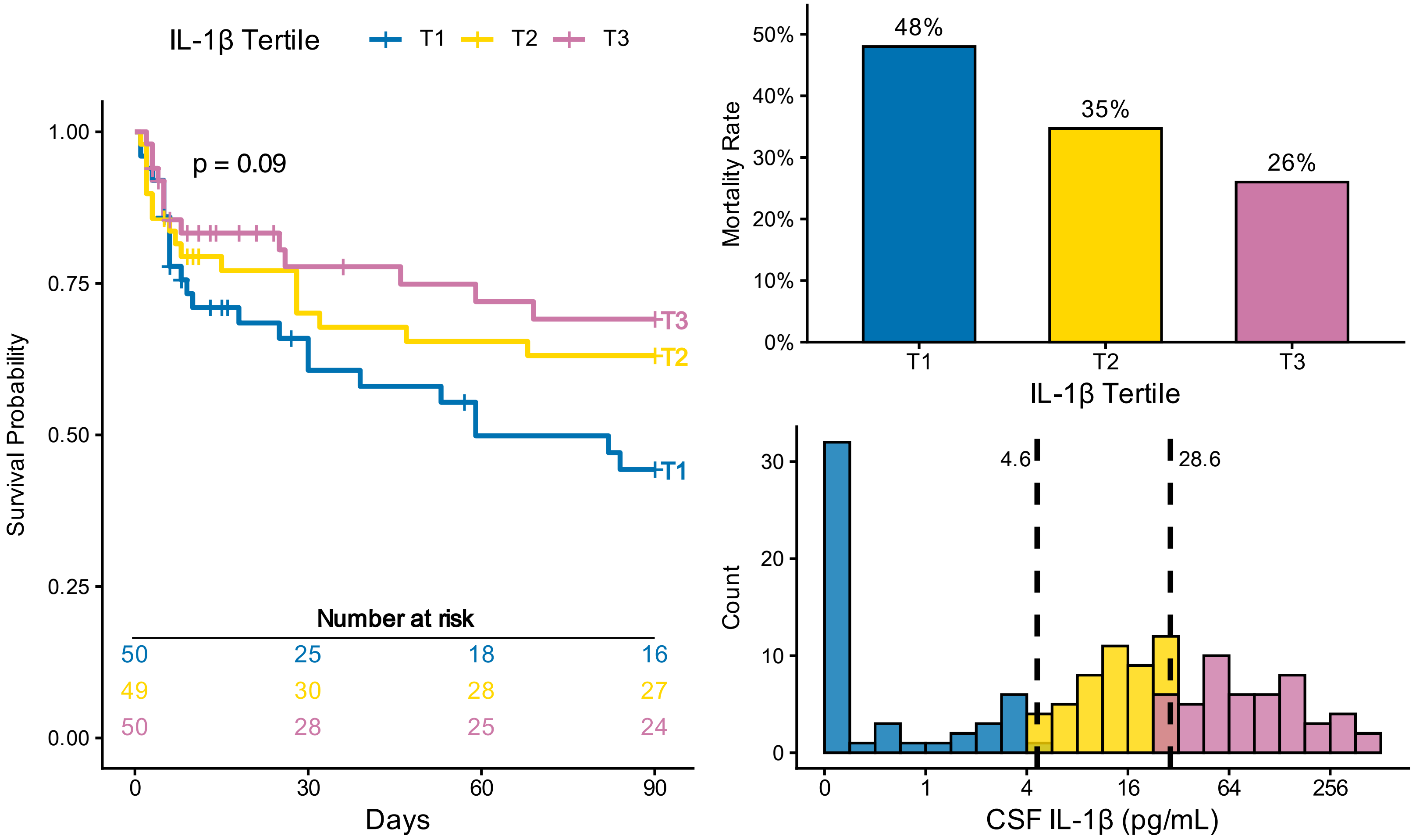
IL-1β Tertile T1 T2 T3
p = 0.09
Survival Probability
1.00
0.75
0.50
0.25
0.00
T3
T2
T1
Number at risk
50 25 18 16
49 30 28 27
50 28 25 24
0 30 60 90
Days
Mortality Rate
50%
40%
30%
20%
10%
0%
48%
35%
26%
T1 T2 T3
IL-1β Tertile
Count
30
20
10
0
4.6
28.6
0 1 4 16 64 256
CSF IL-1β (pg/mL)

### GZMB

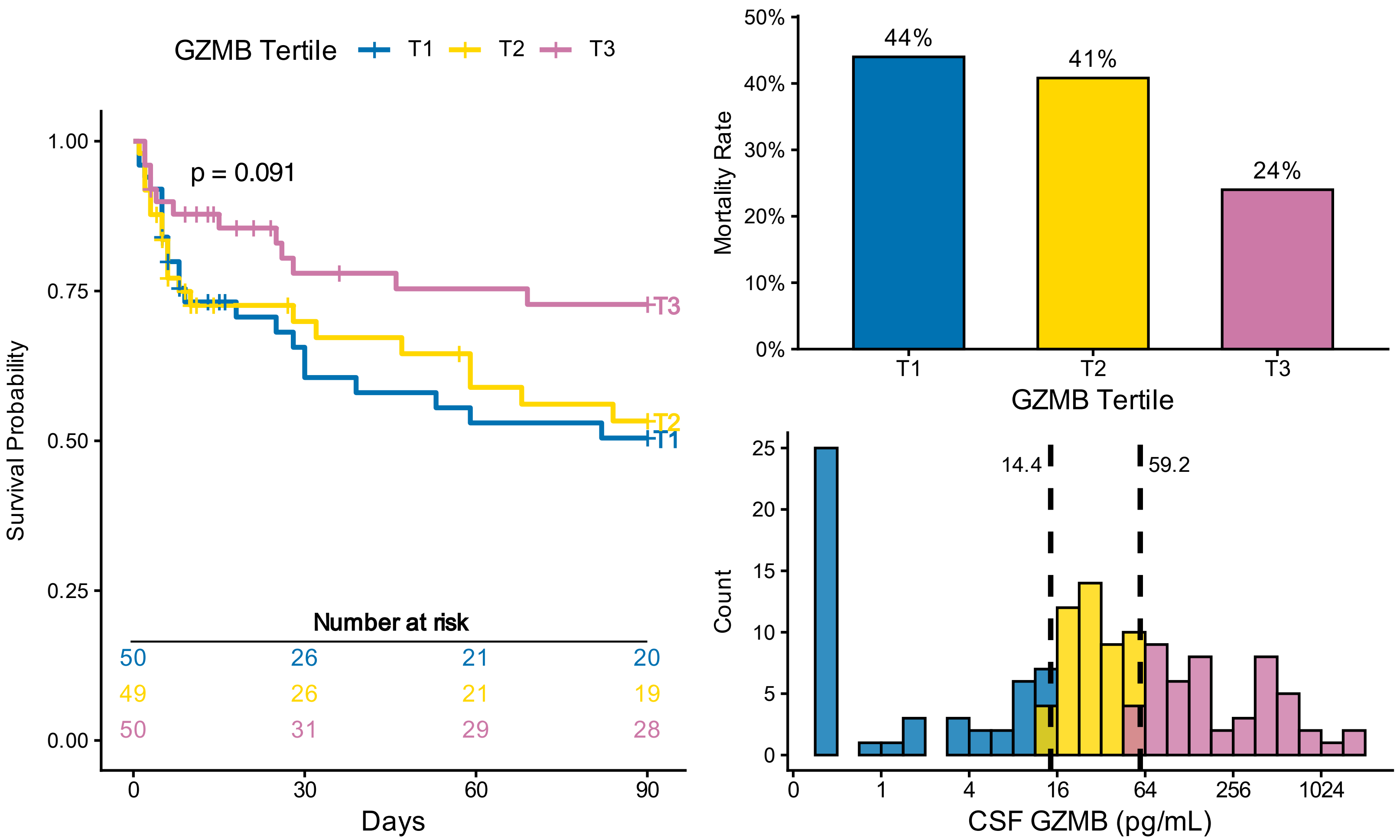
GZMB Tertile
T1
T2
T3
p = 0.091
Survival Probability
1.00
0.75
0.50
0.25
0.00
Number at risk
50 26 21 20
49 26 21 19
50 31 29 28
0
30
60
90
Days
Mortality Rate
50%
40%
30%
20%
10%
0%
44%
41%
24%
GZMB Tertile
Count
14.4
59.2
CSF GZMB (pg/mL)
0
1
4
16
64
256
1024

### CXCL10

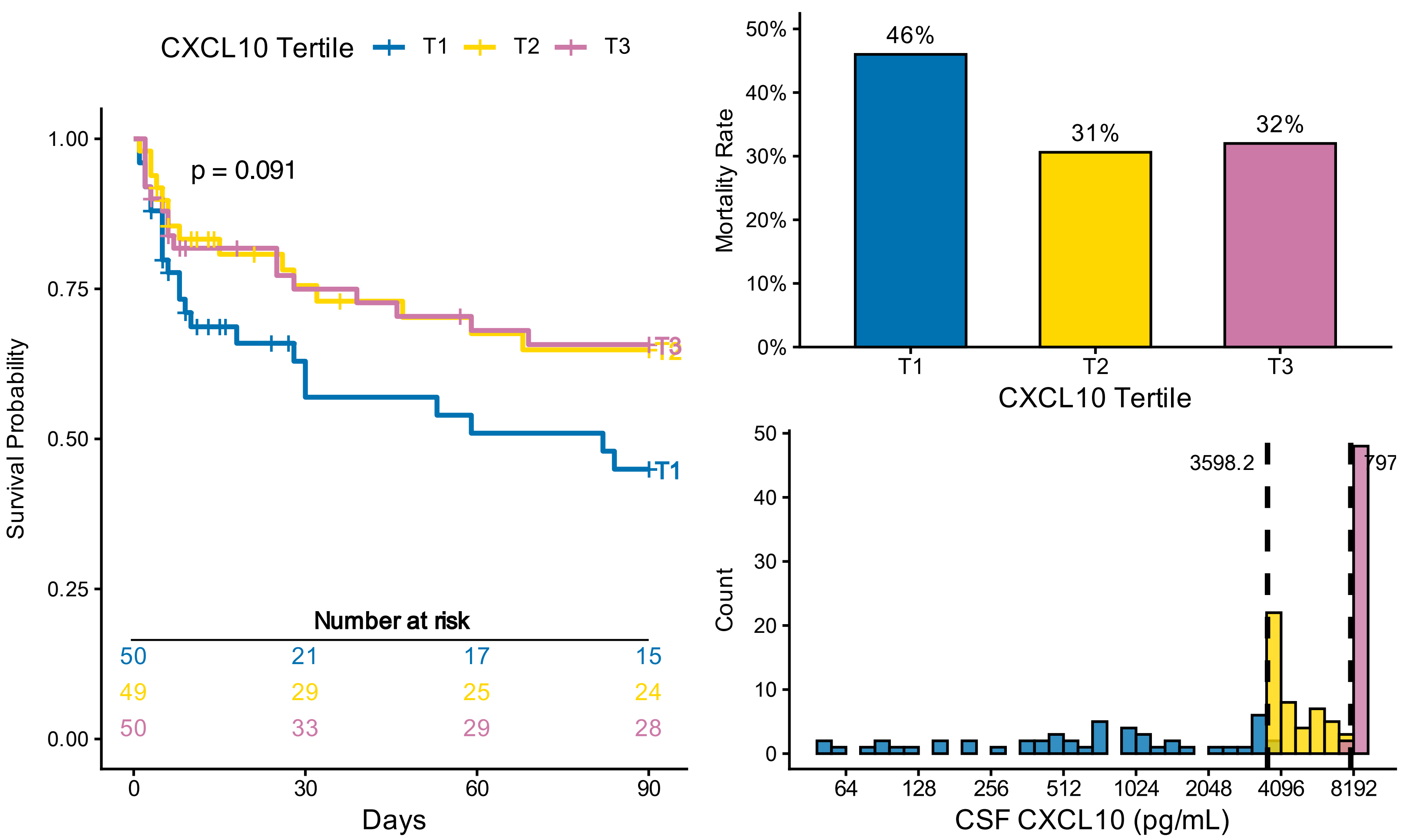
CXCL10 Tertile
T1
T2
T3
p = 0.091
Survival Probability
1.00
0.75
0.50
0.25
0.00
Number at risk
50 21 17 15
49 29 25 24
50 33 29 28
0
30
60
90
Days
Mortality Rate
50%
40%
30%
20%
10%
0%
46%
31%
32%
CXCL10 Tertile
Count
3598.2
797
CSF CXCL10 (pg/mL)
64
128
256
512
1024
2048
4096
8192

### PDGFaa

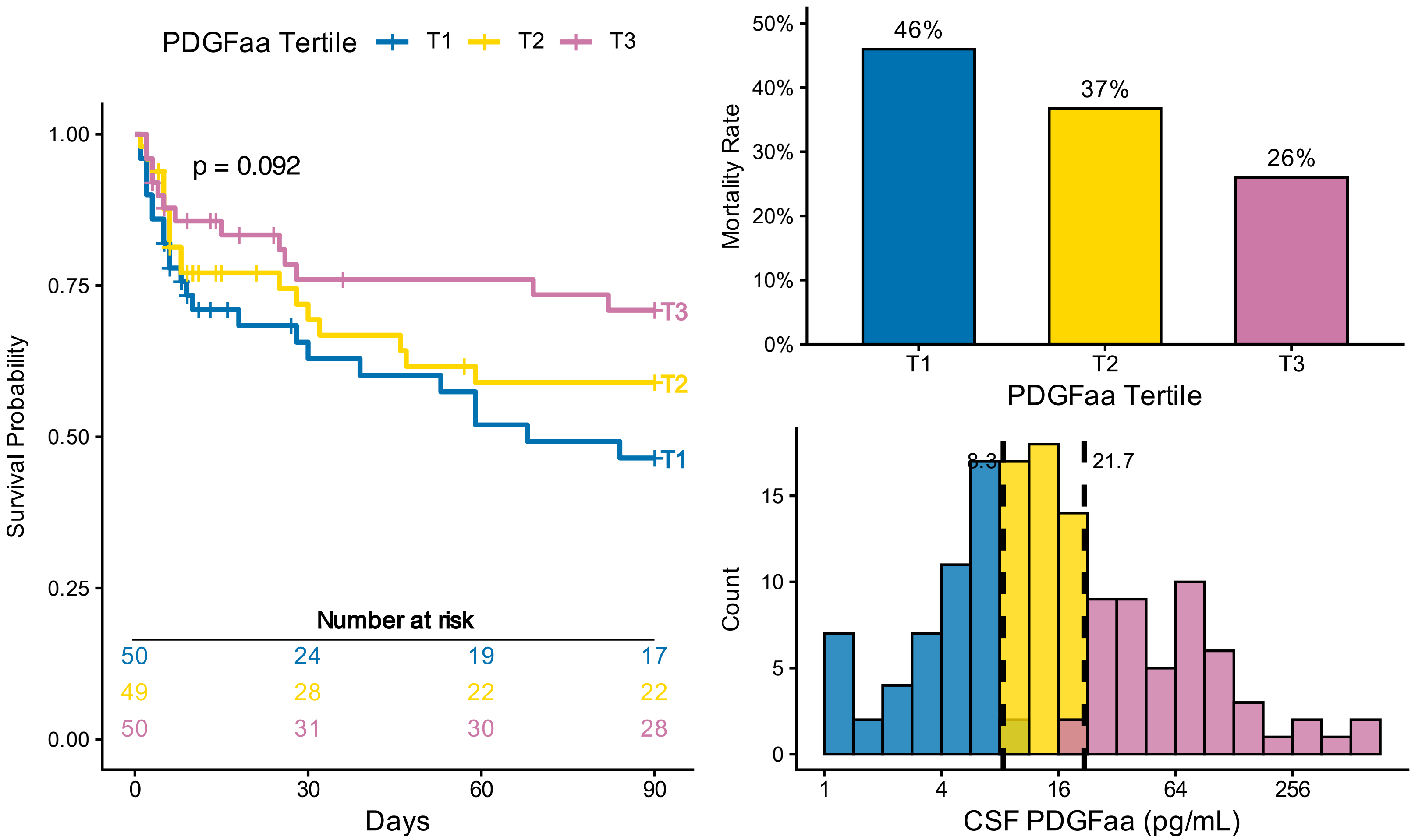


### CCL20

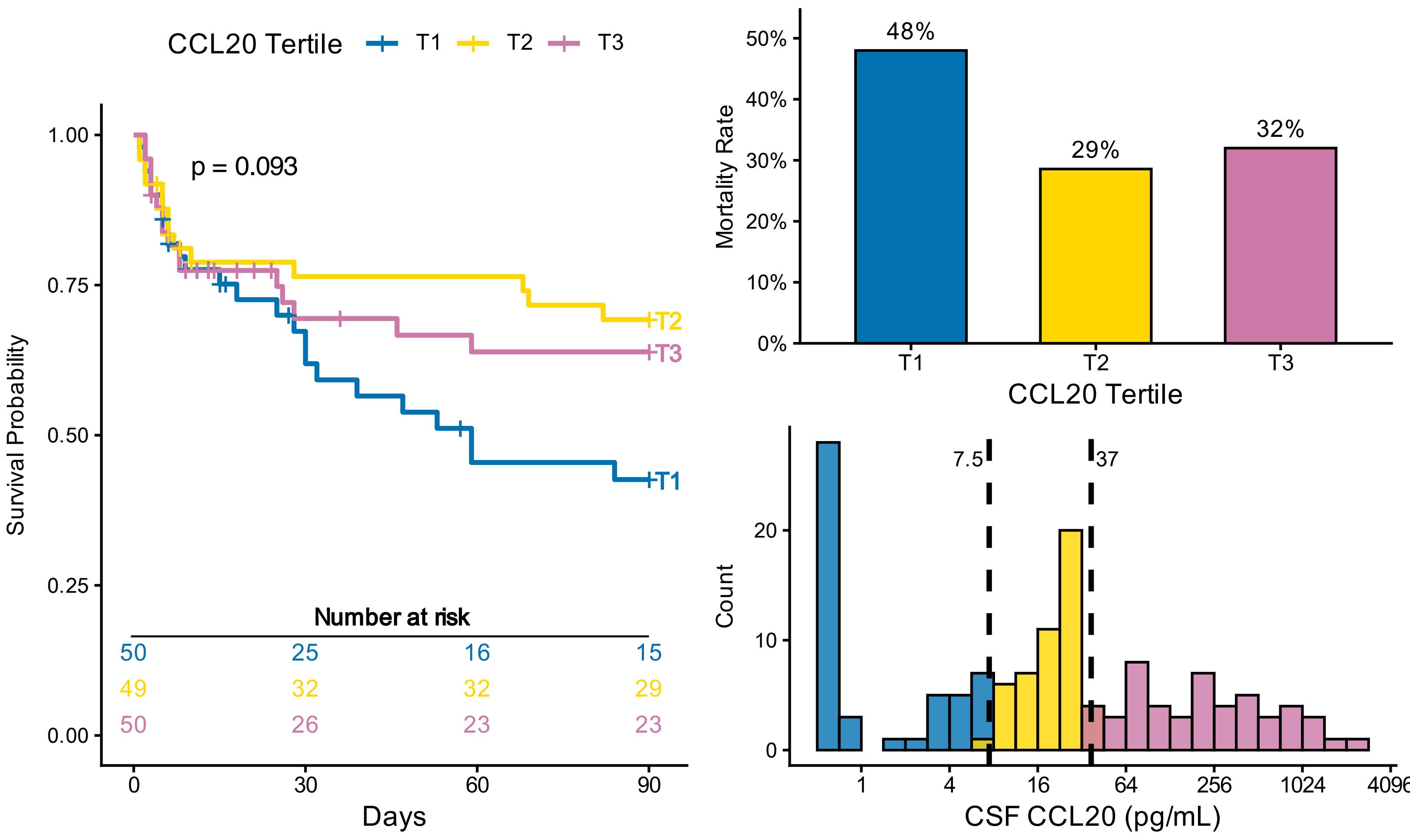

# IL-1α

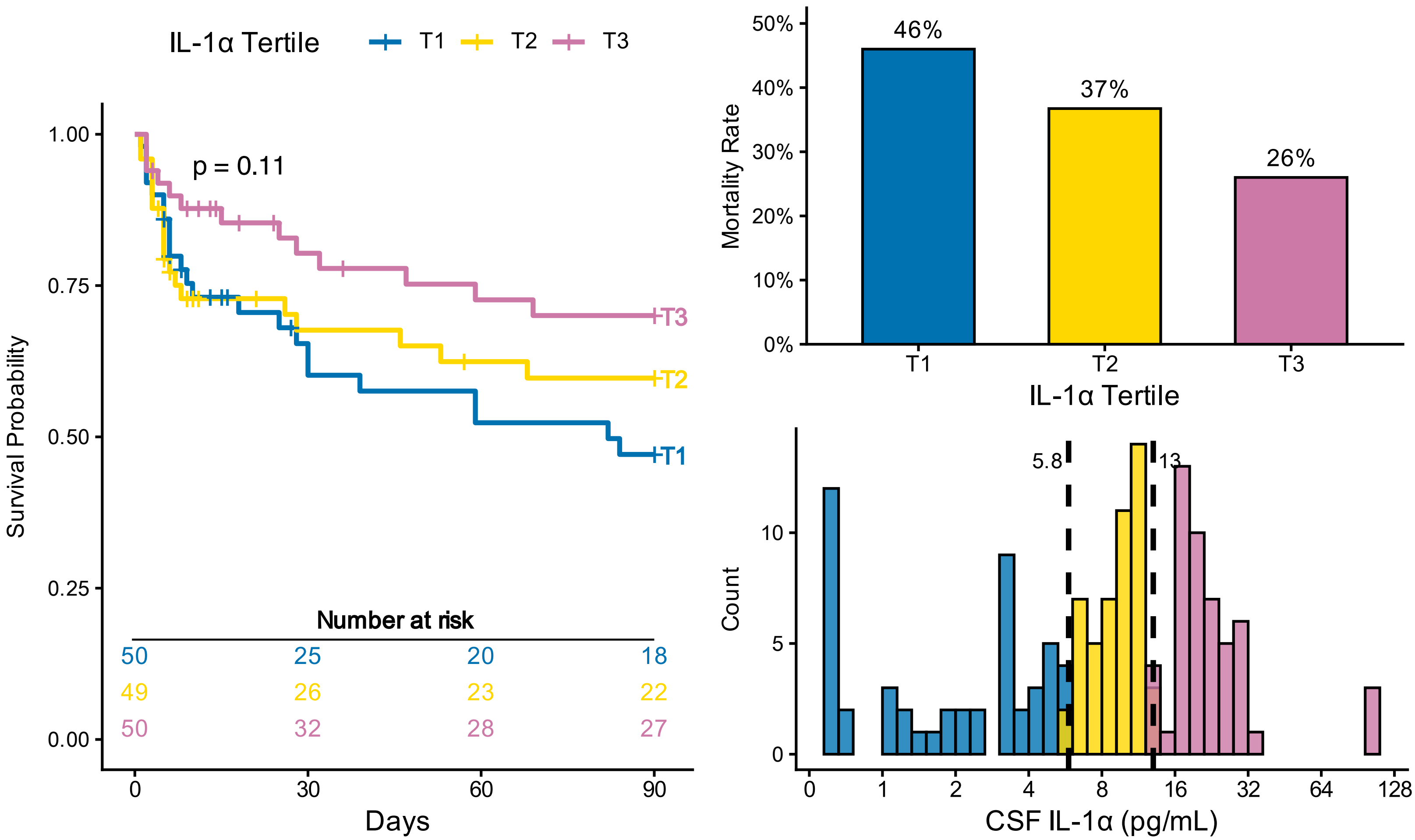
IL-1α Tertile
T1
T2
T3
p = 0.11
Survival Probability
1.00
0.75
0.50
0.25
0.00
Number at risk
50 25 20 18
49 26 23 22
50 32 28 27
0
30
60
90
Days
Mortality Rate
50%
40%
30%
20%
10%
0%
46%
37%
26%
IL-1α Tertile
Count
5.8
13
CSF IL-1α (pg/mL)
0 1 2 4 8 16 32 64 128

### FGF-2

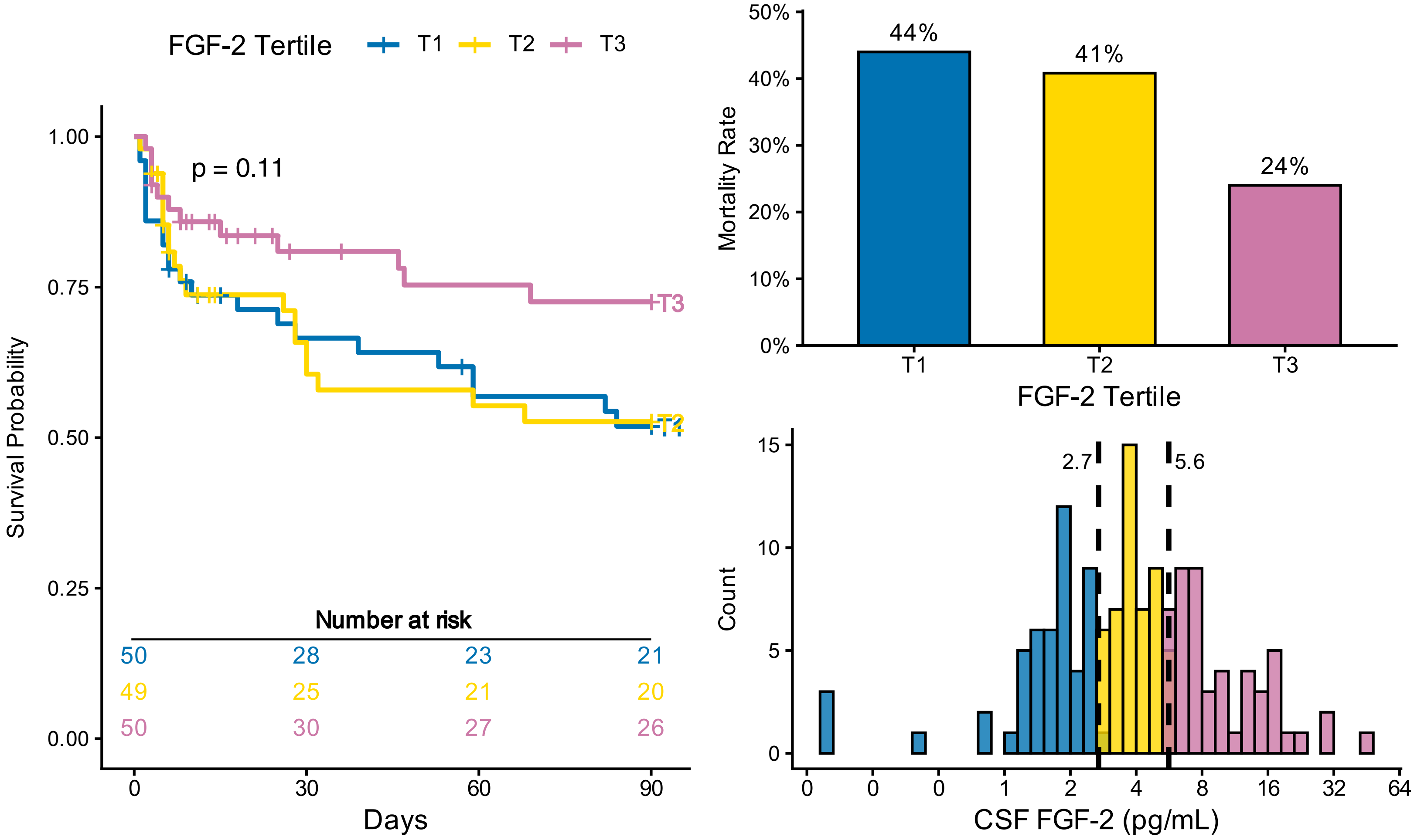
FGF-2 Tertile
T1
T2
T3
p = 0.11
Survival Probability
1.00
0.75
0.50
0.25
0.00
Number at risk
50 28 23 21
49 25 21 20
50 30 27 26
0
30
60
90
Days
Mortality Rate
50%
40%
30%
20%
10%
0%
44%
41%
24%
FGF-2 Tertile
Count
15
10
5
0
2.7
5.6
CSF FGF-2 (pg/mL)
0 0 0 1 2 4 8 16 32 64

# IL-10

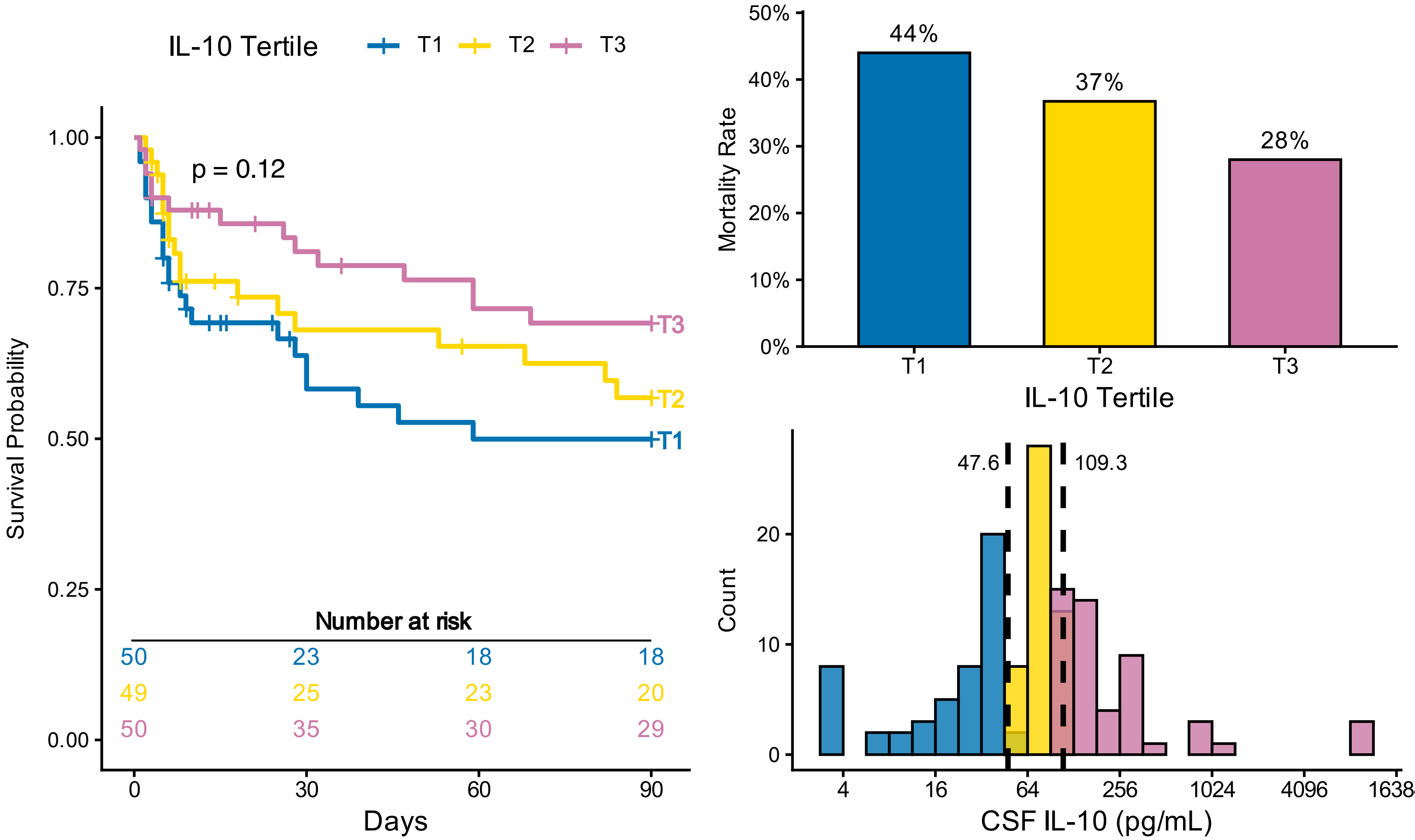
IL-10 Tertile
T1
T2
T3
p = 0.12
Survival Probability
1.00
0.75
0.50
0.25
0.00
Number at risk
50 23 18 18
49 25 23 20
50 35 30 29
0
30
60
90
Days
Mortality Rate
50%
40%
30%
20%
10%
0%
44%
37%
28%
IL-10 Tertile
Count
47.6
109.3
4
16
64
256
1024
4096
CSF IL-10 (pg/mL)

### PDGFab

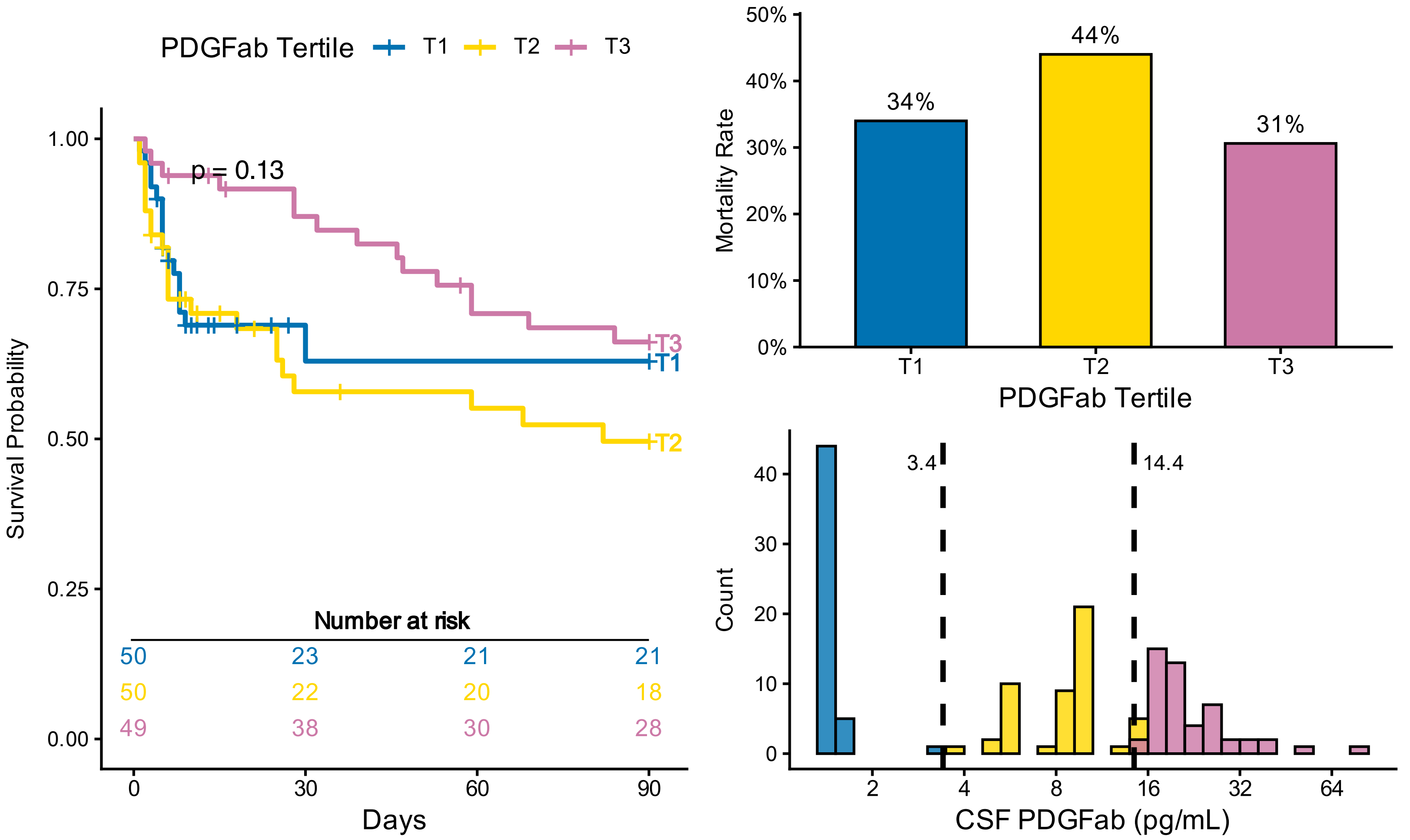
PDGFab Tertile
T1
T2
T3
p = 0.13
Survival Probability
1.00
0.75
0.50
0.25
0.00
Number at risk
50 23 21 21
50 22 20 18
49 38 30 28
0
30
60
90
Days
Mortality Rate
50%
40%
30%
20%
10%
0%
34%
44%
31%
PDGFab Tertile
Count
3.4
14.4
2
4
8
16
32
64
CSF PDGFab (pg/mL)

### IFN-α

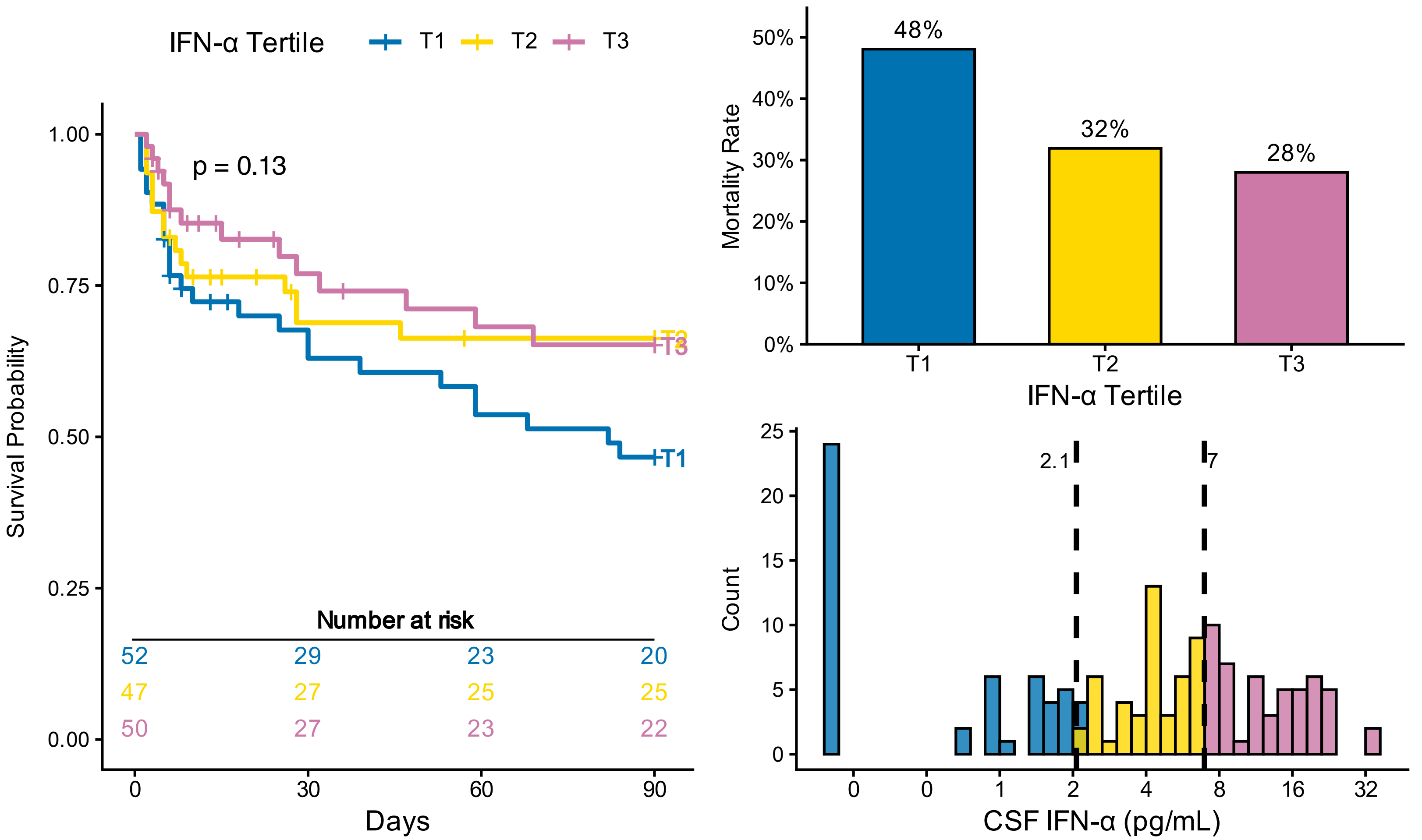
IFN-α Tertile
T1
T2
T3
p = 0.13
Survival Probability
Number at risk
52 29 23 20
47 27 25 25
50 27 23 22
Days
Mortality Rate
48%
32%
28%
IFN-α Tertile
Count
2.1
7
CSF IFN-α (pg/mL)

### Flt3-L

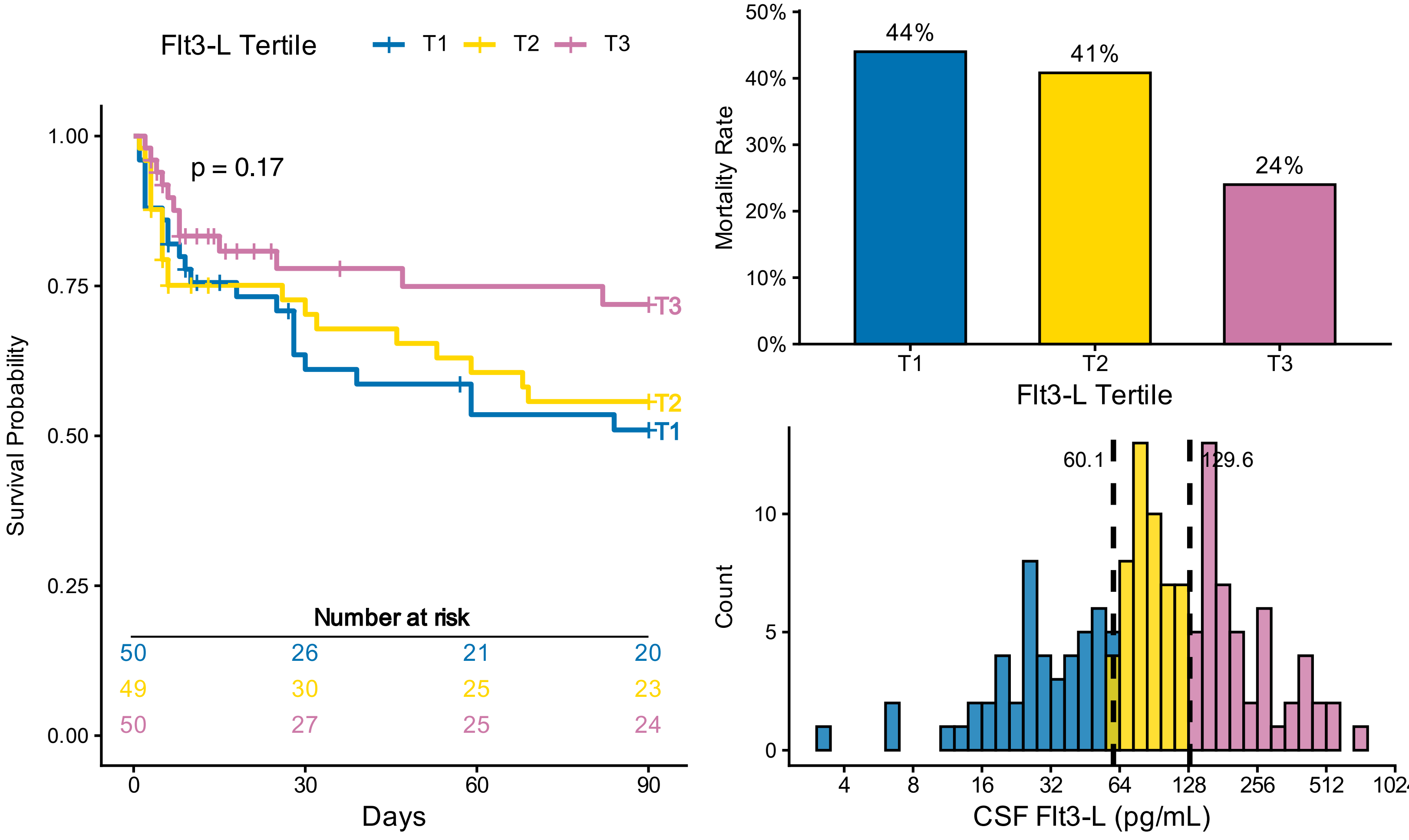
Flt3-L Tertile
T1
T2
T3
p = 0.17
Survival Probability
Number at risk
50 26 21 20
49 30 25 23
50 27 25 24
Days
Mortality Rate
44%
41%
24%
Flt3-L Tertile
Count
60.1
129.6
CSF Flt3-L (pg/mL)

# IL-12

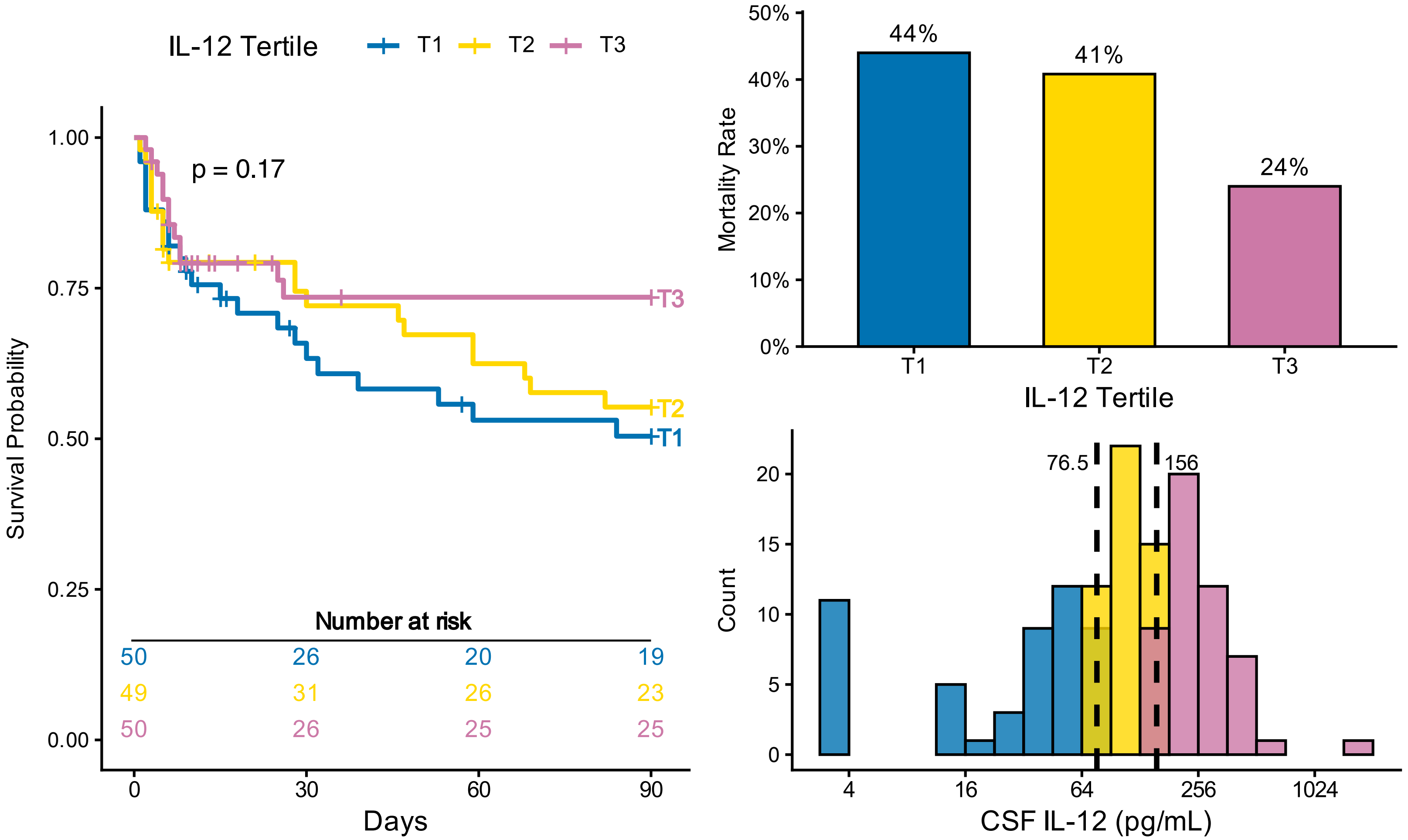
IL-12 Tertile
T1
T2
T3
p = 0.17
Survival Probability
1.00
0.75
0.50
0.25
0.00
Number at risk
50 26 20 19
49 31 26 23
50 26 25 25
0
30
60
90
Days
Mortality Rate
50%
40%
30%
20%
10%
0%
44%
41%
24%
IL-12 Tertile
Count
76.5
156
4
16
64
256
1024
CSF IL-12 (pg/mL)

# IL-33

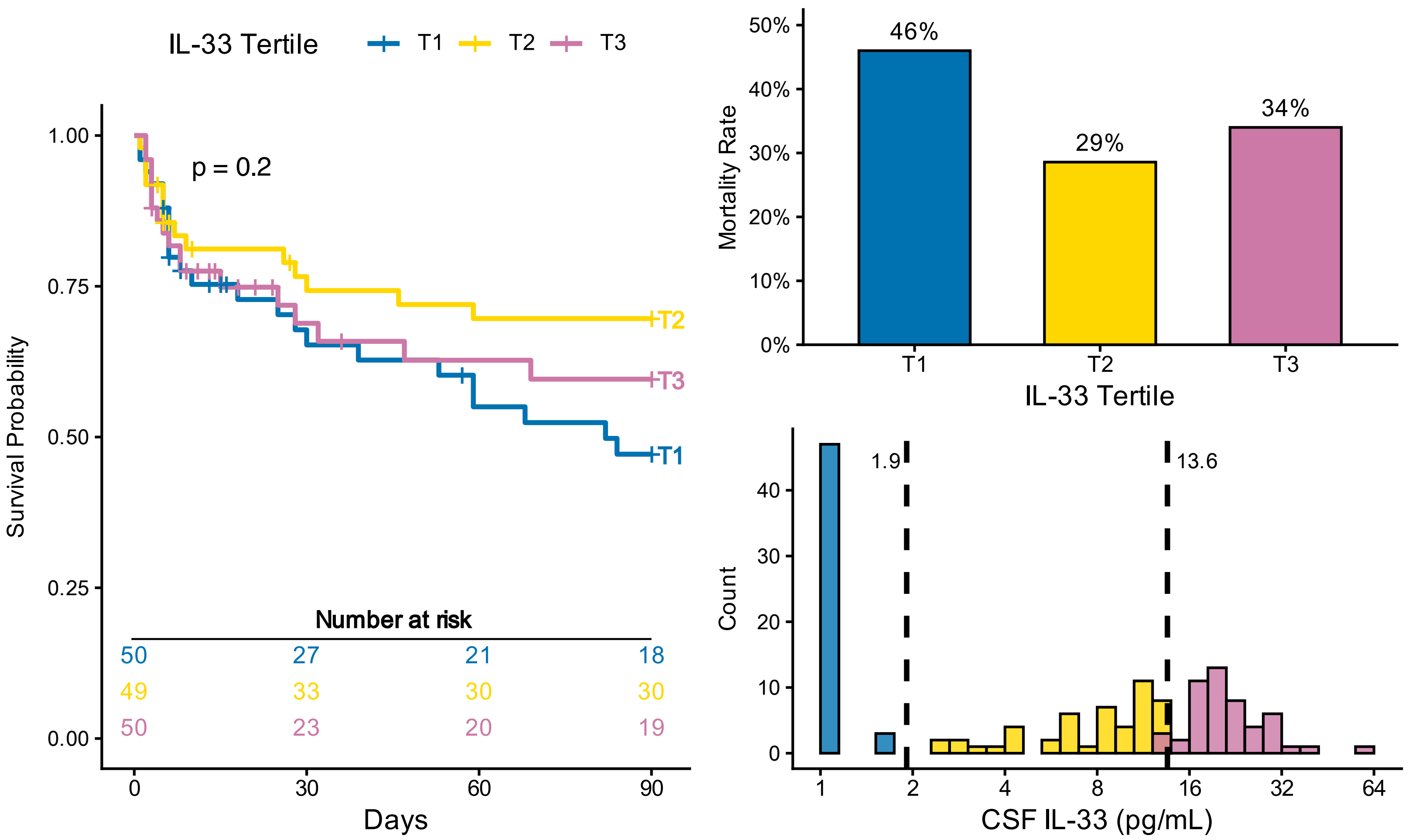
IL-33 Tertile
T1
T2
T3
p = 0.2
Survival Probability
1.00
0.75
0.50
0.25
0.00
Number at risk
50 27 21 18
49 33 30 30
50 23 20 19
0
30
60
90
Days
Mortality Rate
50%
40%
30%
20%
10%
0%
46%
29%
34%
IL-33 Tertile
Count
1.9
13.6
1
2
4
8
16
32
64
CSF IL-33 (pg/mL)

### CCL11

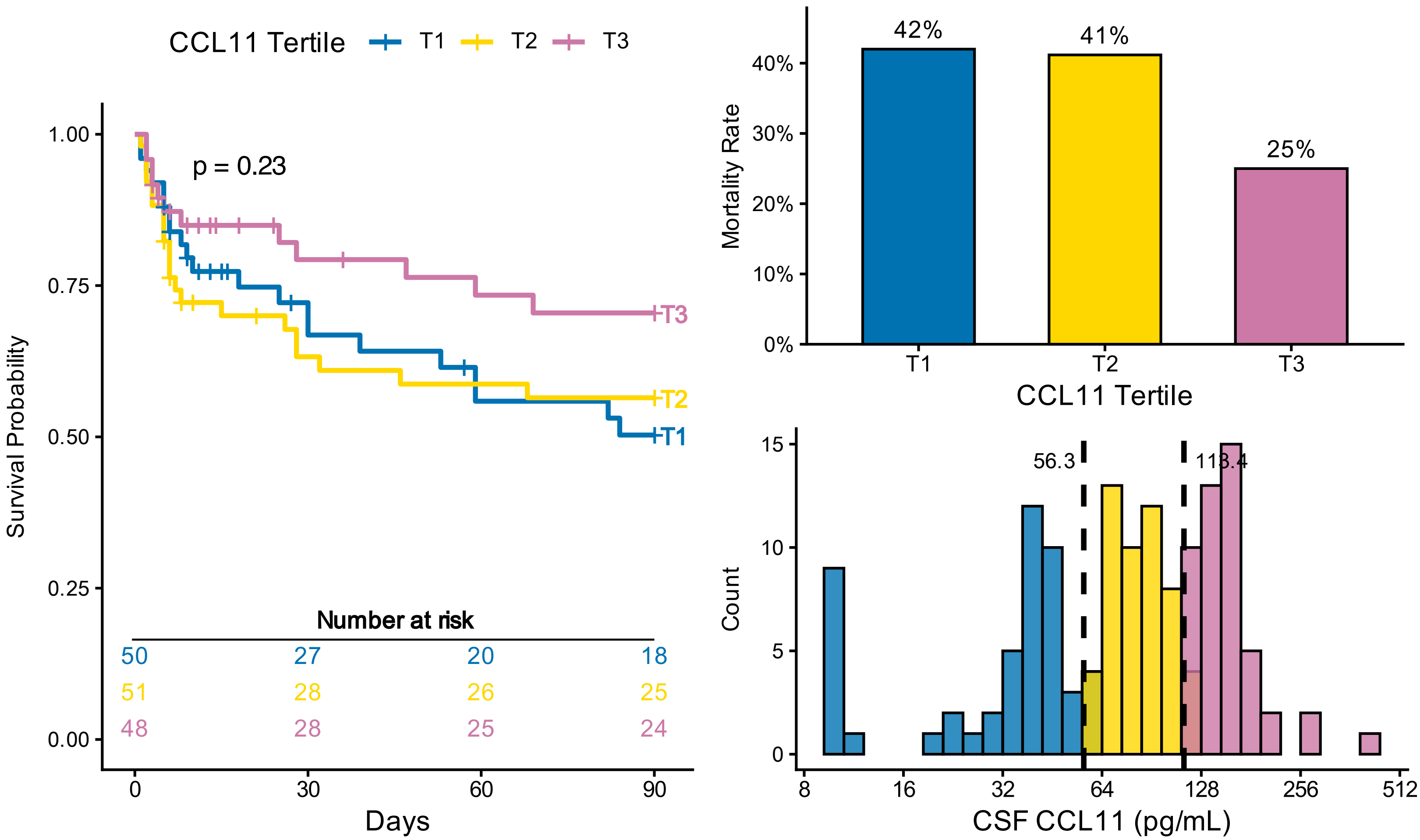

CCL11 Tertile
T1
T2
T3
p = 0.23
Survival Probability
1.00
0.75
0.50
0.25
0.00
Number at risk
50 27 20 18
51 28 26 25
48 28 25 24
0
30
60
90
Days
Mortality Rate
42%
41%
25%
40%
30%
20%
10%
0%
CCL11 Tertile
Count
56.3
113.4
15
10
5
0
8
16
32
64
128
256
512
CSF CCL11 (pg/mL)


# IL-23

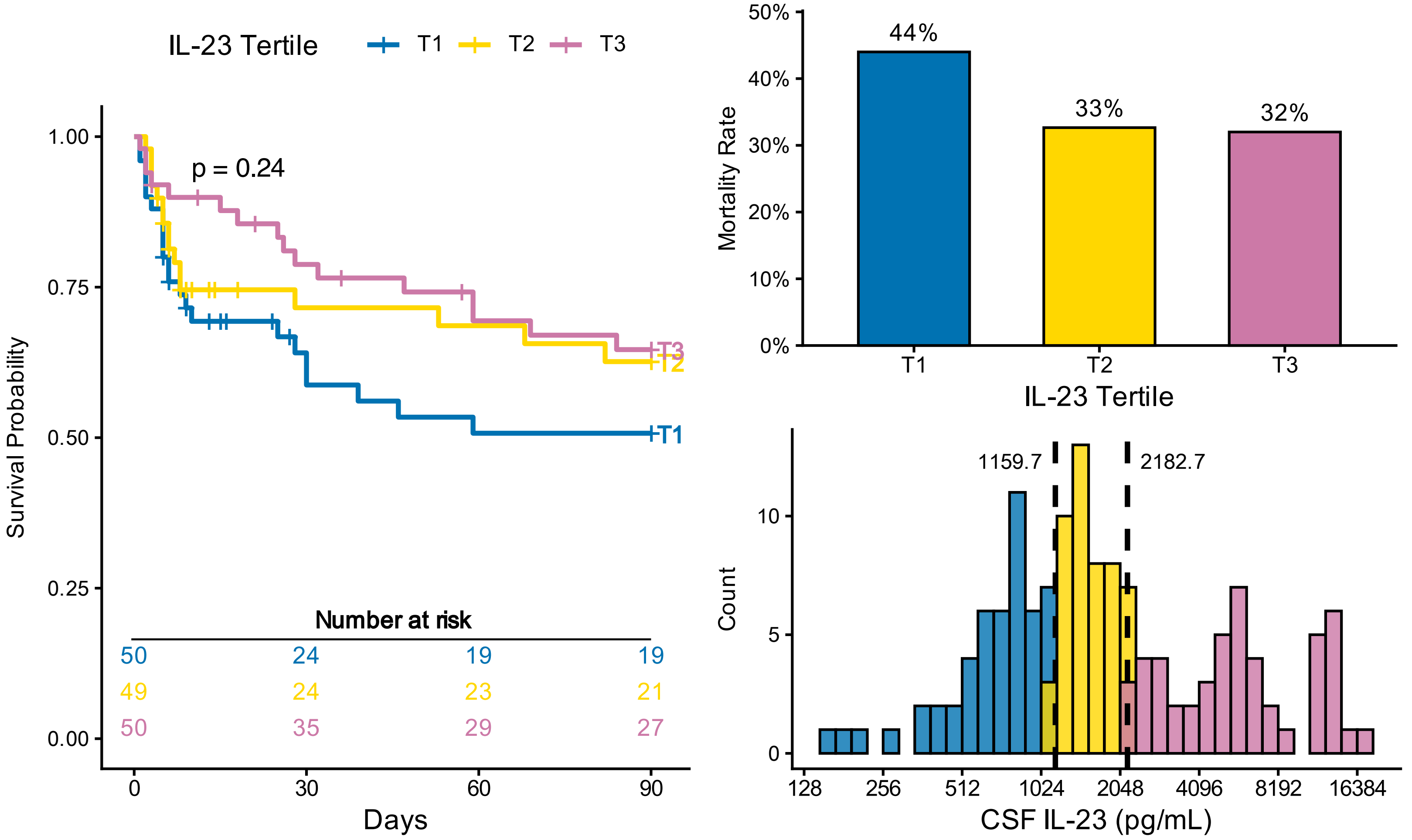

IL-23 Tertile
T1
T2
T3
p = 0.24
Survival Probability
1.00
0.75
0.50
0.25
0.00
Number at risk
50 24 19 19
49 24 23 21
50 35 29 27
0
30
60
90
Days
Mortality Rate
44%
33%
32%
50%
40%
30%
20%
10%
0%
IL-23 Tertile
Count
1159.7
2182.7
10
5
0
128
256
512
1024
2048
4096
8192
16384
CSF IL-23 (pg/mL)

# IL-3

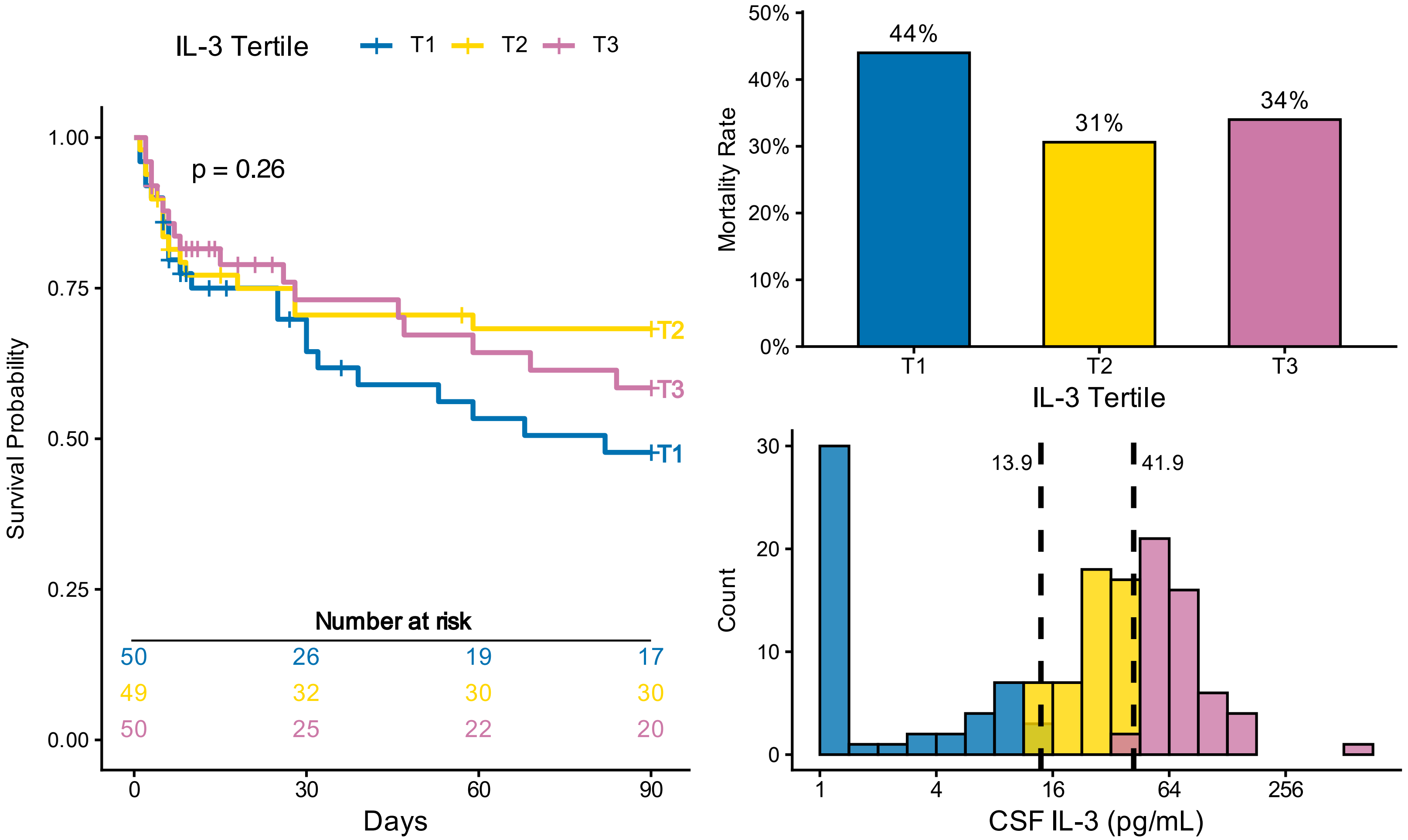

IL-3 Tertile
T1
T2
T3
p = 0.26
Survival Probability
1.00
0.75
0.50
0.25
0.00
Number at risk
50 26 19 17
49 32 30 30
50 25 22 20
0
30
60
90
Days
Mortality Rate
44%
31%
34%
T1
T2
T3
IL-3 Tertile
Count
13.9
41.9
CSF IL-3 (pg/mL)
1
4
16
64
256


# IL-15

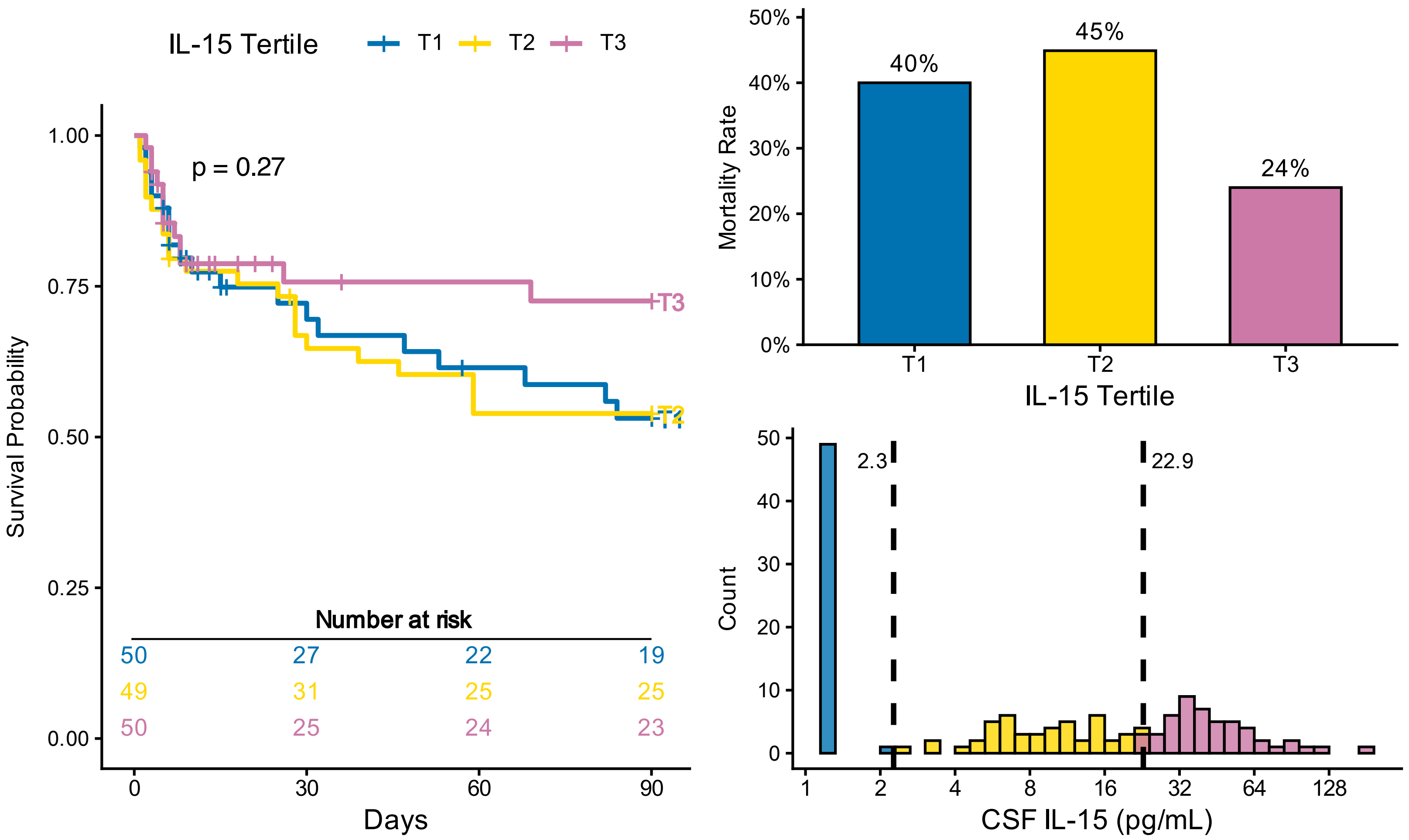

IL-15 Tertile
T1
T2
T3
p = 0.27
Survival Probability
Number at risk
50 27 22 19
49 31 25 25
50 25 24 23
Days
Mortality Rate
40%
45%
24%
IL-15 Tertile
Count
2.3
22.9
CSF IL-15 (pg/mL)
1
2
4
8
16
32
64
128

### VEGF

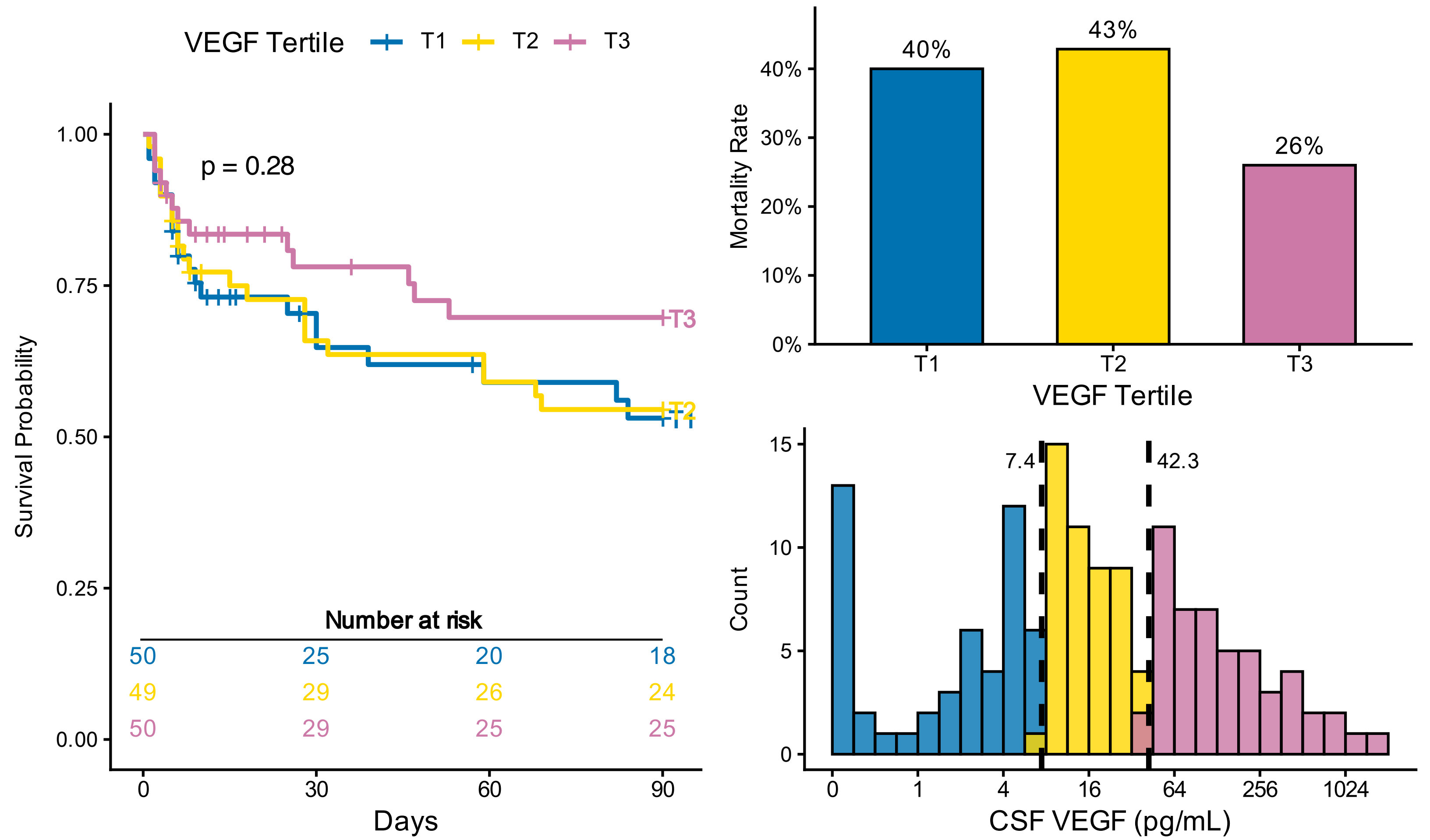

VEGF Tertile
T1
T2
T3
p = 0.28
Survival Probability
1.00
0.75
0.50
0.25
0.00
Number at risk
50 25 20 18
49 29 26 24
50 29 25 25
Days
0 30 60 90
Mortality Rate
40%
43%
26%
0% 10% 20% 30% 40%
VEGF Tertile
7.4
42.3
Count
0 5 10 15
0 1 4 16 64 256 1024
CSF VEGF (pg/mL)


# IL-18

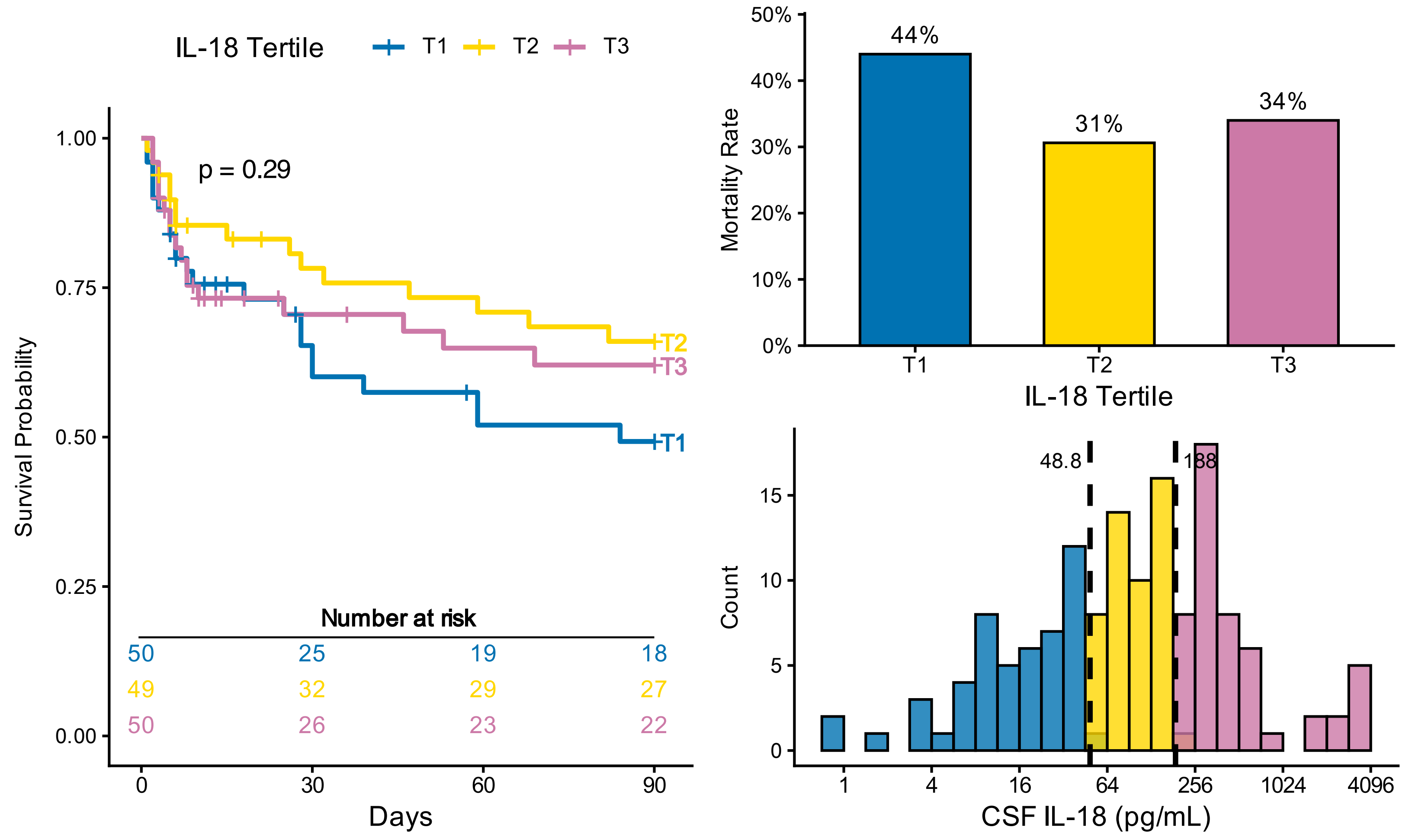

IL-18 Tertile
T1
T2
T3
p = 0.29
Survival Probability
1.00
0.75
0.50
0.25
0.00
Number at risk
50 25 19 18
49 32 29 27
50 26 23 22
Days
0 30 60 90
Mortality Rate
44%
31%
34%
0% 10% 20% 30% 40% 50%
IL-18 Tertile
48.8
188
Count
0 5 10 15
1 4 16 64 256 1024 4096
CSF IL-18 (pg/mL)

# IL-27

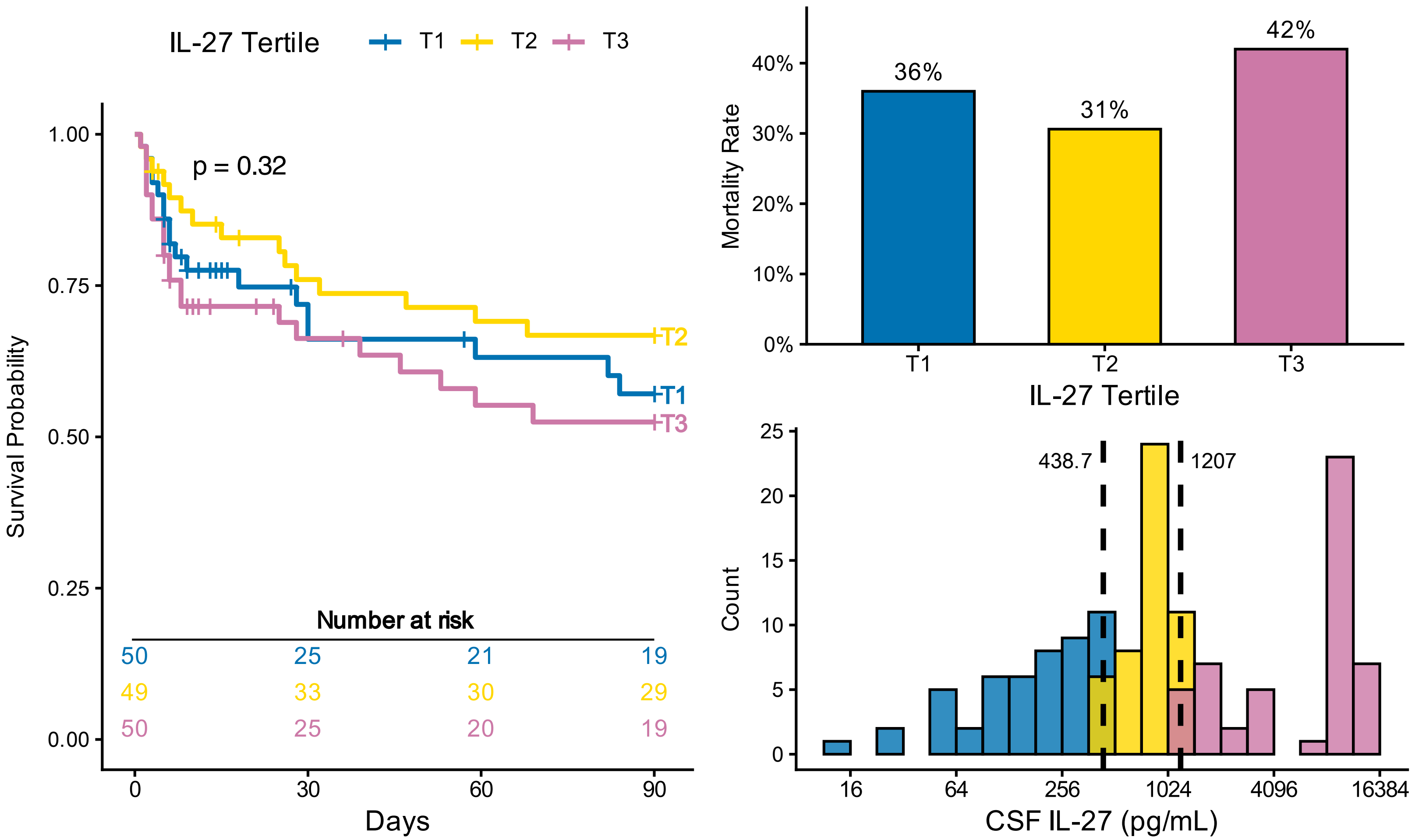
IL-27 Tertile T1 T2 T3
p = 0.32
Survival Probability
Number at risk
50 25 21 19
49 33 30 29
50 25 20 19
Days
Mortality Rate
36%
31%
42%
IL-27 Tertile
Count
438.7
1207
CSF IL-27 (pg/mL)

# IL-8

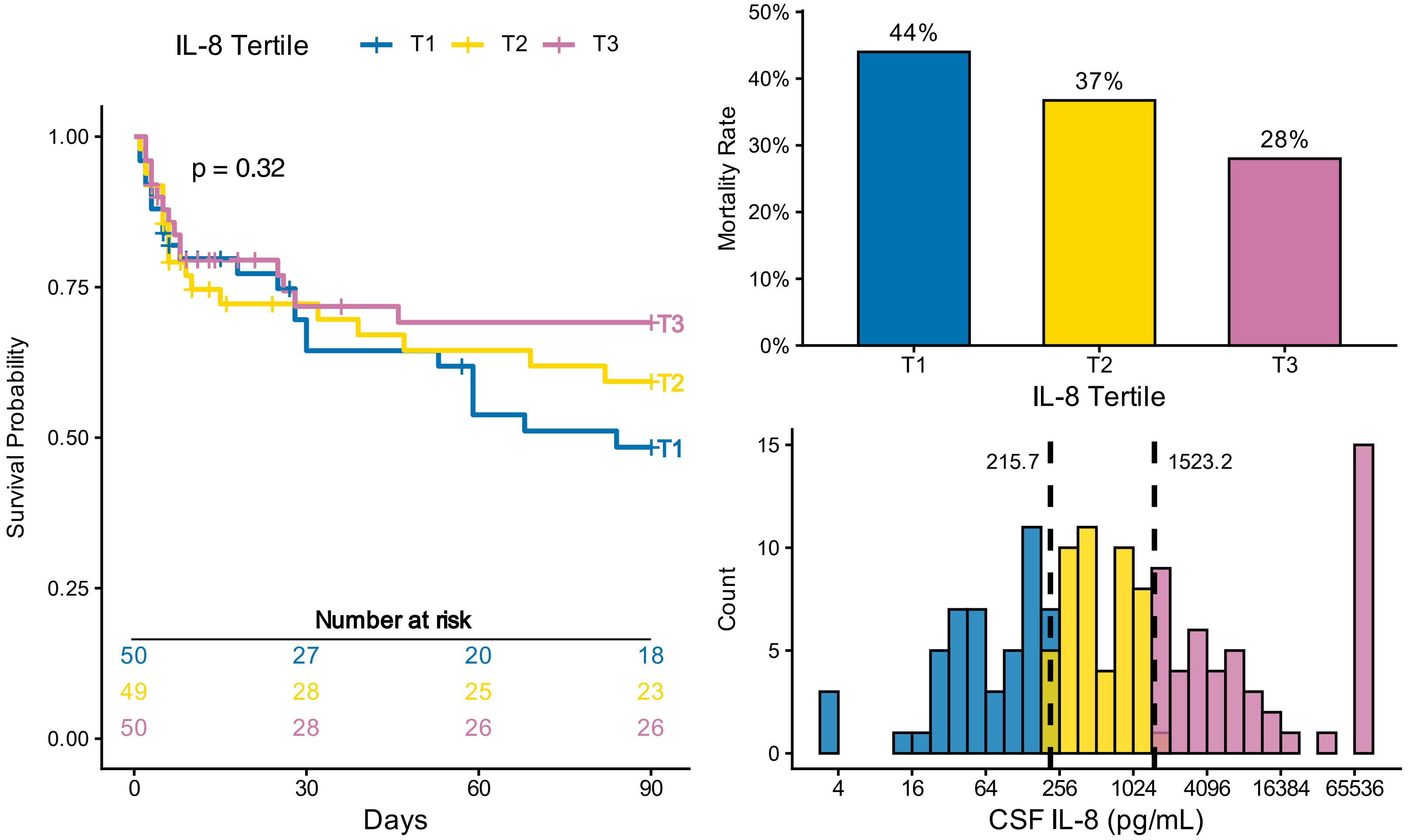
IL-8 Tertile T1 T2 T3
p = 0.32
Survival Probability
Number at risk
50 27 20 18
49 28 25 23
50 28 26 26
Days
Mortality Rate
44%
37%
28%
IL-8 Tertile
Count
215.7
1523.2
CSF IL-8 (pg/mL)

# IL-17A

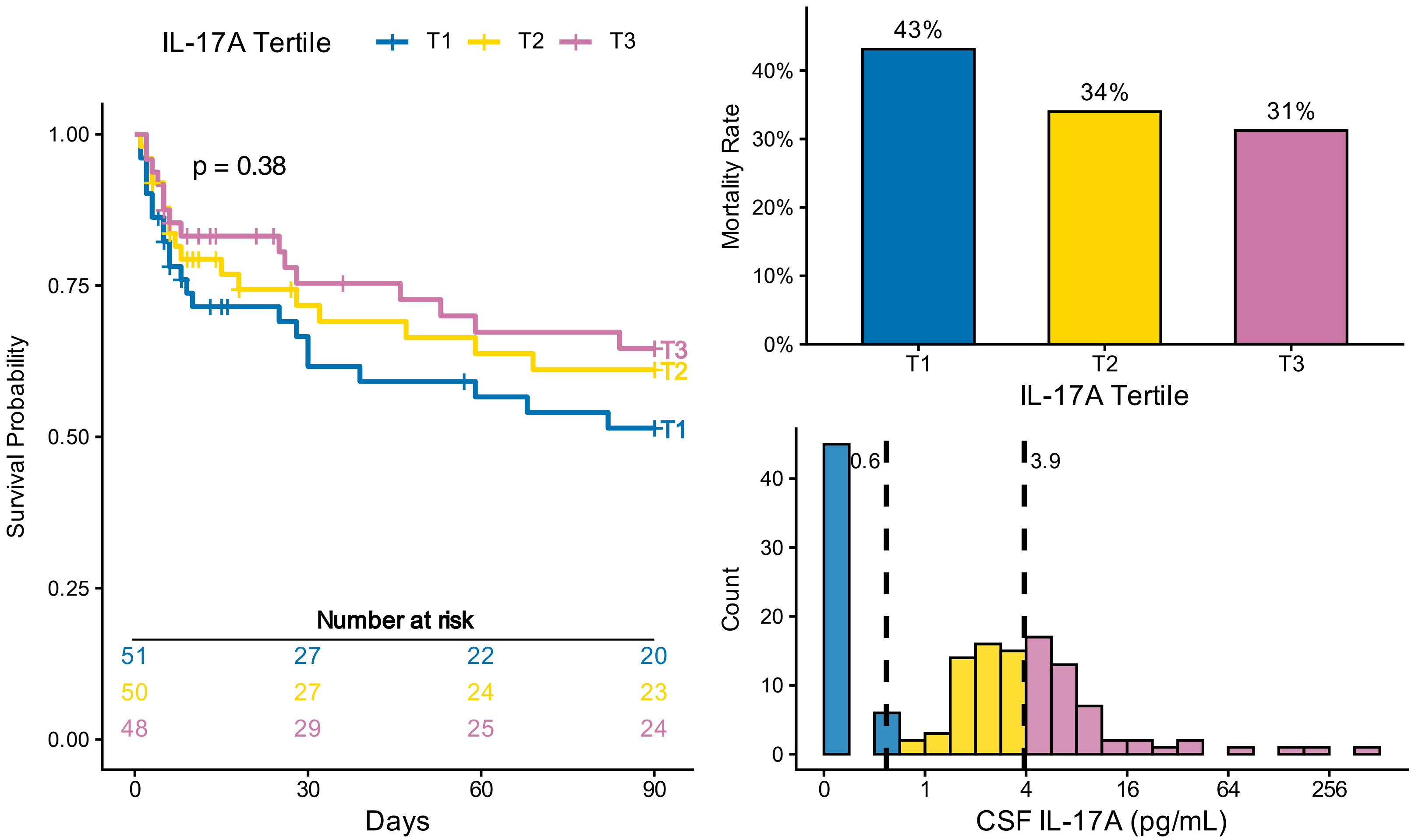
IL-17A Tertile
T1
T2
T3
p = 0.38
Survival Probability
1.00
0.75
0.50
0.25
0.00
Number at risk
51 27 22 20
50 27 24 23
48 29 25 24
0 30 60 90
Days
Mortality Rate
43%
34%
31%
40%
30%
20%
10%
0%
IL-17A Tertile
Count
0.6
3.9
40
30
20
10
0
0 1 4 16 64 256
CSF IL-17A (pg/mL)

### CCL4

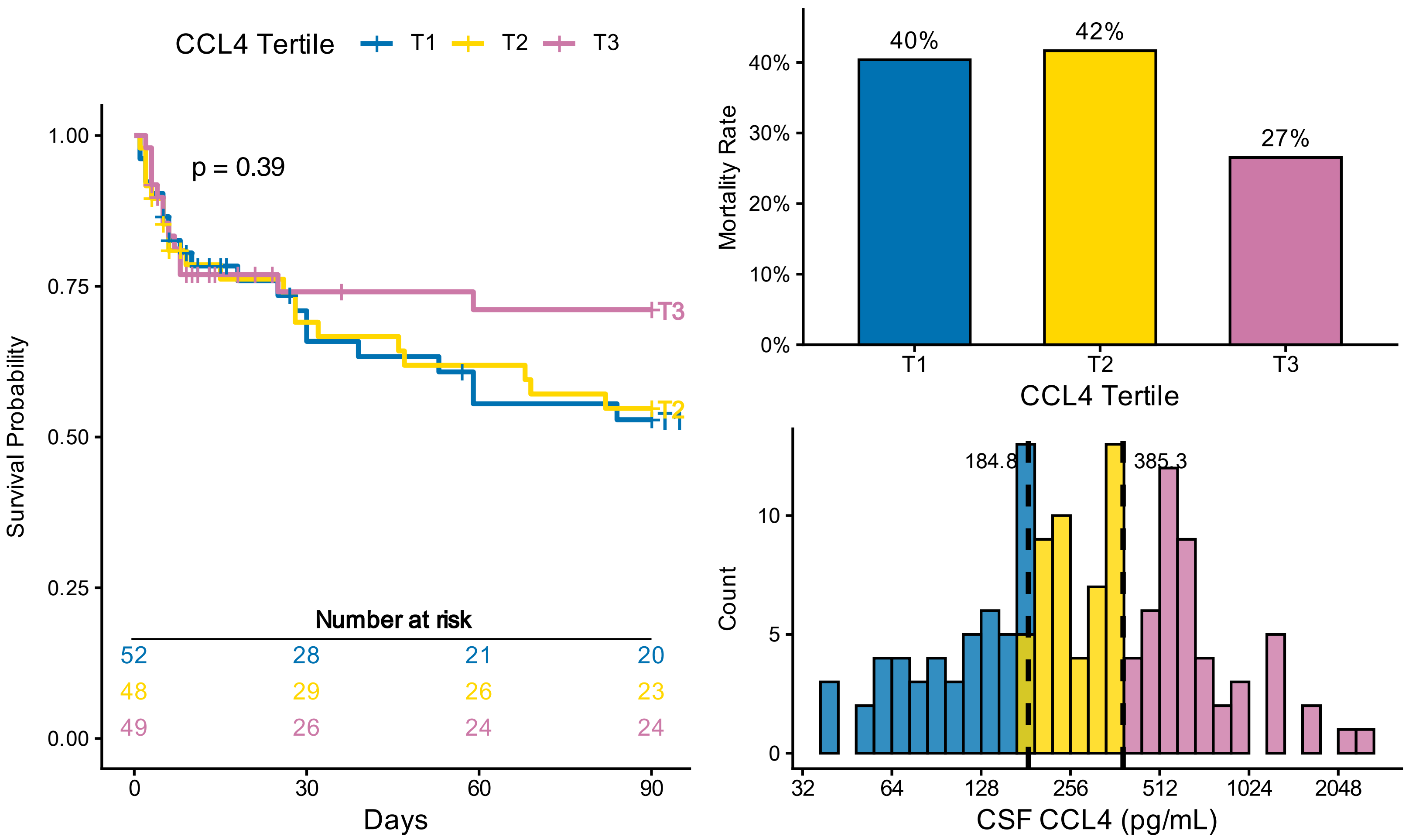
CCL4 Tertile
T1
T2
T3
p = 0.39
Survival Probability
1.00
0.75
0.50
0.25
0.00
Number at risk
52 28 21 20
48 29 26 23
49 26 24 24
0 30 60 90
Days
Mortality Rate
40%
42%
27%
40%
30%
20%
10%
0%
CCL4 Tertile
Count
184.8
385.3
10
5
0
32 64 128 256 512 1024 2048
CSF CCL4 (pg/mL)

# IL-4

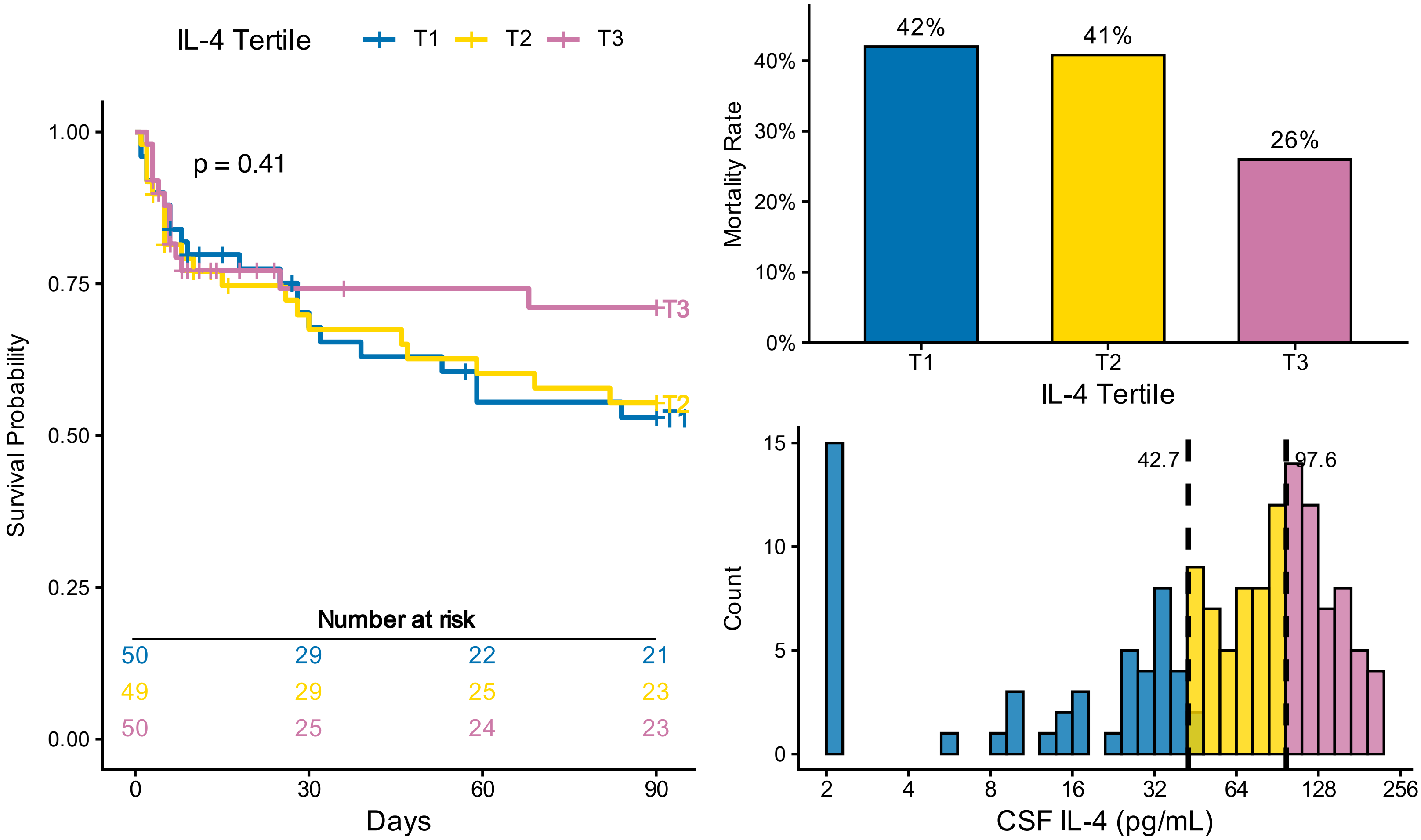
IL-4 Tertile
T1
T2
T3
p = 0.41
Survival Probability
1.00
0.75
0.50
0.25
0.00
Number at risk
50 29 22 21
49 29 25 23
50 25 24 23
0
30
60
90
Days
Mortality Rate
42%
41%
26%
40%
30%
20%
10%
0%
IL-4 Tertile
Count
42.7
97.6
15
10
5
0
2
4
8
16
32
64
128
256
CSF IL-4 (pg/mL)

# IL-6

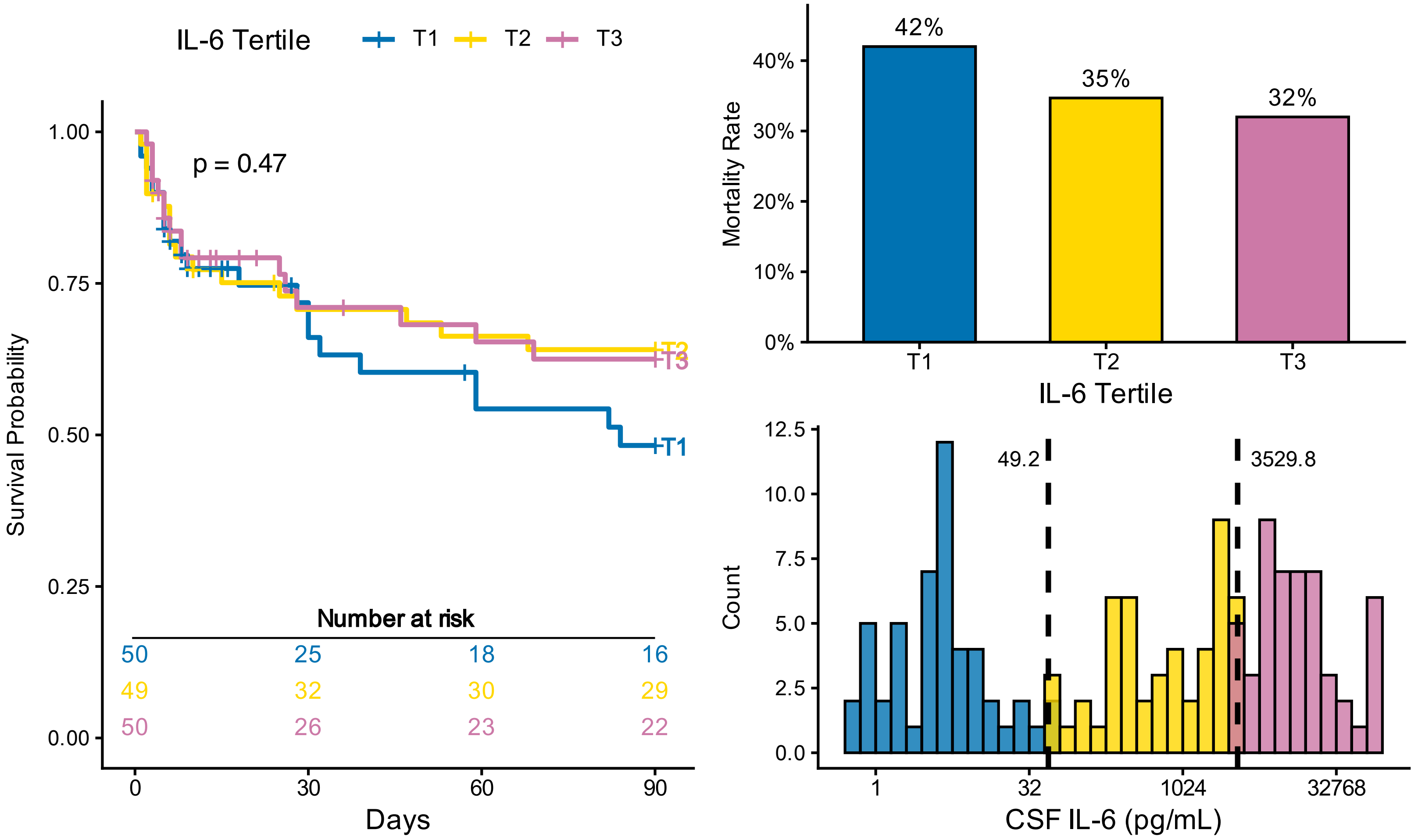
IL-6 Tertile
T1
T2
T3
p = 0.47
Survival Probability
1.00
0.75
0.50
0.25
0.00
Number at risk
50 25 18 16
49 32 30 29
50 26 23 22
0
30
60
90
Days
Mortality Rate
42%
35%
32%
40%
30%
20%
10%
0%
IL-6 Tertile
Count
49.2
3529.8
12.5
10.0
7.5
5.0
2.5
0.0
1
32
1024
32768
CSF IL-6 (pg/mL)

# IL-7

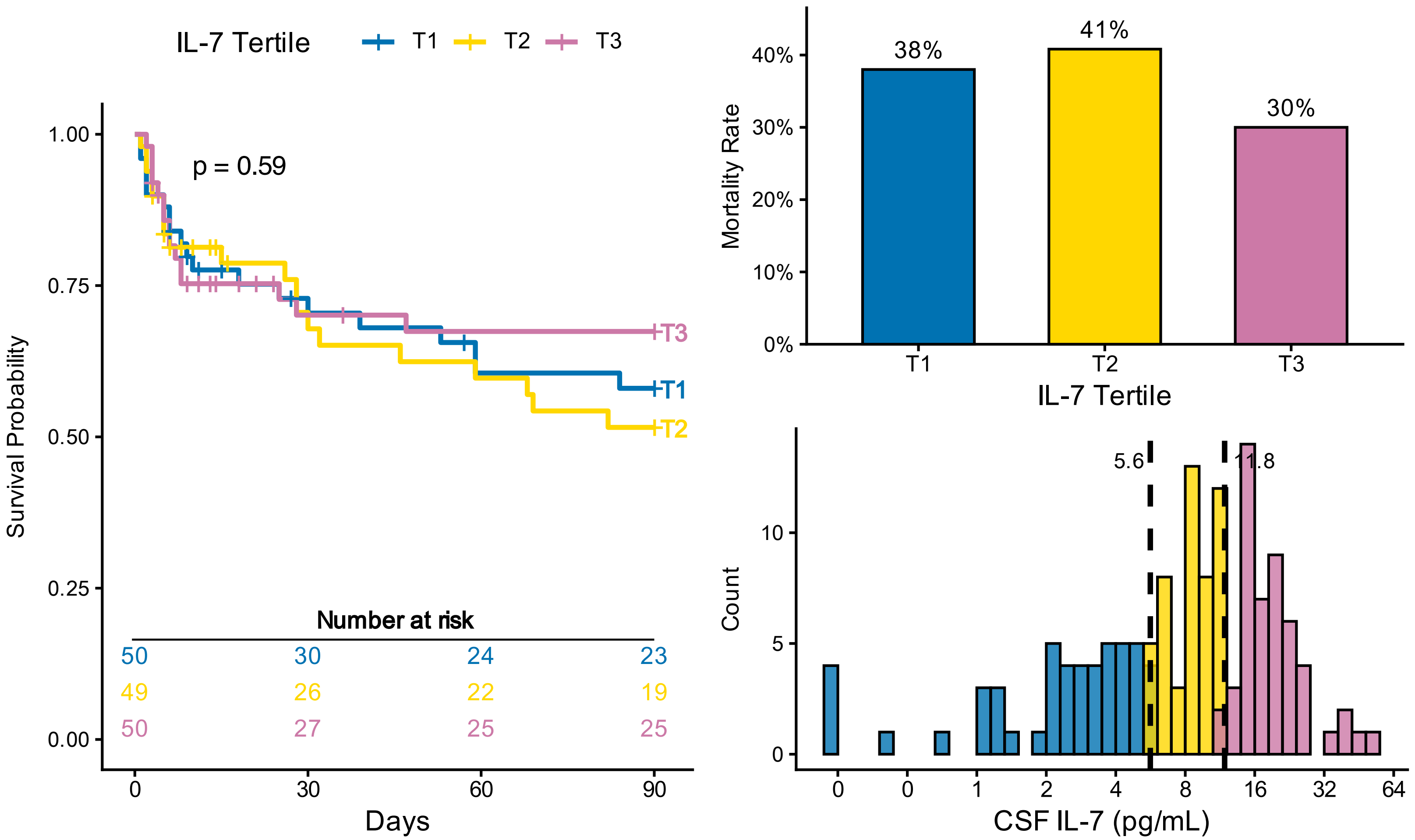

IL-7 Tertile
T1
T2
T3
p = 0.59
Survival Probability
1.00
0.75
0.50
0.25
0.00
Number at risk
50 30 24 23
49 26 22 19
50 27 25 25
0
30
60
90
Days
Mortality Rate
38%
41%
30%
40%
30%
20%
10%
0%
IL-7 Tertile
5.6
11.8
Count
CSF IL-7 (pg/mL)


# IL-21

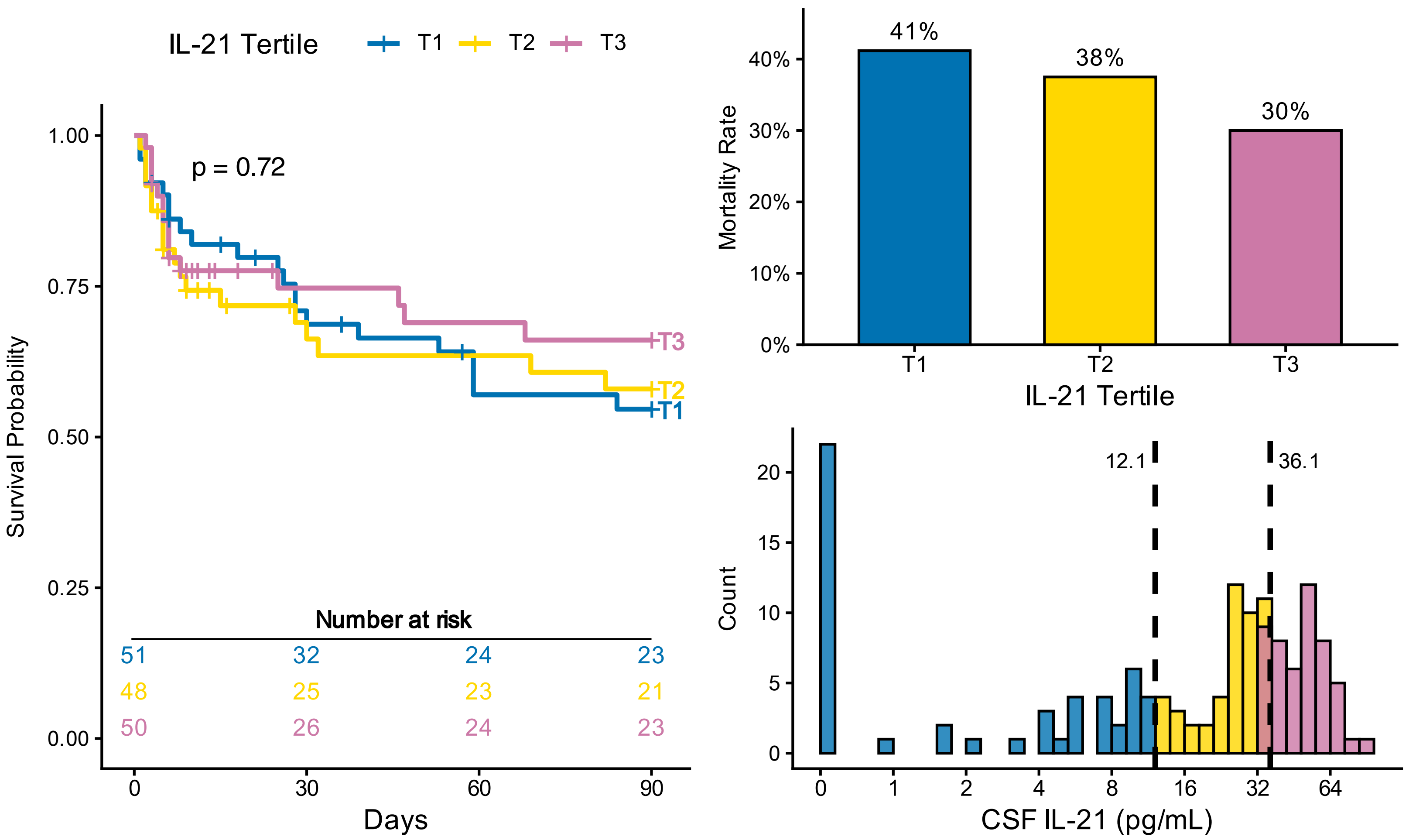

IL-21 Tertile
T1
T2
T3
p = 0.72
Survival Probability
1.00
0.75
0.50
0.25
0.00
Number at risk
51 32 24 23
48 25 23 21
50 26 24 23
0
30
60
90
Days
Mortality Rate
41%
38%
30%
40%
30%
20%
10%
0%
IL-21 Tertile
12.1
36.1
Count
CSF IL-21 (pg/mL)

# IL-1ra

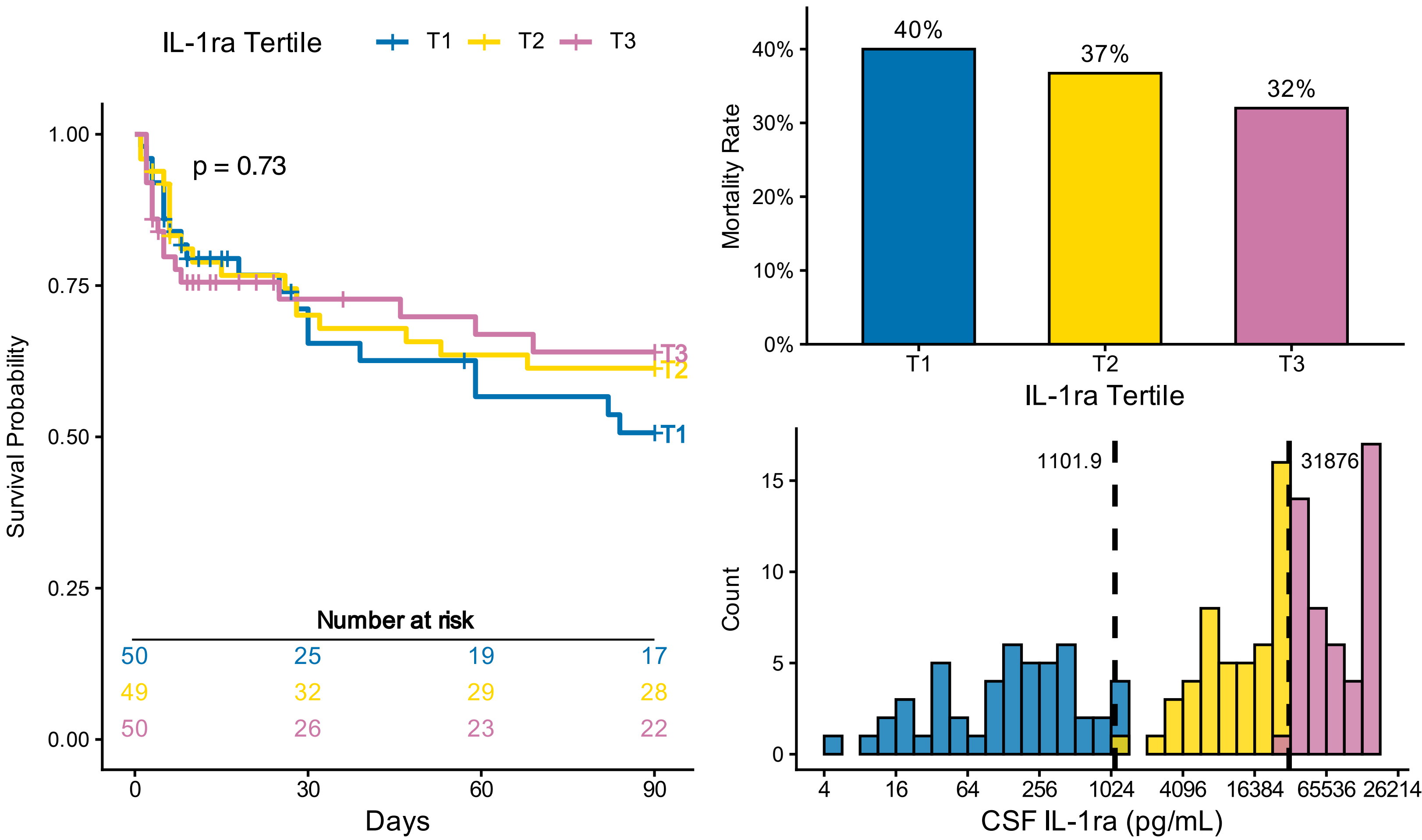
IL-1ra Tertile
T1
T2
T3
p = 0.73
Survival Probability
1.00
0.75
0.50
0.25
0.00
Number at risk
50 25 19 17
49 32 29 28
50 26 23 22
0
30
60
90
Days
Mortality Rate
40%
37%
32%
0%
10%
20%
30%
40%
T1
T2
T3
IL-1ra Tertile
Count
1101.9
31876
0
5
10
15
4
16
64
256
1024
4096
16384
65536
26214
CSF IL-1ra (pg/mL)

### G-CSF

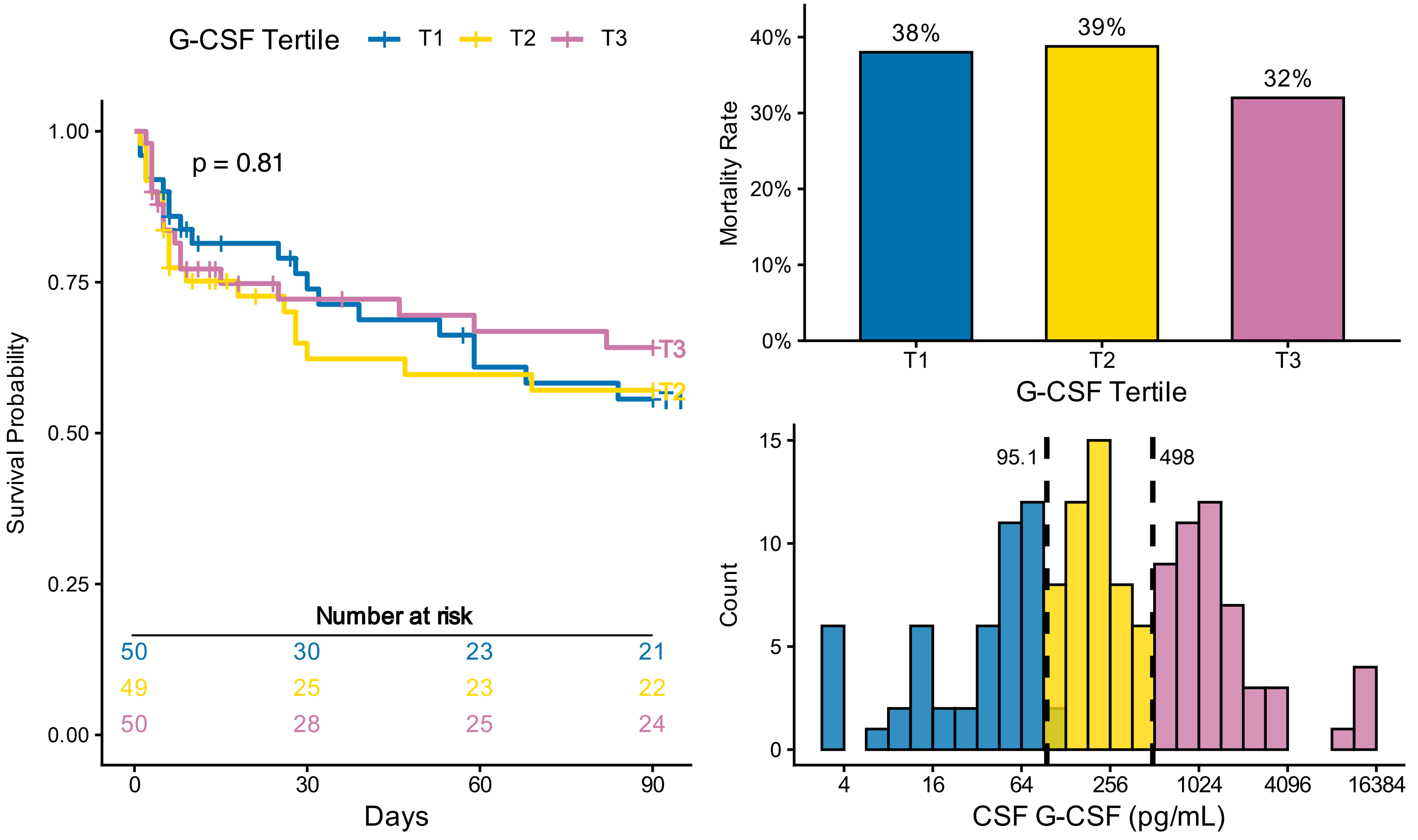
G-CSF Tertile
T1
T2
T3
p = 0.81
Survival Probability
1.00
0.75
0.50
0.25
0.00
Number at risk
50 30 23 21
49 25 23 22
50 28 25 24
0
30
60
90
Days
Mortality Rate
38%
39%
32%
0%
10%
20%
30%
40%
T1
T2
T3
G-CSF Tertile
Count
95.1
498
0
5
10
15
4
16
64
256
1024
4096
16384
CSF G-CSF (pg/mL)

# IL-5

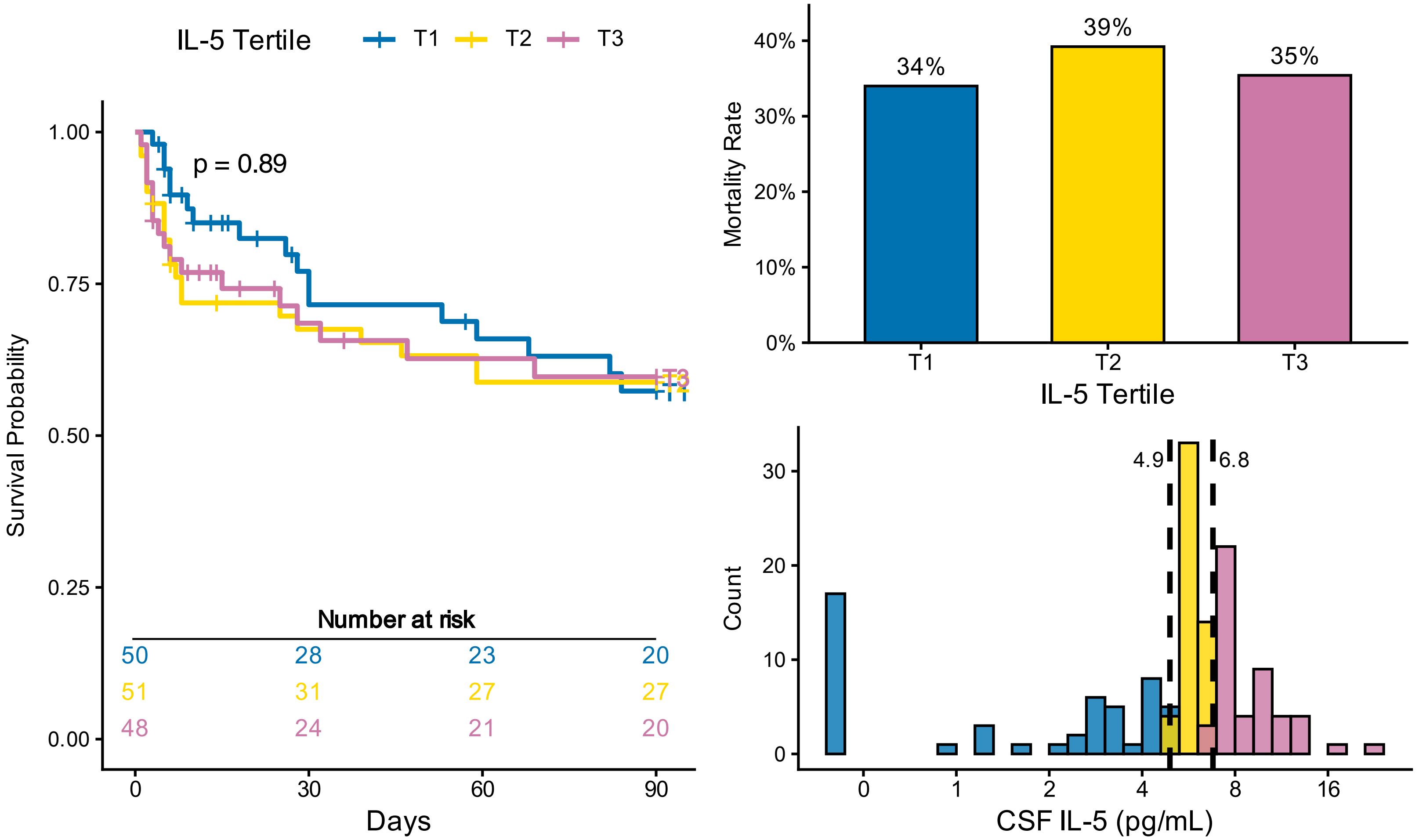
IL-5 Tertile
T1
T2
T3
p = 0.89
Survival Probability
1.00
0.75
0.50
0.25
0.00
Number at risk
50 28 23 20
51 31 27 27
48 24 21 20
0
30
60
90
Days
Mortality Rate
40%
30%
20%
10%
0%
34%
39%
35%
IL-5 Tertile
Count
4.9
6.8
30
20
10
0
0
1
2
4
8
16
CSF IL-5 (pg/mL)

### CCL2

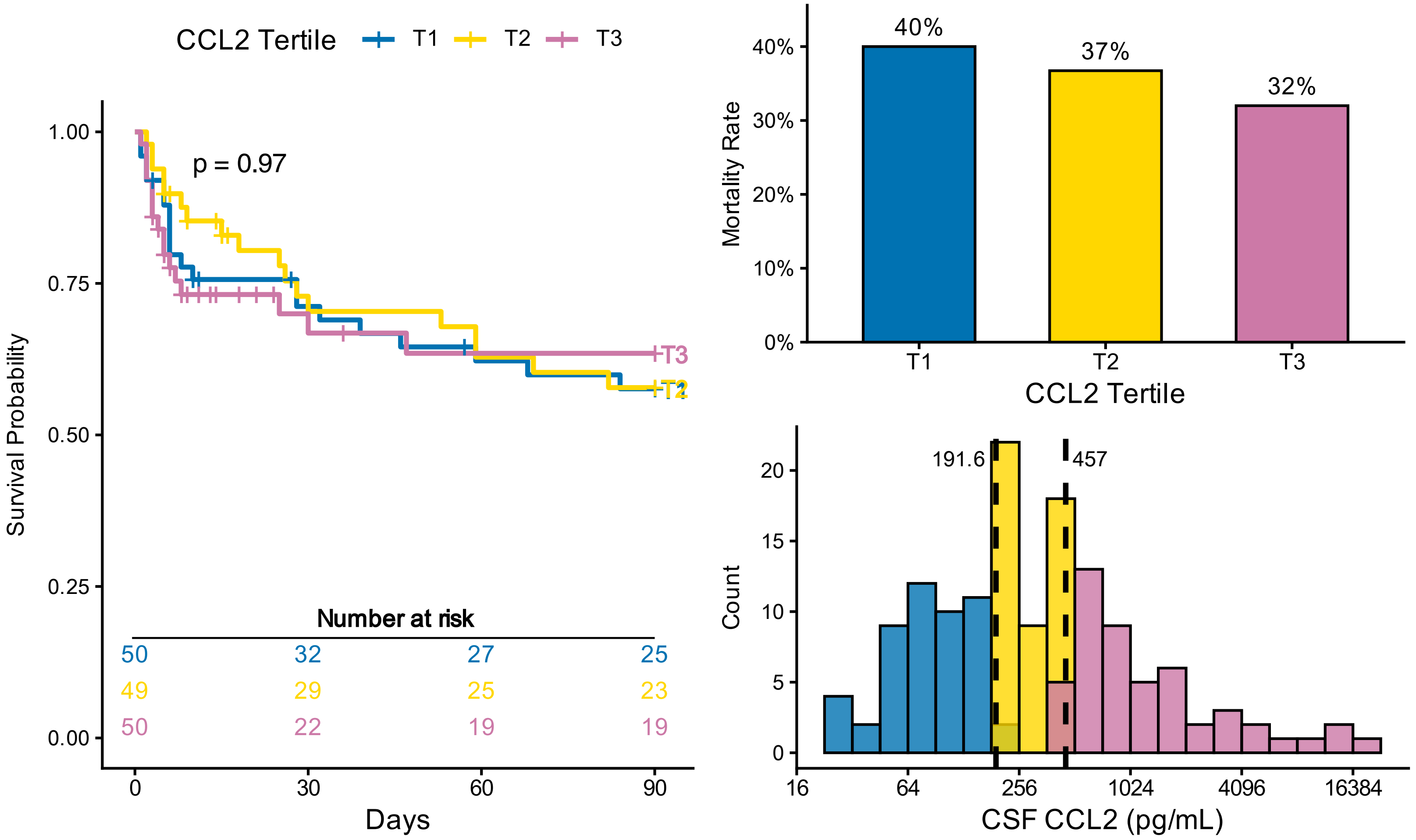
CCL2 Tertile
T1
T2
T3
p = 0.97
Survival Probability
1.00
0.75
0.50
0.25
0.00
Number at risk
50 32 27 25
49 29 25 23
50 22 19 19
0
30
60
90
Days
Mortality Rate
40%
30%
20%
10%
0%
40%
37%
32%
CCL2 Tertile
Count
191.6
457
20
15
10
5
0
16
64
256
1024
4096
16384
CSF CCL2 (pg/mL)
